# Quantifying life-expectancy Losses and Gains over 31 years (1990-2021): A population-level study on West African Countries

**DOI:** 10.1101/2024.05.29.24308136

**Authors:** David Lagoro Kitara, Joelle Abi abboud, Ritah Nantale, Camille Lassale, Emmanuel Olal, Gaye Bamba

## Abstract

**Background:** Life expectancy at birth (LE_0_) is one of the most widely used indicators for determining the overall development of a country. Worldwide, LE_0_ has increased over the last ten years in most countries. This parameter is essential for developing countries as they strive to achieve socio-economic progress by investing significantly in social sectors like social services and safety nets. LE_0_ among West African countries has consistently been the lowest in African regions, which calls for more research. This study aimed to quantify LE_0_ losses and gains by sex in West African countries from 1990 to 2021. We hypothesize that lower life expectancy rates in the West African countries were likely due to poorer socioeconomic indicators than in other African regions.

**Methods:** Life tables by sex and country were calculated for sixteen (16) West African countries from 1990 to 2021. LE_0_ for 1990, 2000, 2010, and 2020 were contextualized alongside recent trends between the two sexes and country. We used decomposition techniques to examine which sex and country contributed to gains and losses in LE_0_ between 1990 and 2021. RStudio software was used to calculate differences in LE_0_ from one year to another. In addition, linear regression, life disparity, and the Gini coefficient were used to trace the evolution of LE_0_ in the last three decades in West Africa.

**Results:** There were LE_0_ gains from 1990 to 2021 in all sixteen West African countries (Benin, Burkina Faso, Cape Verde, Ivory Coast, Gambia, Ghana, Guinea, Guinea Bissau, Liberia, Mali, Mauritania, Niger, Nigeria, Senegal, Sierra Leone, and Togo). The highest LE_0_ gains between 1990 and 2021 were observed in Guinea-Bissau among males (28.32 years), followed by females in Niger (20.41 years), followed by males in Mauritania (18.91 years), and females in Liberia (18.13 years). The least LE_0_ gains were observed in males in Mali (4.48 years). Most West African countries achieved the highest LE in 2019 (14/16, 75.0%), except for Cape Verde in 2017 (5.28), and Togo in 2021 (0.94).

**Conclusion:** Although most West African countries posted progressive LE_0_ gains from 1990 to 2021, there were LE_0_ losses in 2020 and 2021 when the COVID-19 pandemic emerged. The West African region has the lowest LE_0_ of all African regions probably due to lower socio-economic indicators compared to all other African regions. Also, during the COVID-19 pandemic in 2020 and 2021, there were LE_0_ losses in all West African countries, except Togo. In addition, LE_0_ gaps between males and females were highest in the late 1990s and least during the late 2000s. Even though several studies reported that morbidity and mortality rates of COVID-19 were lower in Africa than in the rest of the world, a more comprehensive study is warranted to assess the actual impact of COVID-19 on West African countries.

## Introduction

Life expectancy at birth (LE_0_) is one of the most widely used population health and longevity metrics.^1,2^ It refers to the average number of years a synthetic cohort of newborns would live if they were to experience the death rates observed in a given period throughout their lifespan.^1,2^ Life expectancy is calculated as conditional on surviving to a given age, e.g., 60 years, which refers to the remaining life expectancy from age 60 years.^1,2^ The study on life expectancy in the context of the COVID-19 pandemic matters because it enables us to compare the cumulative impacts of the pandemic against past mortality shocks and recent trends across different countries using a standardized indicator routinely monitored to capture differences in mortality.^1,2,3,4^

Also, life expectancy is a statistical measure of the average number of years a person is expected to live, and it is an indicator for measuring the well-being of a population.^4^ Factors contributing to life expectancy values include educational level, access to the quality healthcare systems, economic empowerment, and health behaviors, among others.^5^

The average life expectancy in Africa in 2019 was 61 years for males and 65 years for females.^6^ In addition, life expectancy values for the Northern Africa region have been higher than those from other regions of Africa.^7^ Many African countries, especially West African countries, have lower life expectancies because they are still grappling with high under-five mortality rates, extreme poverty, hunger, high levels of illiteracy, lack of access to quality medical care, environmental hazards, HIV and AIDS, malaria, road accidents, conflicts, wars, lifestyle diseases, among others.^7,8,9,10,11^

As of August 2^nd^, 2023, there were 769 million confirmed COVID-19 cases, with 6.95 million deaths resulting from infection with the severe-acute-respiratory-syndrome-corovirus-2 (SARS-CoV-2) around the world.^12^ This estimate, although astounding, disguises the uneven impact of the pandemic across different countries and demographic characteristics like age and sex^13^, as well as its impact on population health, years of life lost^14^, and longevity.^15^

Moreover, variations in testing capacity coupled with definitional inconsistencies in counting COVID-19 deaths make the actual global toll of COVID-19 infections challenging to estimate accurately.^2^

In order to address these measurement challenges worldwide, significant efforts have been directed at harmonizing and analyzing all-cause mortality data.^1,2^ In addition, a widely used approach to quantify the burden of the pandemic using all-cause mortality is through the analysis of excess mortality, defined as the number of deaths observed during the pandemic above a baseline of recent trends.^1,2^ Before the COVID-19 pandemic, life expectancy at birth typically increased almost monotonically in most countries over the twentieth and into the twenty-first centuries.^15,16^ In recent decades, improvements in life expectancy among high-income countries were predominantly driven by gains made at older ages (≥65 years)^17^, although significant cross-country heterogeneity persists.^15,16,17^ This heterogeneity has become more prominent since 2010.^15,16,17^ For example, the life expectancy in the USA^18^, England & Wales, and Scotland saw only limited gains in the last decade.^19,20^ These uncharacteristic trends have been linked to slower improvements in old-age mortality and increased working-age death rates.^19^ This study aimed to quantify life expectancy (LE_0_) losses and gains by sex in West African countries from 1990 to 2021.

We hypothesize that lower life expectancy rates in the West African countries were likely due to poorer socio-economic indicators compared to other regions of Africa.

## Materials and Methods

### Study sites

We conducted our study on sixteen West Africa countries (Nigeria, Niger, Togo, Benin, Ghana, Cote D’Ivoire, Sierra Leone, Gambia, Mauritania, Cape Verde, Mali, Guinea-Bissau, Burkina-Faso, Guinea, Liberia, and Gambia) (Figure 1).

**Figure 1:**
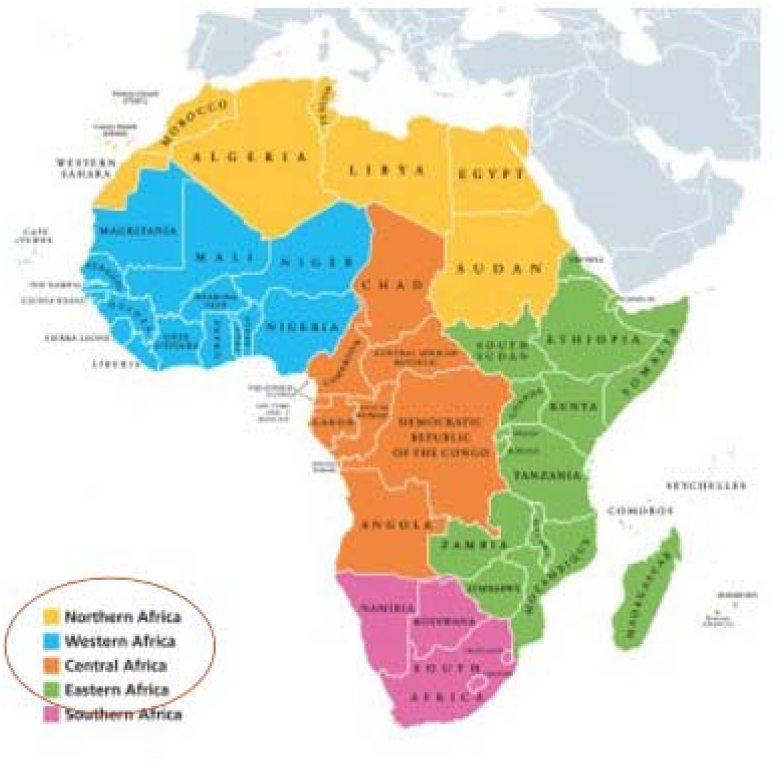
Map of Africa showing African countries by their regions (West Africa in blue).

### Data

Life table data, categorized by single age and sex (female, male, and both), were extracted from the World Population Prospects (WPPs) estimate for 2022.^21,22^ From these life tables, we extracted data for all West African countries. We extracted the data from 1990 to the latest available year at the time of writing in 2022.^21,22^

All life tables were downloaded on March 28^th^, 2023. Life tables on WPPs offer detailed information on the two sexes and countries, including single-age mortality rates up to one hundred years.^21,22^ It presents a comprehensive set of values, showcasing the mortality experience of a hypothetical cohort of infants born simultaneously and subjected to the specific mortality rates of a given year throughout their lifetime.^21,22^

The tables encompassed essential metrics, such as probabilities of dying (qx) and surviving (px), counts of individuals surviving (lx) and dying (dx), total person-years lived (Lx), survivorship ratios (Sx), cumulative stationary population (Tx), average remaining life expectancy (ex), and average number of years lived (ax).^21,22^

### Life expectancy values

In this study, we utilized life expectancy (ex) measures from life tables to observe the evolution of (ex) over the last thirty-one years. Additionally, we focused on life expectancy at age 0 (ex0) as it is the most commonly used metric for determining population health and longevity.^21,23^ We calculated differences in (ex0) to compare life expectancy evolution between countries, years, and sexes.

### Statistical Analyses

For descriptive analysis, the life table was divided into two categories: one for West African males and another for females. These categories were then used to characterize life expectancy at birth and 60 years using means and standard deviations (Sd). A linear regression model, considering life expectancy as a continuous variable, was used to estimate the relationship between age, years, sex, and life expectancy for every country. Additionally, a sex-stratified analysis was conducted to examine the impact of gender on life expectancy. Furthermore, we set a separate group of correlation analyses to investigate the relationship between years, age, and life expectancy. In addition, life disparity^24^ and Gini coefficient^25^ were used to trace the evolution of life expectancy in the last three decades in West African countries.

In this, we defined life disparity as a measure of how much lifespans differ among individuals.^24^ It defines death as premature if postponing it to a later age would decrease life disparity.^24^ On the other hand, we defined the Gini coefficient (Gini index) as a synthetic indicator that captures the level of inequality for a given variable and population.^25^ It varies between 0 (perfect equality) and 1 (extreme inequality). Between 0 and 1, the higher the Gini index, the greater the inequality.^25^ Overall, the statistical analyses were conducted using RStudio version 4.2.2.^26,27^

### The formulae for the Linear Regressions Analyses were

Differences in LE_0_ between female and male=β_0_+β_1_ Year+β_2_ Region+ε (1)

The formula for the linear regression is ex=β_0_+β_1_ Year+β_2_ age+β_3_ sex+ε (2)

## Results

Our study was conducted on sixteen West African countries (Nigeria, Niger, Togo, Benin, Ghana, Cote D’Ivoire, Sierra Leone, Gambia, Mauritania, Cape Verde, Mali, Guinea Bissau, Burkina Faso, Guinea, Liberia, and Gambia) (Figure 1). The most substantial findings were the life expectancy (LE_0_) gains and losses from 1990 to 2021 in the sixteen West African countries.

In our data set, life expectancy at birth (LE_0_) among females in 1990 in West Africa ranged from the highest at 66.58 years in Cape Verde to the least at 42.04 years in Niger (Table 1). Among males in 1990, it ranged from 63.17 in Cape Verde to 31.21 years in Liberia. Thirty years later, in 2020, the highest life expectancy (LE_0_) among West African countries was in Cape Verde at 79.16 years and the lowest in Nigeria at 52.46 years. Overall, females’ life expectancy has been higher than males in every country in West Africa (Table 1).

**Table 1:**
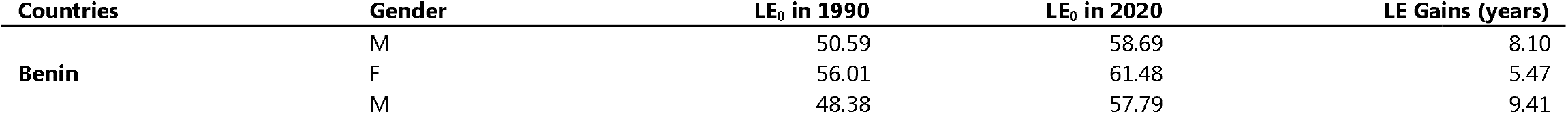
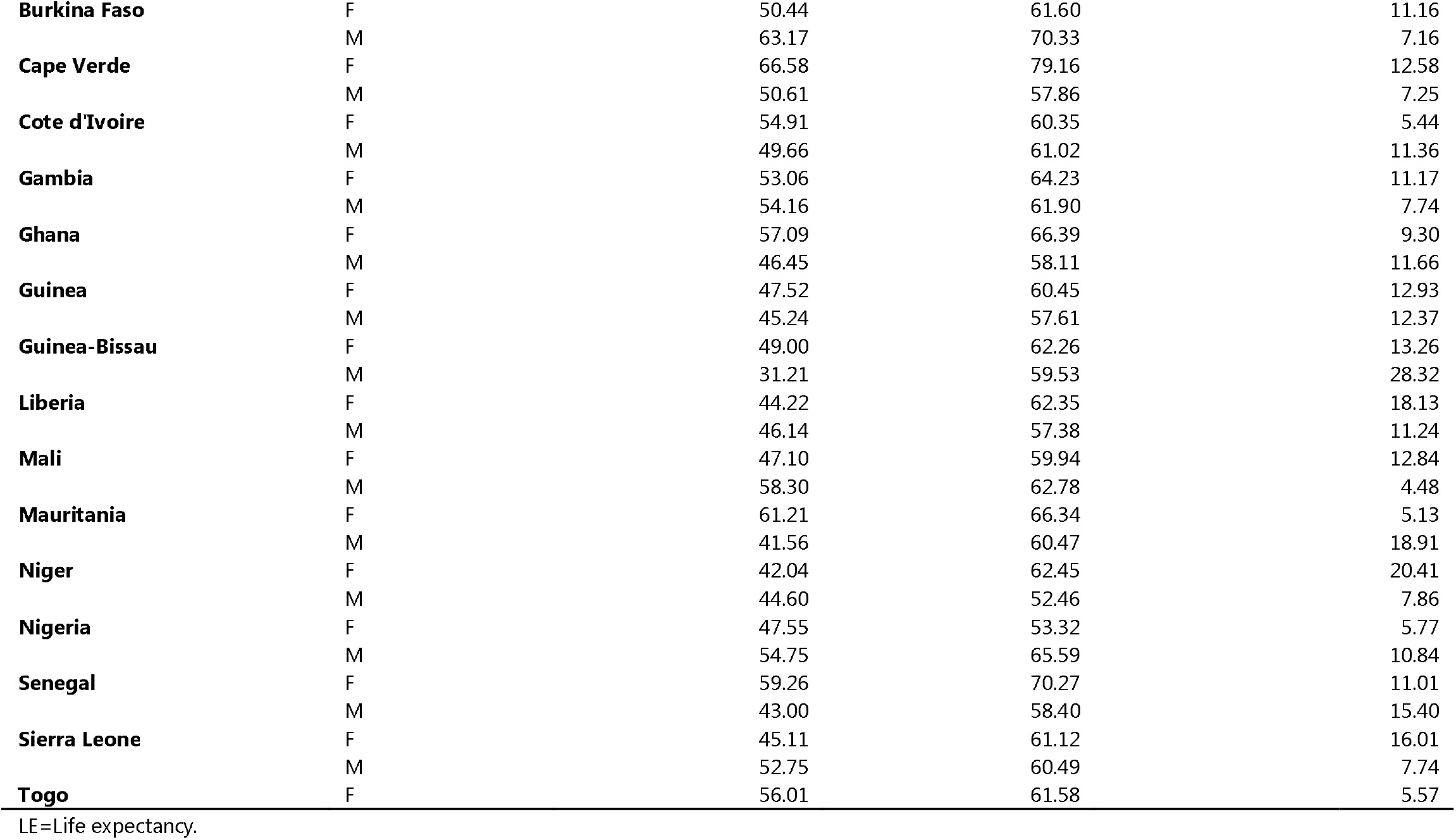
Life expectancy (LE_0_) gains among West African countries by gender from 1990 to 2020. In Table 1, the highest life expectancy (LE_0_) gains between 1990 and 2020 were observed in Guinea-Bissau among males (28.32 years), followed by females in Niger (20.41 years), followed by males in Mauritania (18.91 years) and females in Liberia (18.13 years). The least gains in Life expectancy were observed in males in mali (4.48 years).

A map of West African countries showing their life expectancies (LE_0_) from 1990 to 2020. These maps show life expectancy gains in most West Africa countries over thirty years. However, LE_0_ gains were slow, as can be seen in life expectancy in the 1990s between (40-60 years), while in 2020, it was between (50-65 years), except for Cape Verde, that had a LE_0_ of 65.02 years in the 1990s and 74.81 years in 2020 and this is the highest in West African regions (Figure 2).

**Figure 2:**
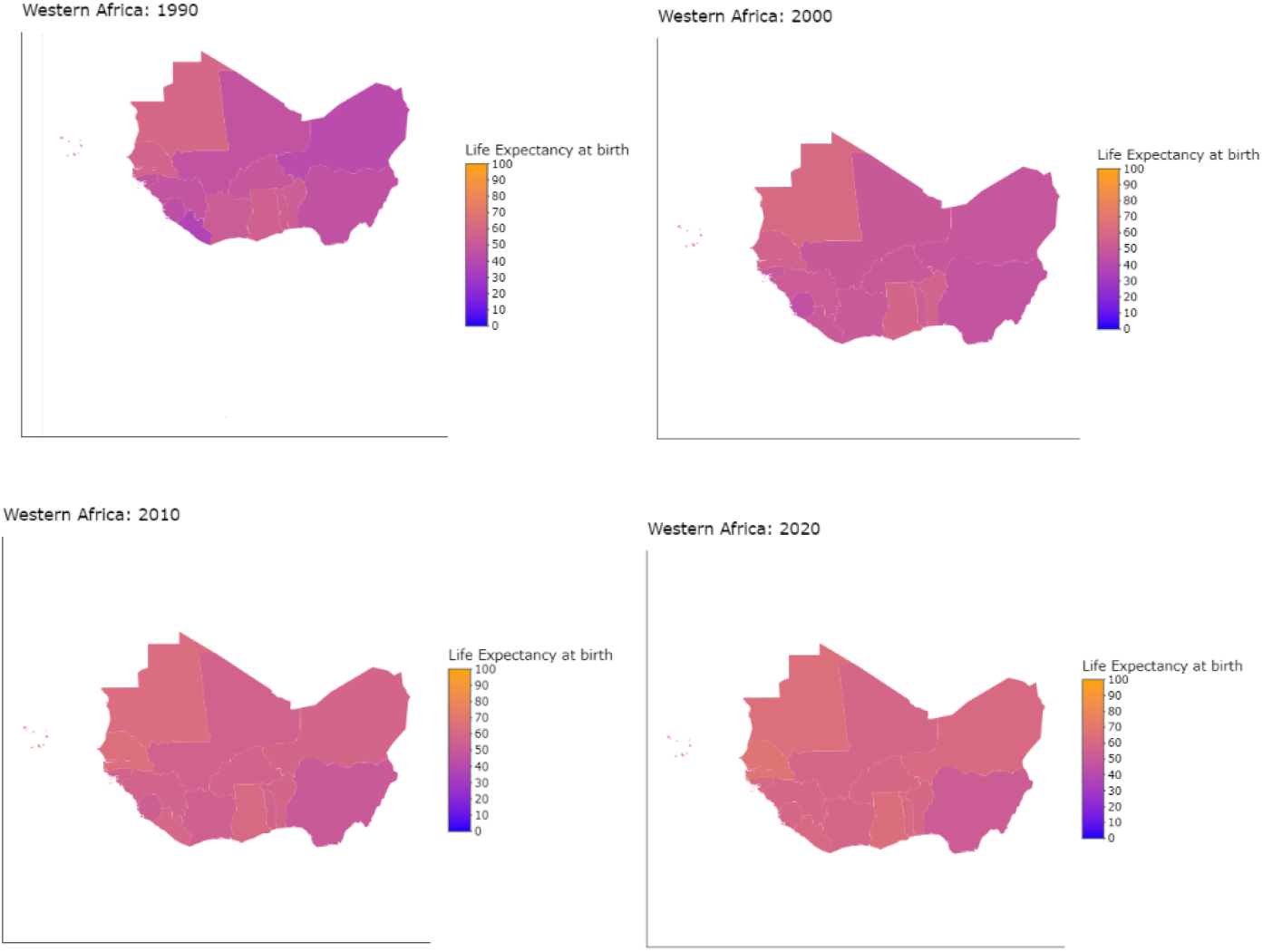
Changes in life expectancy (LE_0_) in West Africa in 1990, 2000, 2010 and 2020. Figure 2 are maps of West African countries and their life expectancies (LE_0_) from 1990, 2000, 2010, and 2020. These maps show life expectancy gains in most West African countries over the 30 years. However, LE_0_ increases were slow, as can be seen in life expectancy in 1990 of 40-60 years, while in 2020, it was between 50-65 years, except for Cape Verde, that have an LE_0_ of 66.58 years in 1990 and at 79.16 years in 2020 and it is the highest in West Africa.

In addition, yearly changes in life expectancy at birth for West African countries were drawn on graphs. These graphs calculated yearly changes in LE_0_ using the formula (yearly change=LEi-LEi-1 with i = year). These graphs show that 2020 LE_0_ decreased compared to the previous year, except in Mali and Sierra Leone, which increased in 2021. In males and females in Togo, LE_0_ did not decrease during the COVID-19 pandemic. However, female LE_0_ increased in Niger in 2021 (0.34 compared to 2020), and for males, it increased (0.6), and the increase of LE_0_ in 2021 for females is seen (Figure 3).

**Figure 3:**
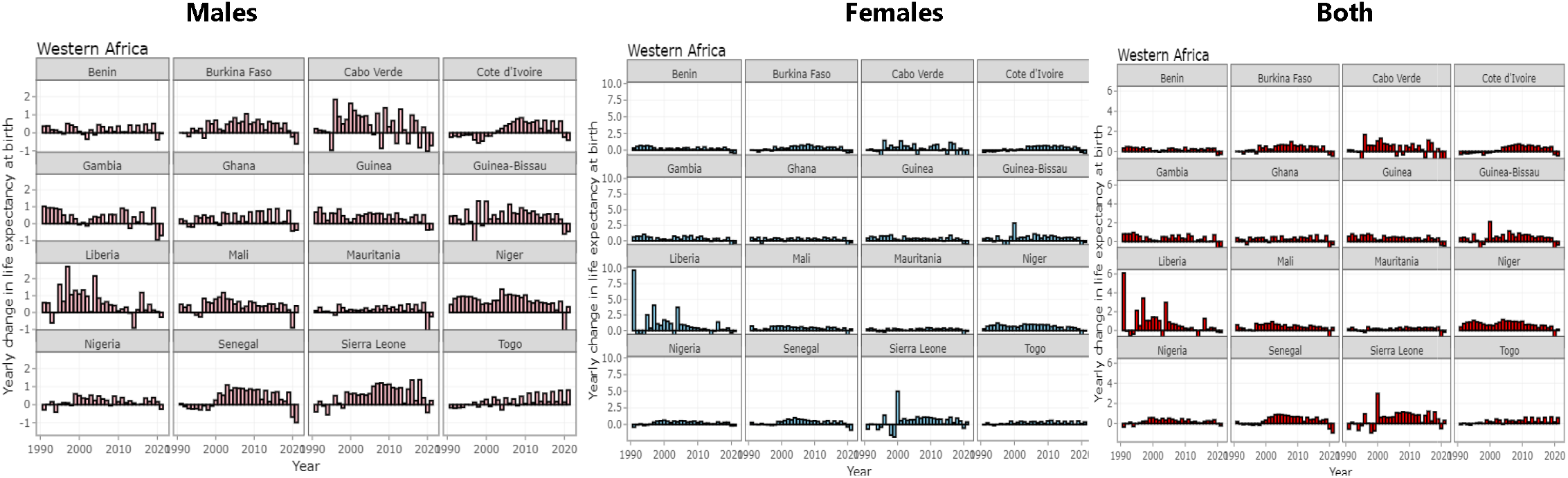
Life expectancy (LE_0_) among males, females and both sexes in West Africa (1990 to 2020) Figure 3 shows yearly changes in life expectancy at birth (LE_0_). These graphs calculated yearly changes in LE_0_ using the formula (yearly chance =LEi-LEi-1 with i = year). The graphs show that in 2020, LE_0_ decreased compared to the year before, except in Mali and Sierra Leone, where it increased in 2021. In Togo, in males and females, LE_0_ did not decrease during the COVID-19 pandemic. However, female LE_0_ increased in Niger in 2021 (0.34 compared to 2020), or for males, it increased (0.6), and the increase of LE_0_ in 2021 for females is seen.

Furthermore, the highest life expectancy gains between 1990 and 2020 in West Africa were observed in Guinea-Bissau among males (28.32 years), followed by females in Niger (20.41 years), followed by males in Mauritania (18.91 years) and females in Liberia (18.13 years). The least gains were observed in males in Mali (4.48 years) (Table 1).

In describing the trend of life expectancy among the West African population from 1990 to 2020, we found no specific pattern or gaps in LE_0_ between females and males in these graphs. For the majority, differences in LE_0_ between the two sexes were high before 2000 but decreased in 2000 and 2010. Also, LE_0_ gaps between males and females at 60 years increased in most West African countries.

On the other hand, LE_0_ showed many variations in many countries, with increases in some countries. In this study, increases in differences between LE_0_ (females)-LE_0_ (males) were calculated and presented as thus (Figure 4).

**Figure 4:**
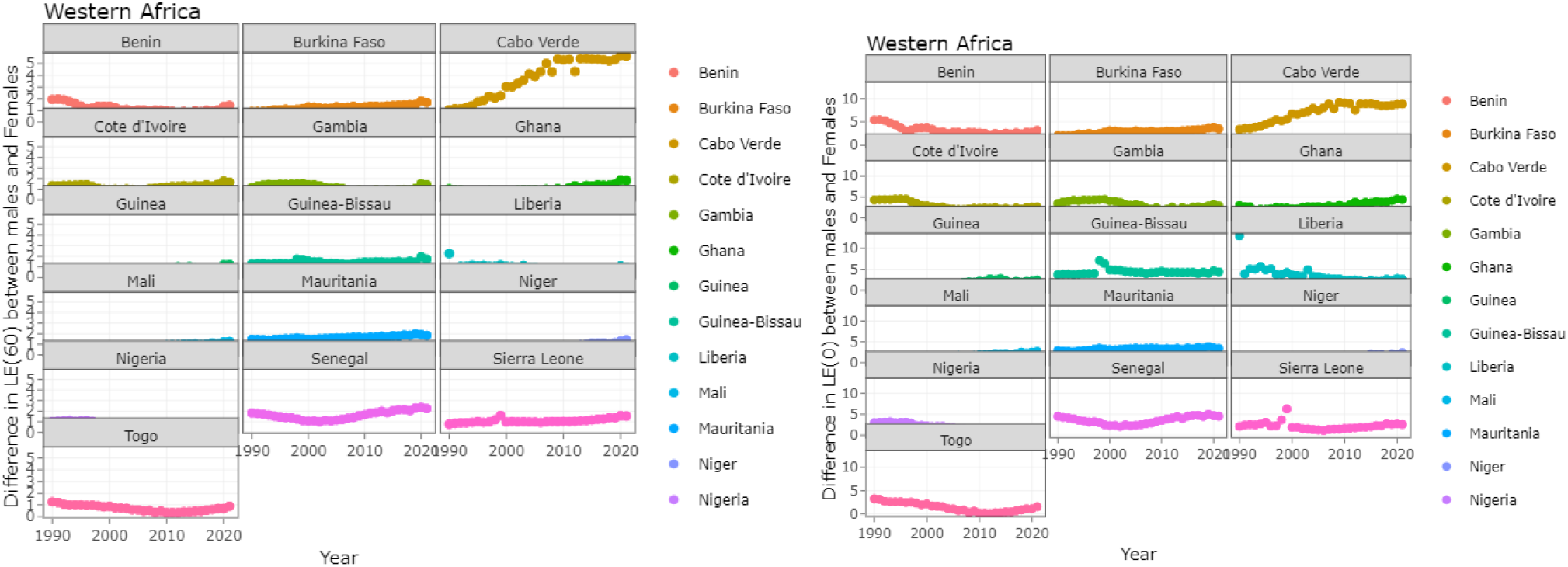
Gaps in life expectancy between males and females at 60 years and at birth in West Africa over 30 years. In Figure 4, these graphs show no specific pattern or differences in LE_0_ between females and males. For the majority, the differences in LE_0_ between sexes were high before 2000 but decreased in 2000 and 2010. Also, LE_0_ differences between males and females at 60 years increased in some West African countries. On the other hand, LE_0_ showed many variations in many countries, with increases in some countries. Increases in the difference between LE_0_ (females)-LE_0_ (males) were calculated.

In another graph from our study, all West African countries registered life expectancy losses in males and females in 2021, except among females in Togo and Mali (Figure 5).

**Figure 5:**
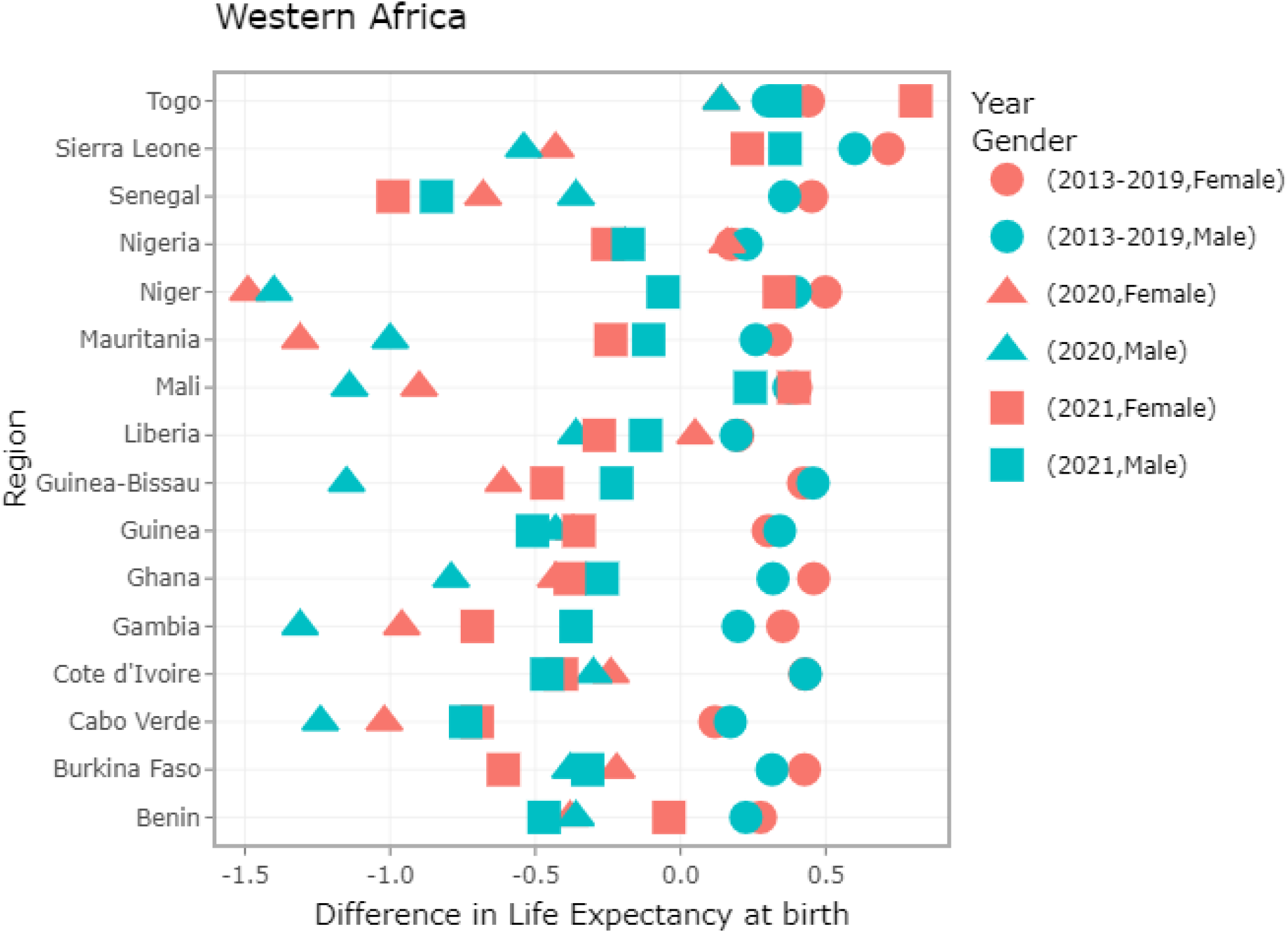
Life expectancy gaps at birth between males and females in West Africa (2013-2021). Figure 5 shows that except for females in Togo and Mali, all West African countries registered life expectancy loss among males and females in 2021.

Furthermore, Figure 6 shows that in 2021, compared to 2013-2019, LE_0_ gaps between males and females in West African countries at birth were less than zero except among females in Togo.

**Figure 6:**
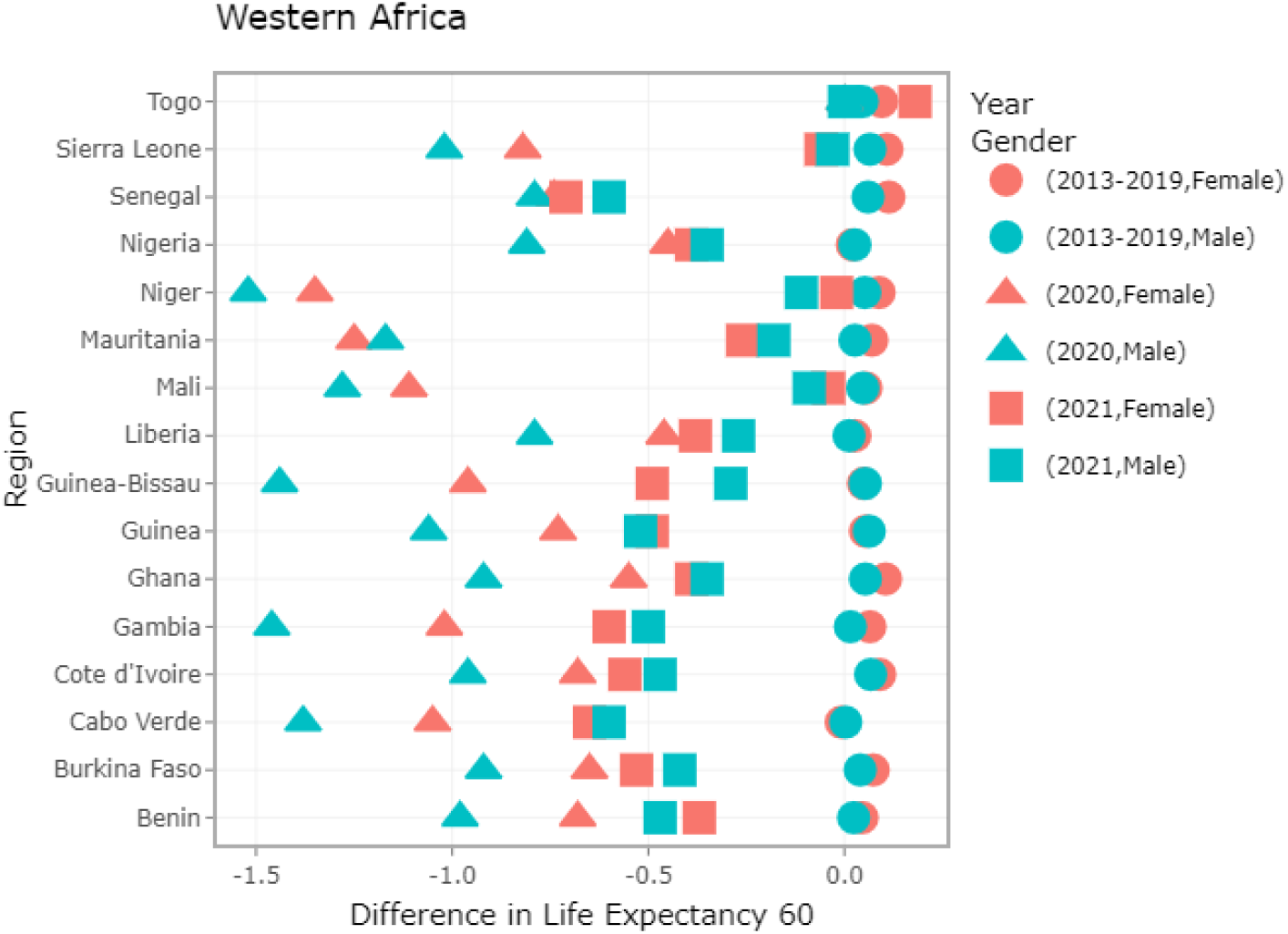
Life expectancy (LE_0_) gaps between males and females in West Africa. Figure 6 shows that in 2021, compared to 2013-2019, LE_0_ differences between males and females in West Africa at birth were less than zero except among females in Togo.

In addition, we found that males and females at 60 years in West African countries recorded losses in life expectancy in 2021 compared to 2013-2019 (Figure 7). Similarly, life expectancy at birth in this study recorded losses among males and females in 2021 compared to 2013-2019 (Figure 8). Further to the findings in Figure 8, as we studied differences in life expectancy at birth between females and males between 1990 and 2020, we noted that the highest life expectancy difference between males and females was in Liberia (at 10.19 years) and the least in Senegal (0.5 years) and Gambia (0.5 years) (Figure 9).

**Figure 7:**
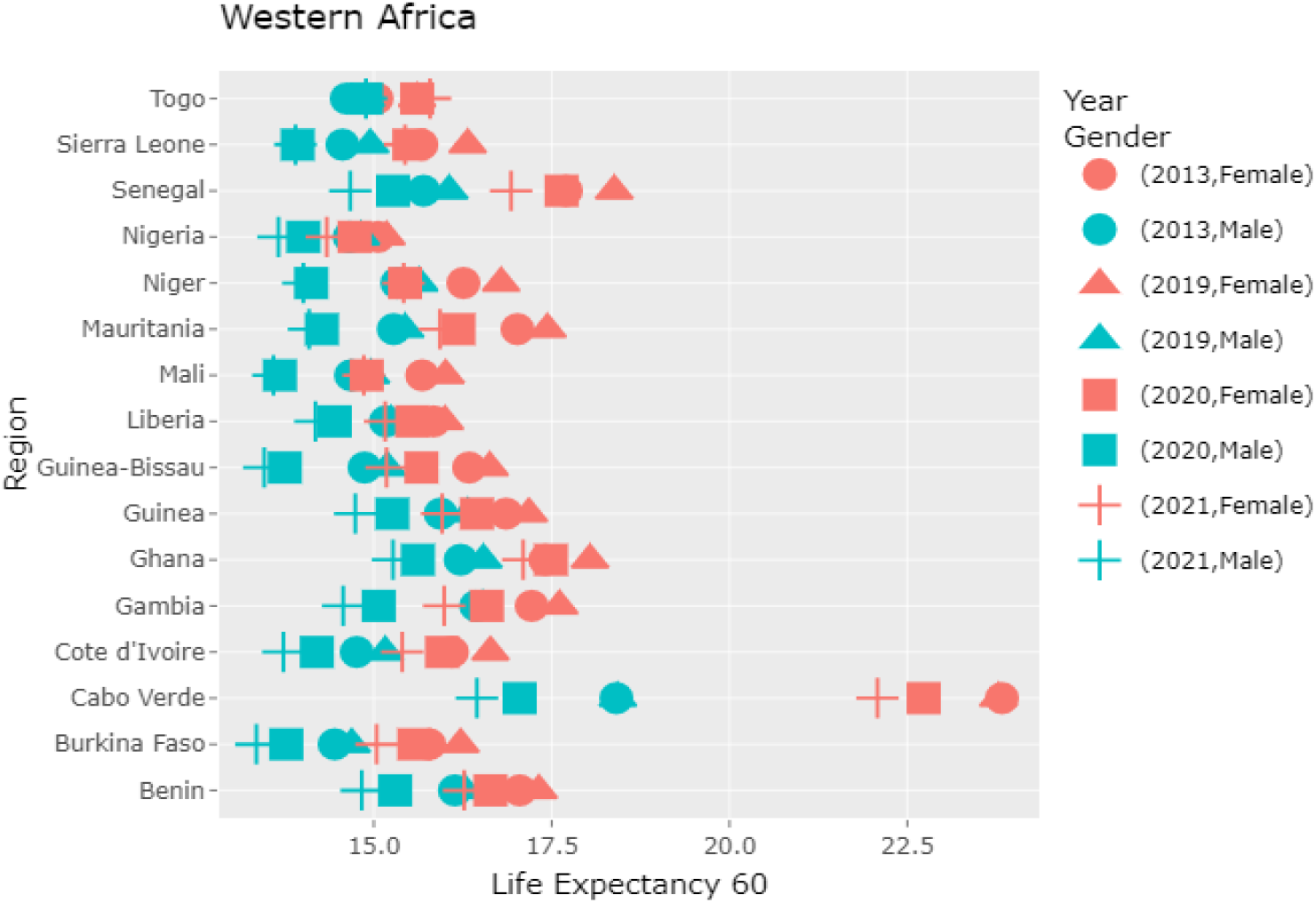
Life expectancy at 60 years between males and females in West Africa. Figure 7 shows that males and females at 60 years in West Africa recorded losses in life expectancy in 2021 compared to 2013-2019.

**Figure 8:**
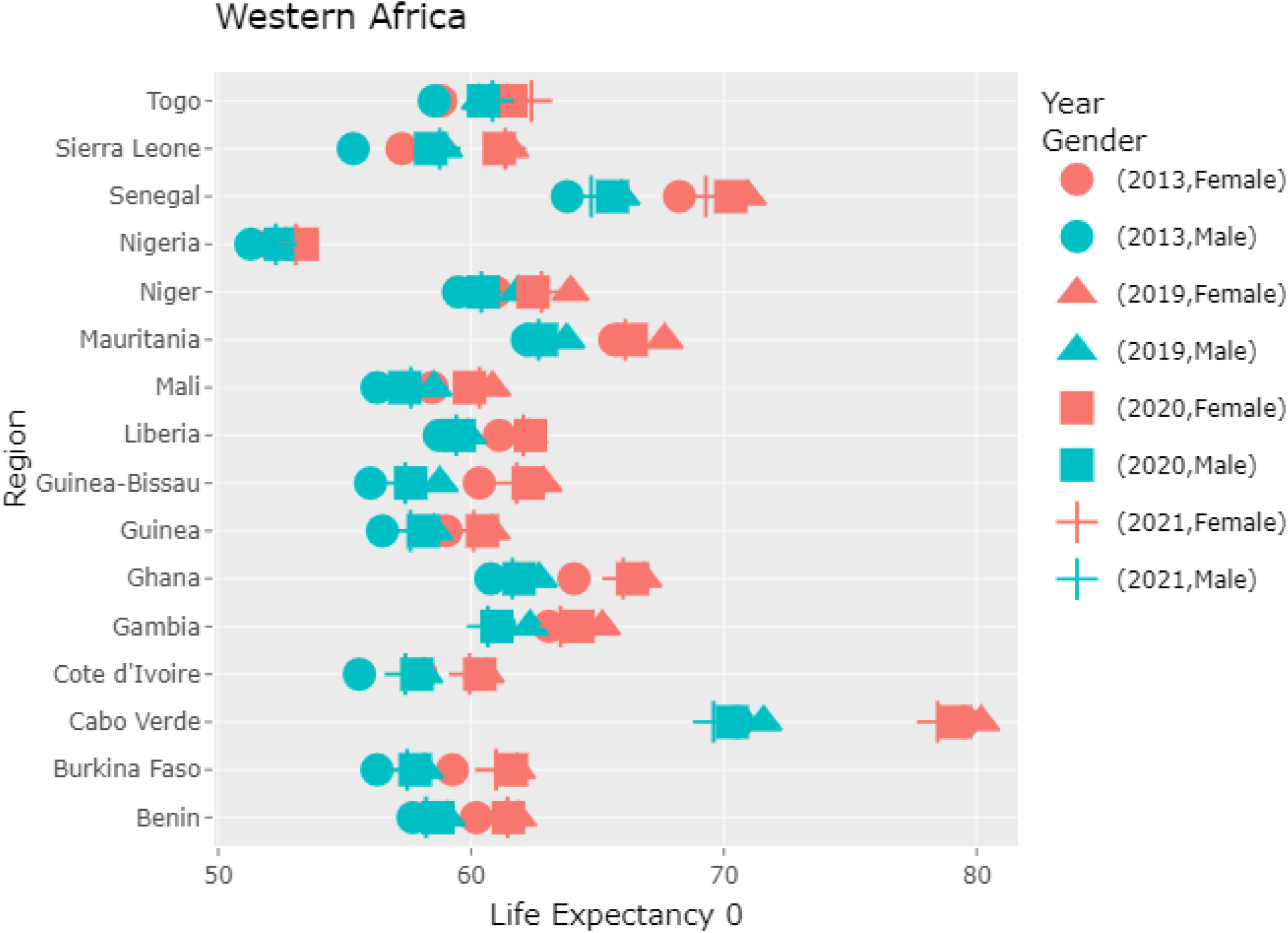
Life expectancy at birth between males and females in West Africa (2013-2021). Figure 8 shows that life expectancy at birth among males and females in West Africa recorded losses in 2021 compared to 2013-2019.

**Figure 9:**
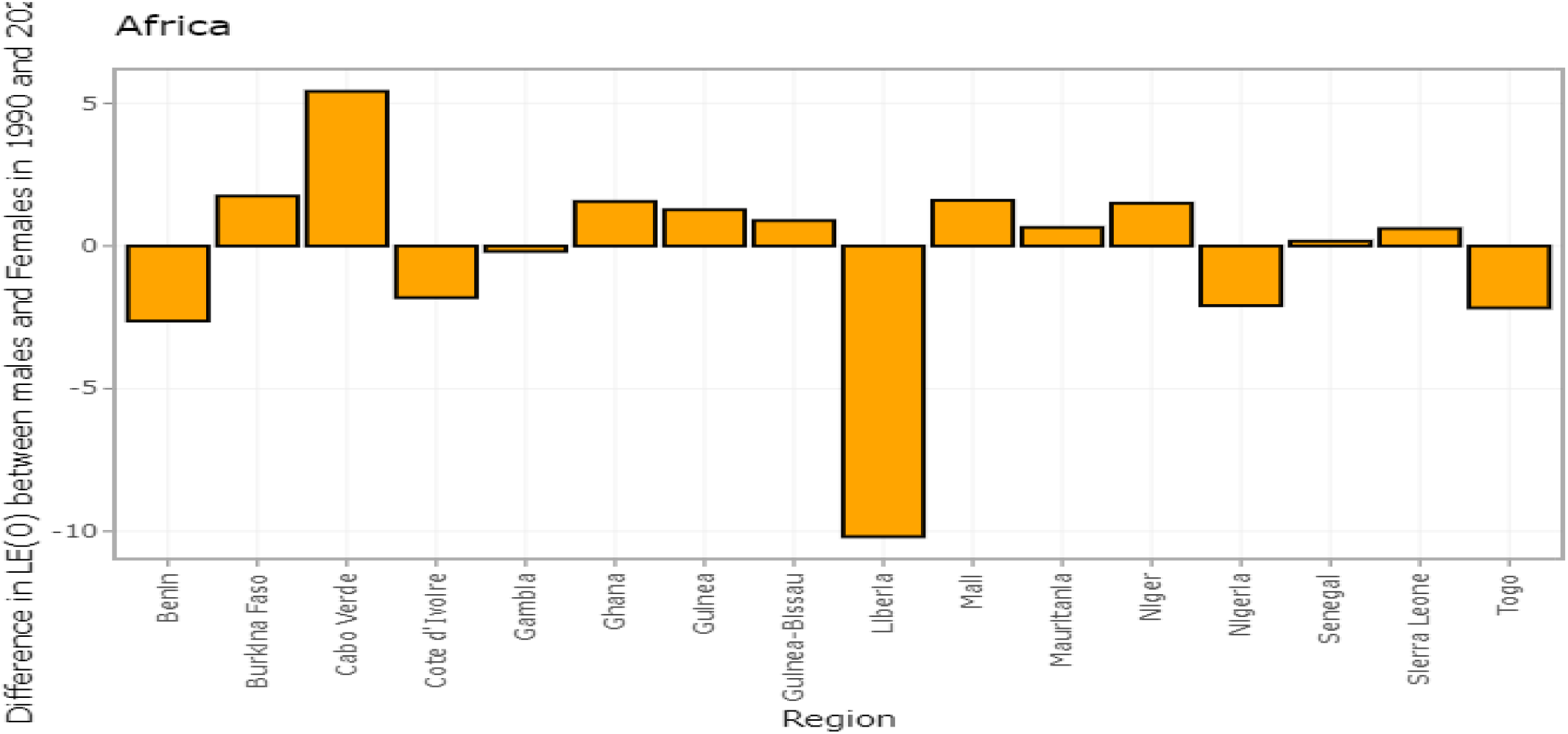
Life expectancy (0) differences between males and females in West Africa between 1990 and 2020. Figure 9 shows differences in life expectancy (LE_0_) between females and males between 1990 and 2020. The highest life expectancy difference between males and females was in Liberia (at 10.19 years) and the least in Senegal (0.5 years) and Gambia (0.5 years).

Also, we found that Liberia had the highest LE gains in the last 30 years (1990-2020) at (28.32 years) and the least in Mauritania at (3.0 years) (Figure 10). In addition, females in Niger had the highest life expectancy gains at (20.41 years) and least in Mauritania at (5 years) during the study period (1990-2020) (Figure 11).

**Figure 10:**
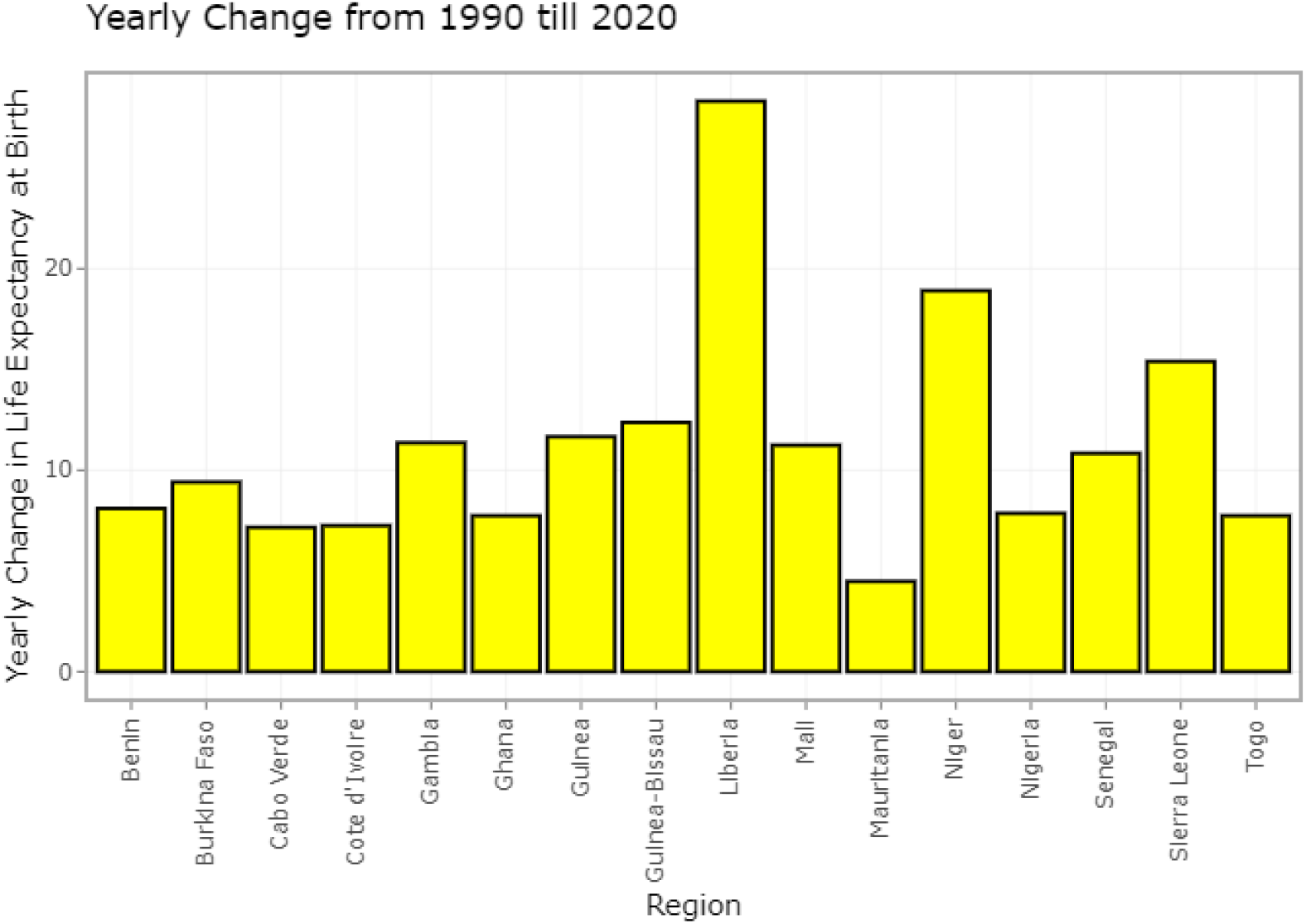
Yearly changes in Life expectancy (LE_0_) in West Africa from 1990 to 2020 among males. In Figure 10, males in Liberia had the highest yearly LE_0_ gains at birth in 30 years (28.32) and the least in Mauritania (3.0).

**Figure 11:**
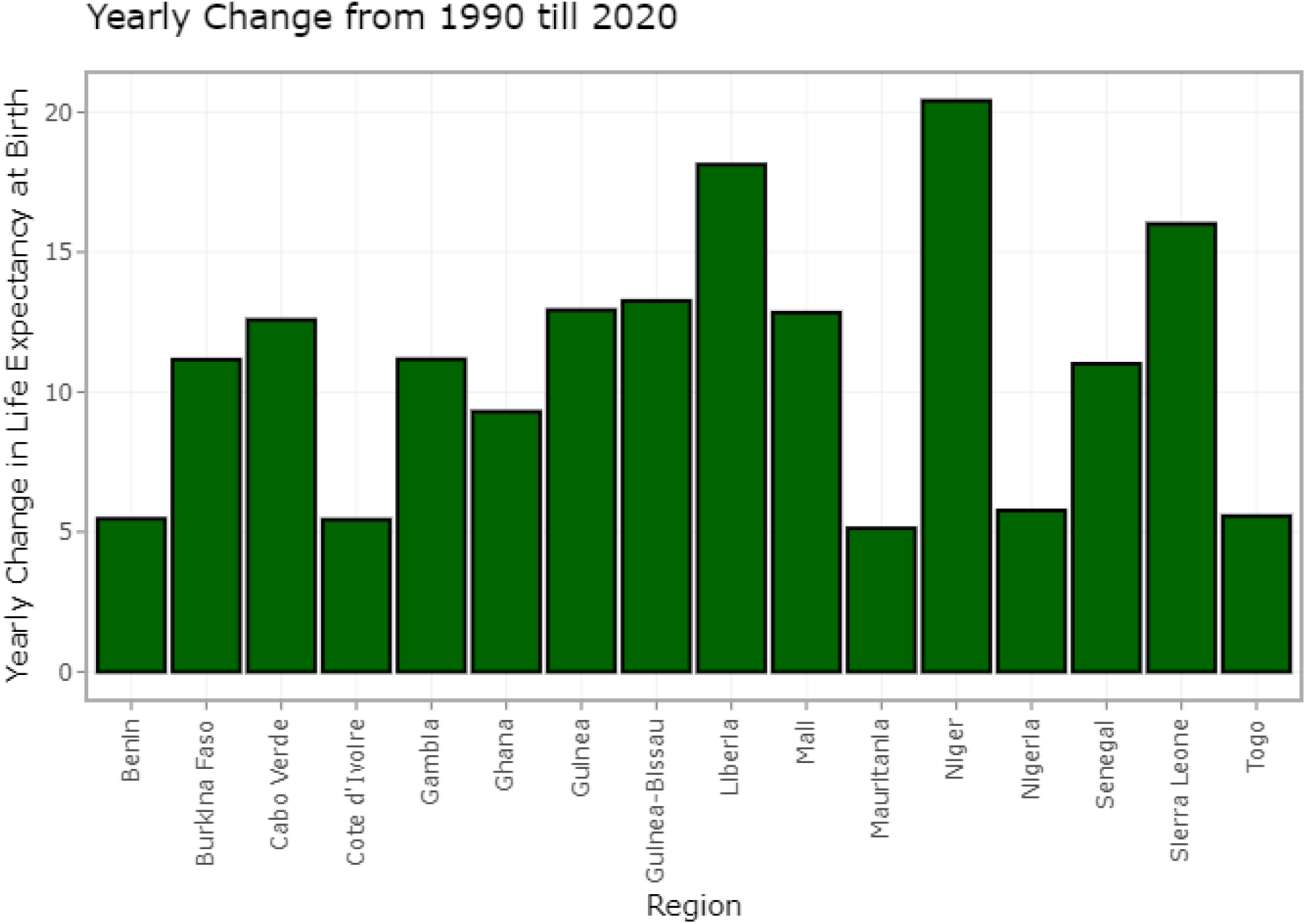
Yearly Life expectancy (LE_0_) in West Africa from 1990 to 2020 among females. In Figure 11, females in Niger had the highest yearly life expectancy (LE_0_) gains at (20.41 years) and the least in Mauritania at (5 years) during the study period (1990-2020).

In Figure 12, the yearly changes in LE between 1990 and 2020 (over 30 years) in West African countries in both sexes show gains ranging from the lowest at (4.76) in Mauritania and highest at (24.26) in Liberia. However, in males, life expectancy changes show a gradual increase over thirty years, but the pattern varied among countries (Figure 13). However, there is a gradual decline in life expectancy disparity (at 60 years) among males in West Africa (Figure 14).

**Figure 12:**
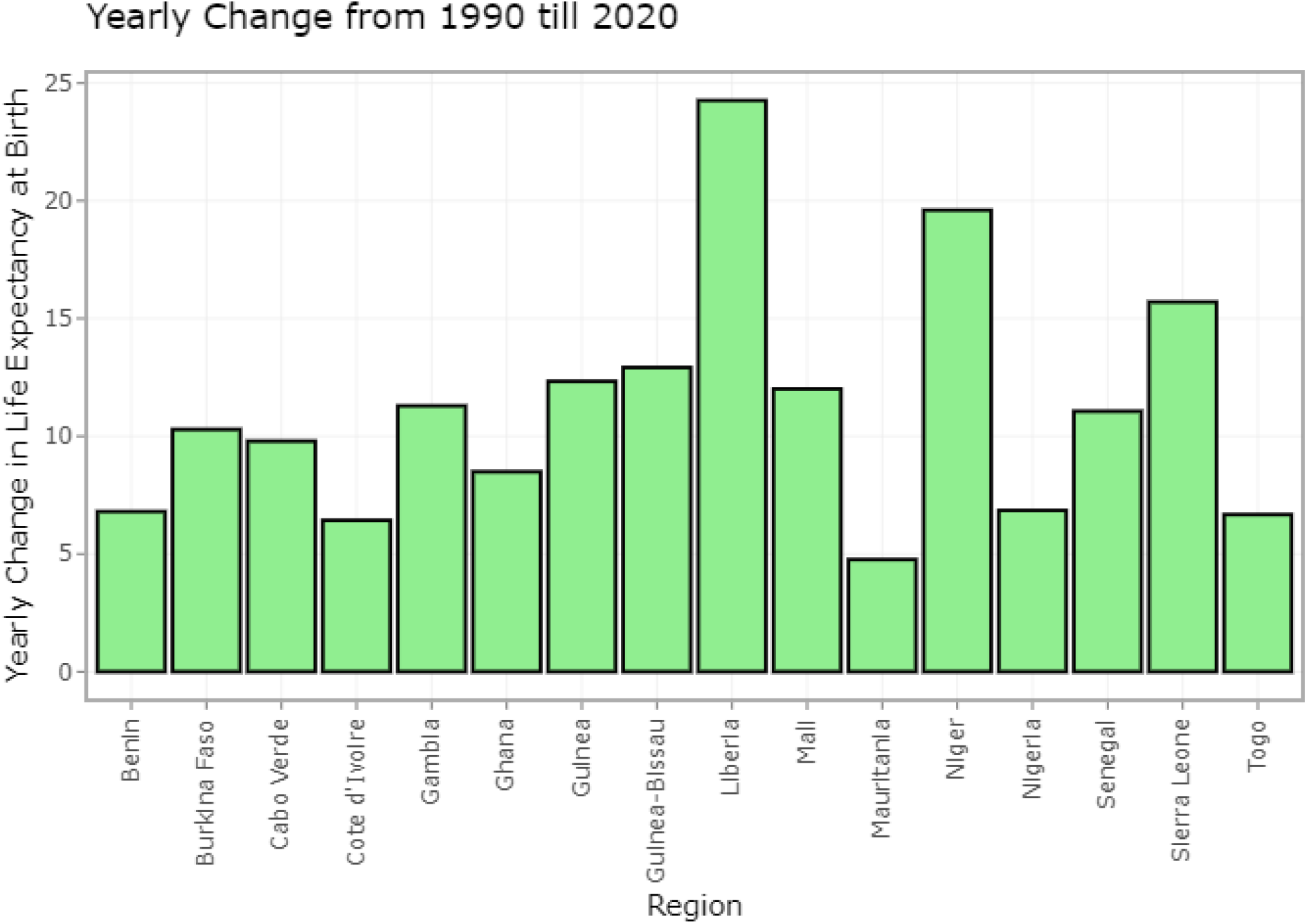
Yearly changes in Life expectancy (LE_0_) in West Africa from 1990 to 2020 in both sexes. In Figure 12, the yearly changes in LE_0_ between 1990 and 2020 (the last 30 years) in West Africa in both sexes show gains ranging from the lowest at 4.76(in Mauritania) to the highest at 24.26(in Liberia).

**Figure 13:**
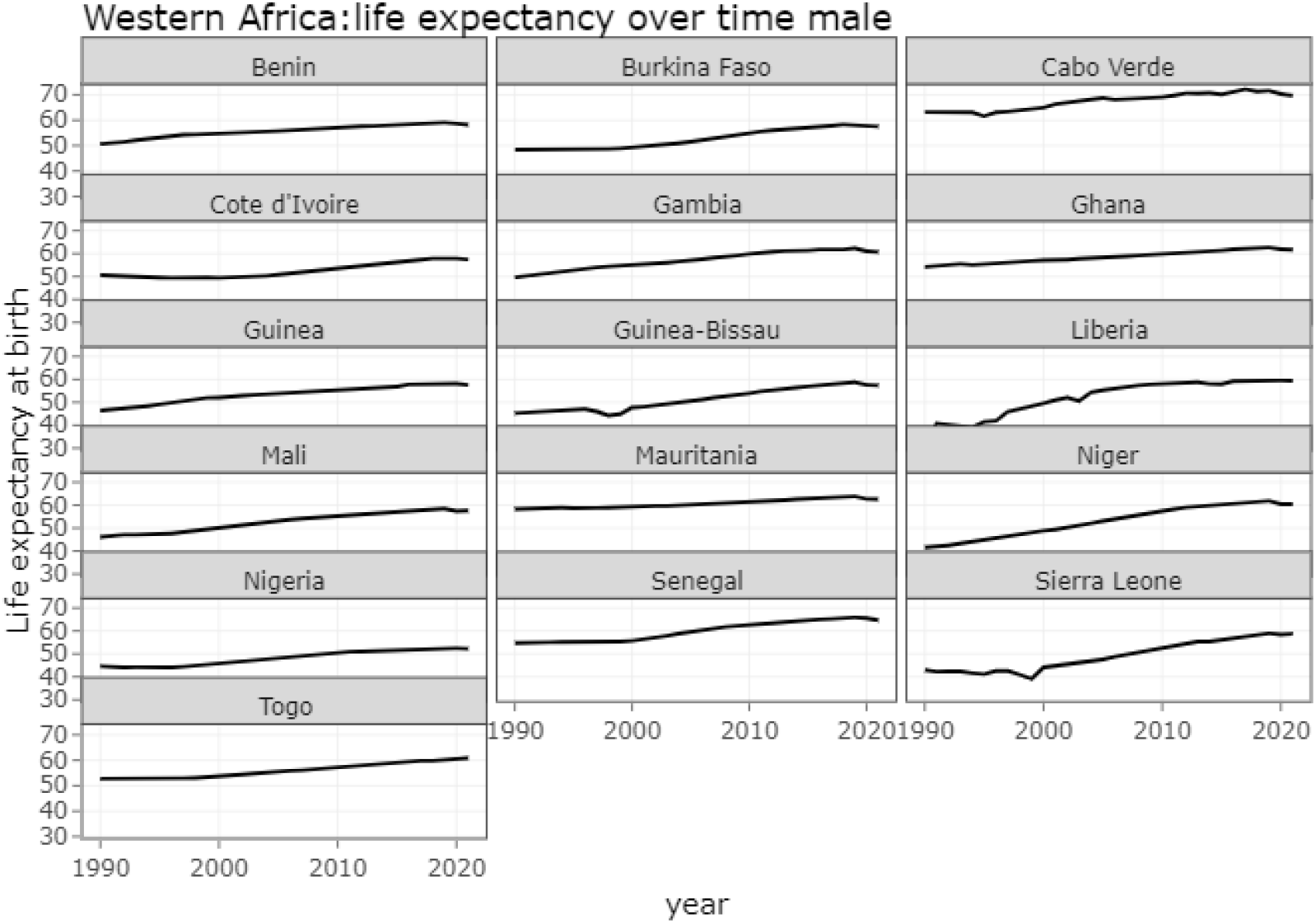
Life expectancy (LE_0_) overtime in West African males over 31 years. Figure 13 shows life expectancy (LE_0_) changes among males in West Africa. It shows a gradual increase over the 31 years, but the pattern varied among countries.

**Figure 14:**
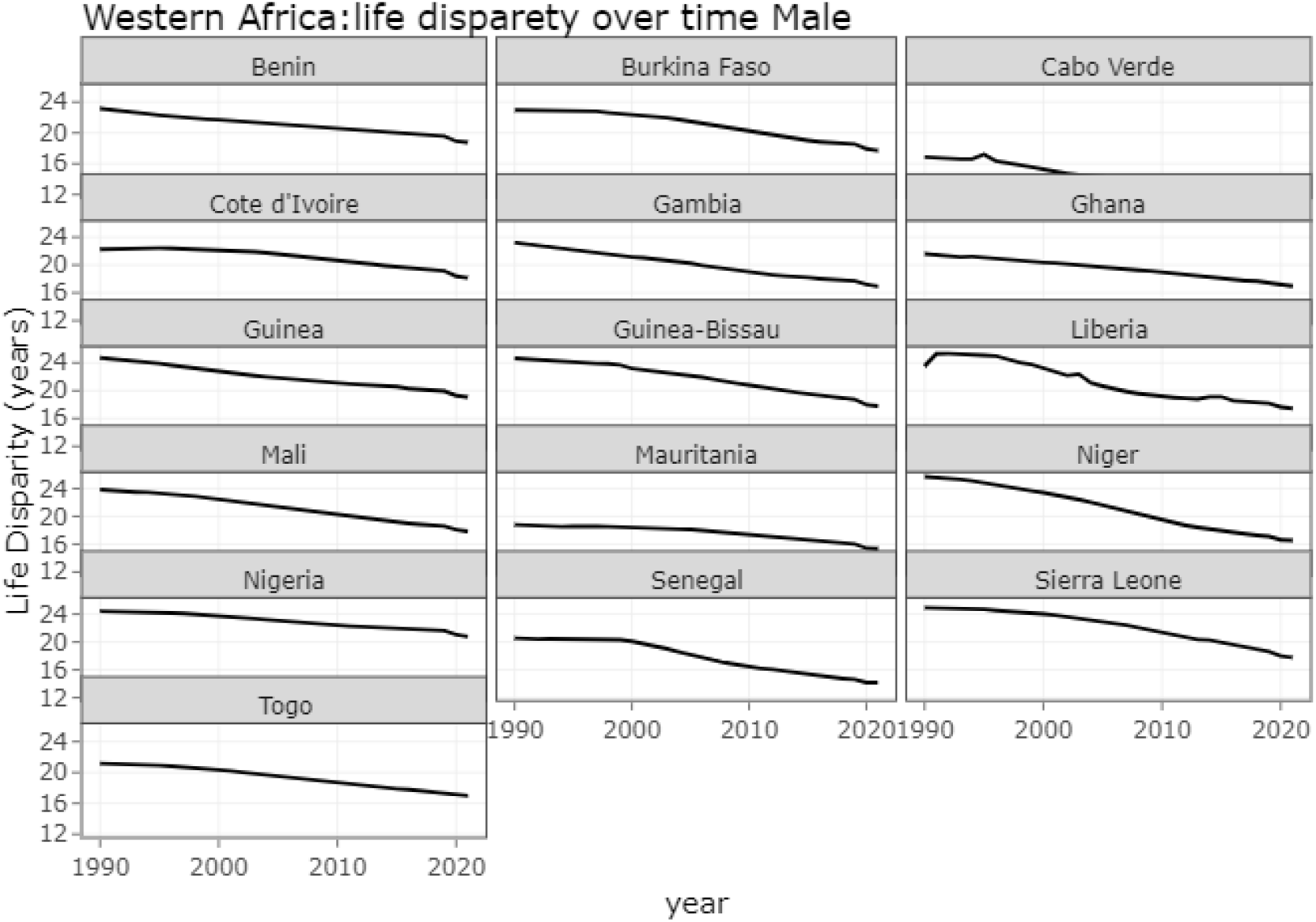
Life disparity (at 60 years) in West Africa among males (1990-2021). Figure 14 shows a gradual decline in life disparity (at 60) among males in West Africa.

On the other hand, there is a gradual increase in life expectancy at birth among the female population in West Africa (Figure 15). In contrast, Figure 16 shows a gradual decline in life expectancy (at 60 years) among the female population in West Africa over the last thirty years (1990-2020).

**Figure 15:**
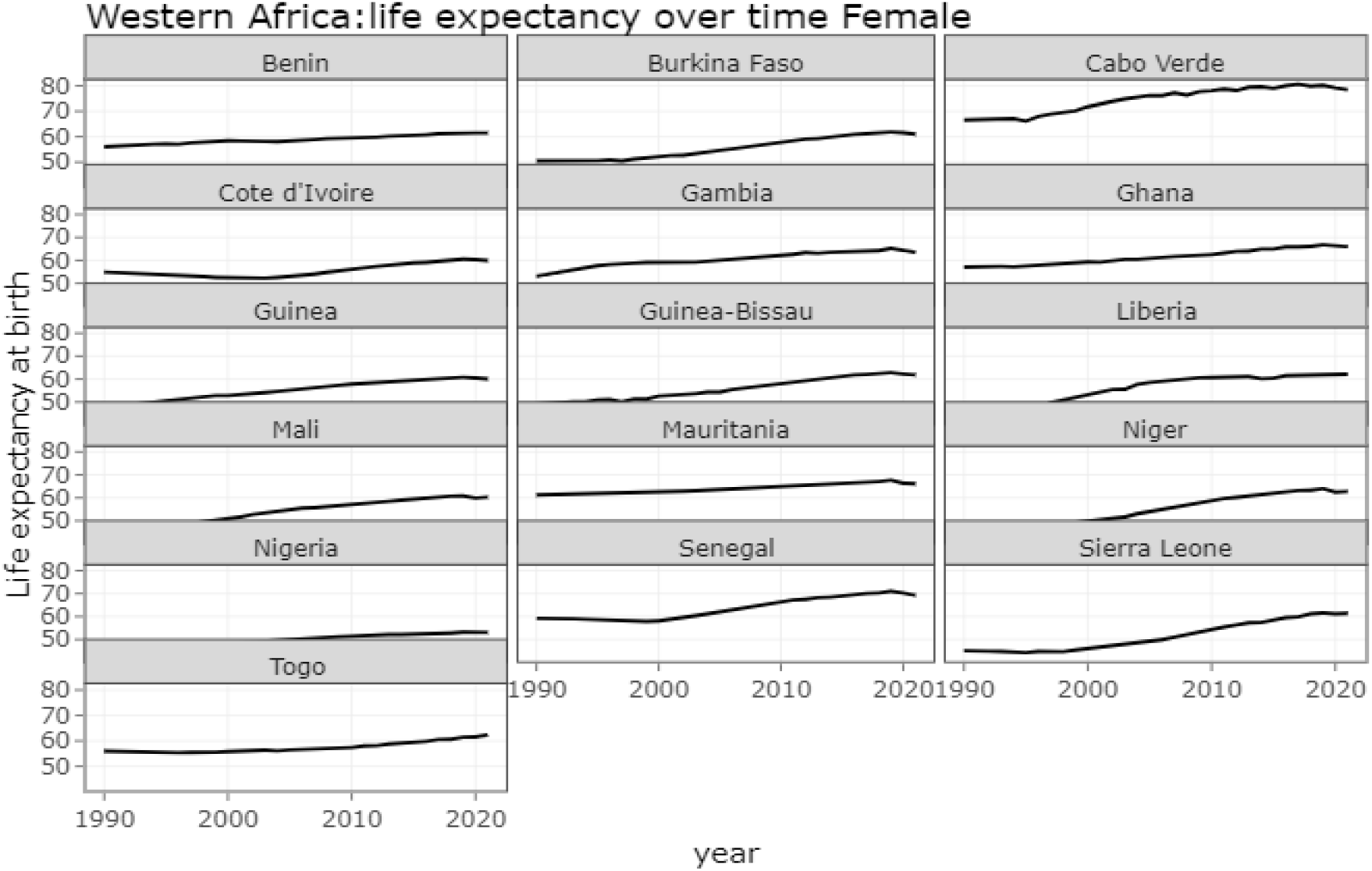
Life expectancy (LE_0_) overtime in West Africa among females (1990-2021). Figure 15 shows gradual increases in life expectancy at birth among female population in West Africa.

**Figure 16:**
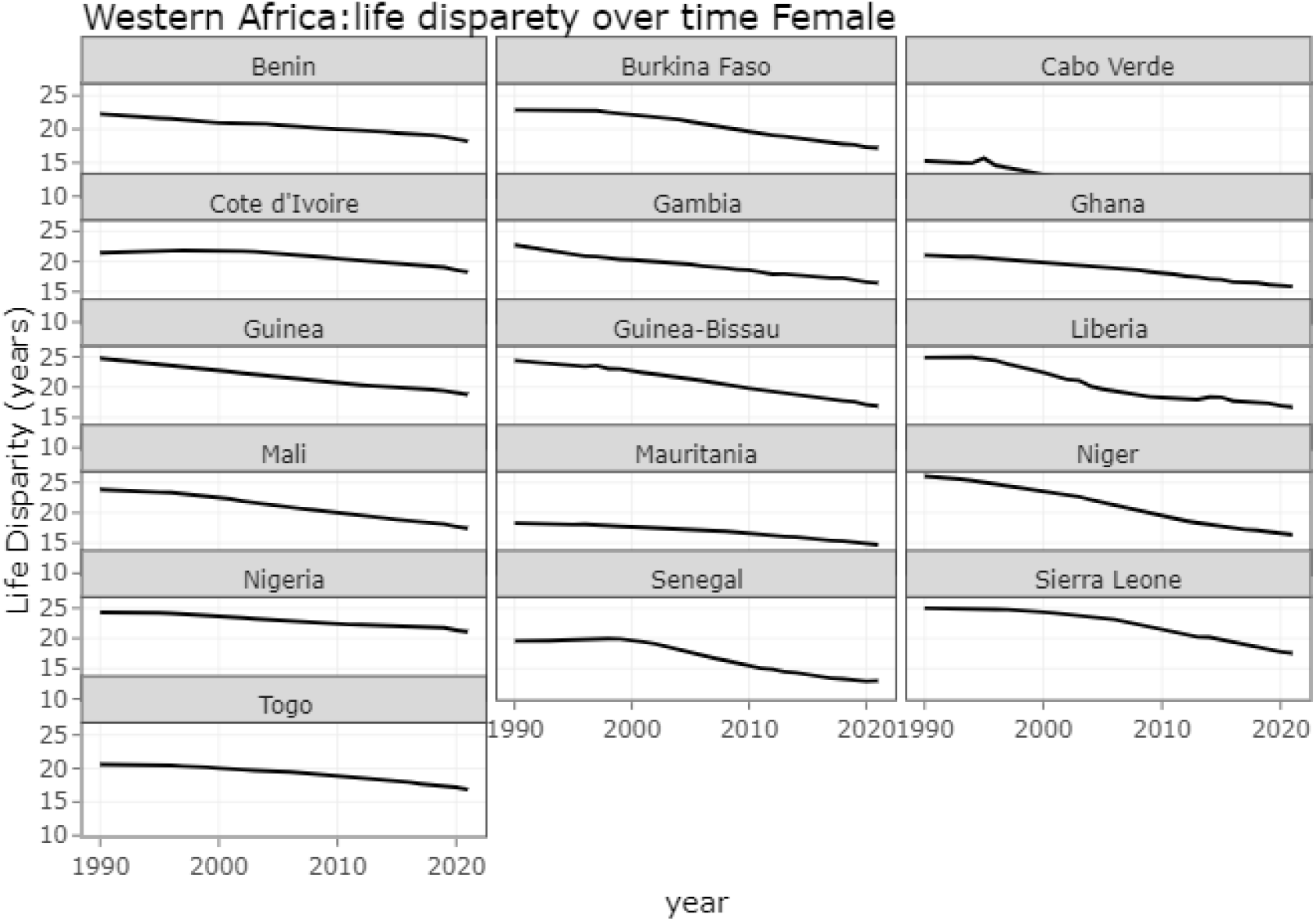
Life disparity (at 60) among females in West Africa (1990-2021). Figure 16 shows a gradual decline in life disparity (at 60 years) among female population in West Africa over the last thirty-one years (1990-2021).

In studying the life expectancy in West Africa in detail, the mortality survivorship curve was constructed. It showed a changing pattern and contributions of deaths at different ages to life disparity contribution in West African countries over 30 years. These graphs show that the number of survivors in West African countries has increased over the last 30 years (1990 to 2020) (Figure 17).

**Figure 17:**
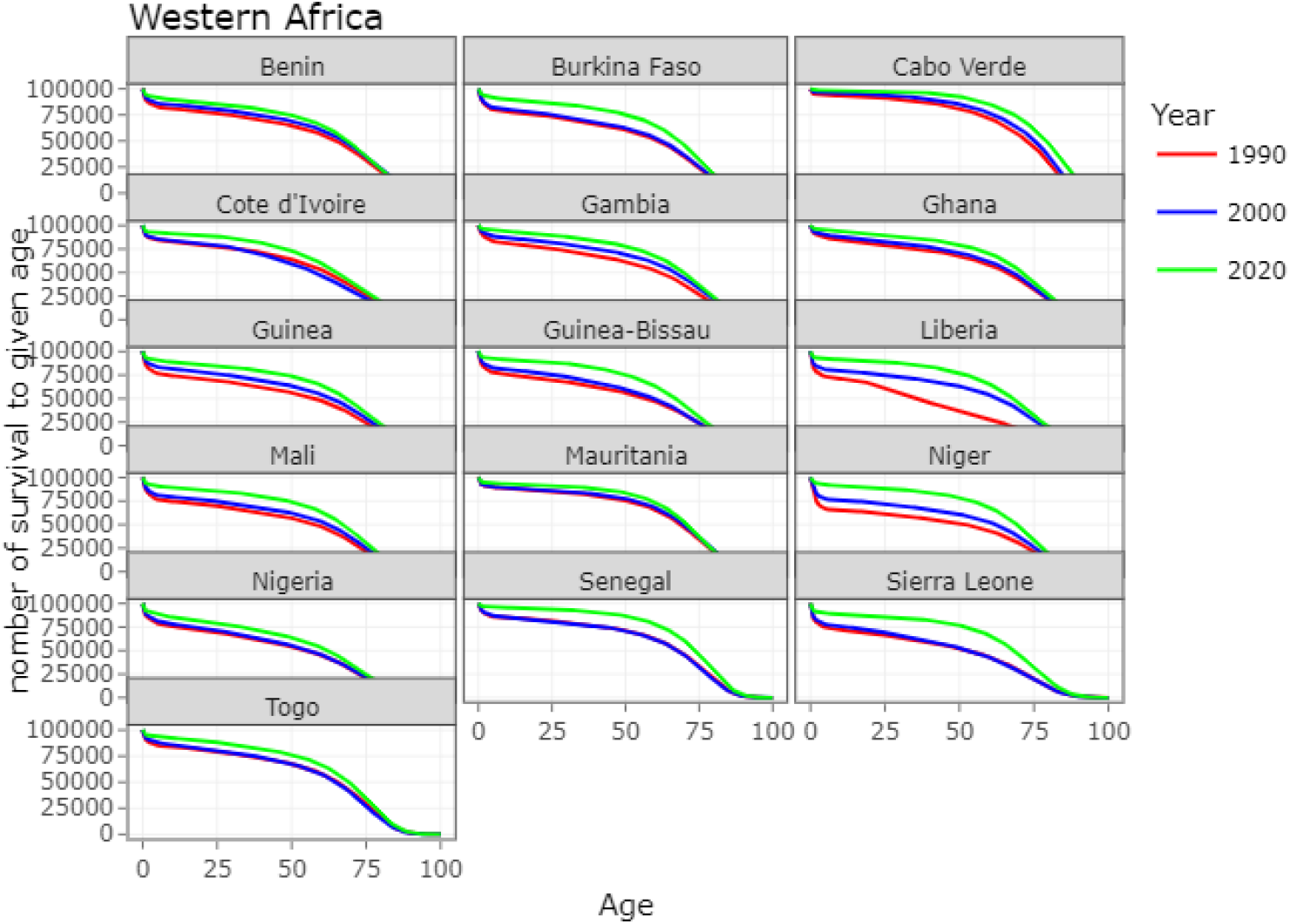
Survivorship curve in West Africa over 30 years (1990-2020). Figure 17 shows the changing mortality survivorship curve and contributions of deaths at different ages to life disparity contribution in West African countries over 30 years. These graphs show that the number of survivors at a given age in West African countries have improved over 30 years (1990 to 2020) but decreases with increasing age of the population.

Figure 18 shows improving life expectancy disparity across all West African countries over the 30 years (1990-2020). Nevertheless, most life disparity was observed among children under ten years.

**Figure 18:**
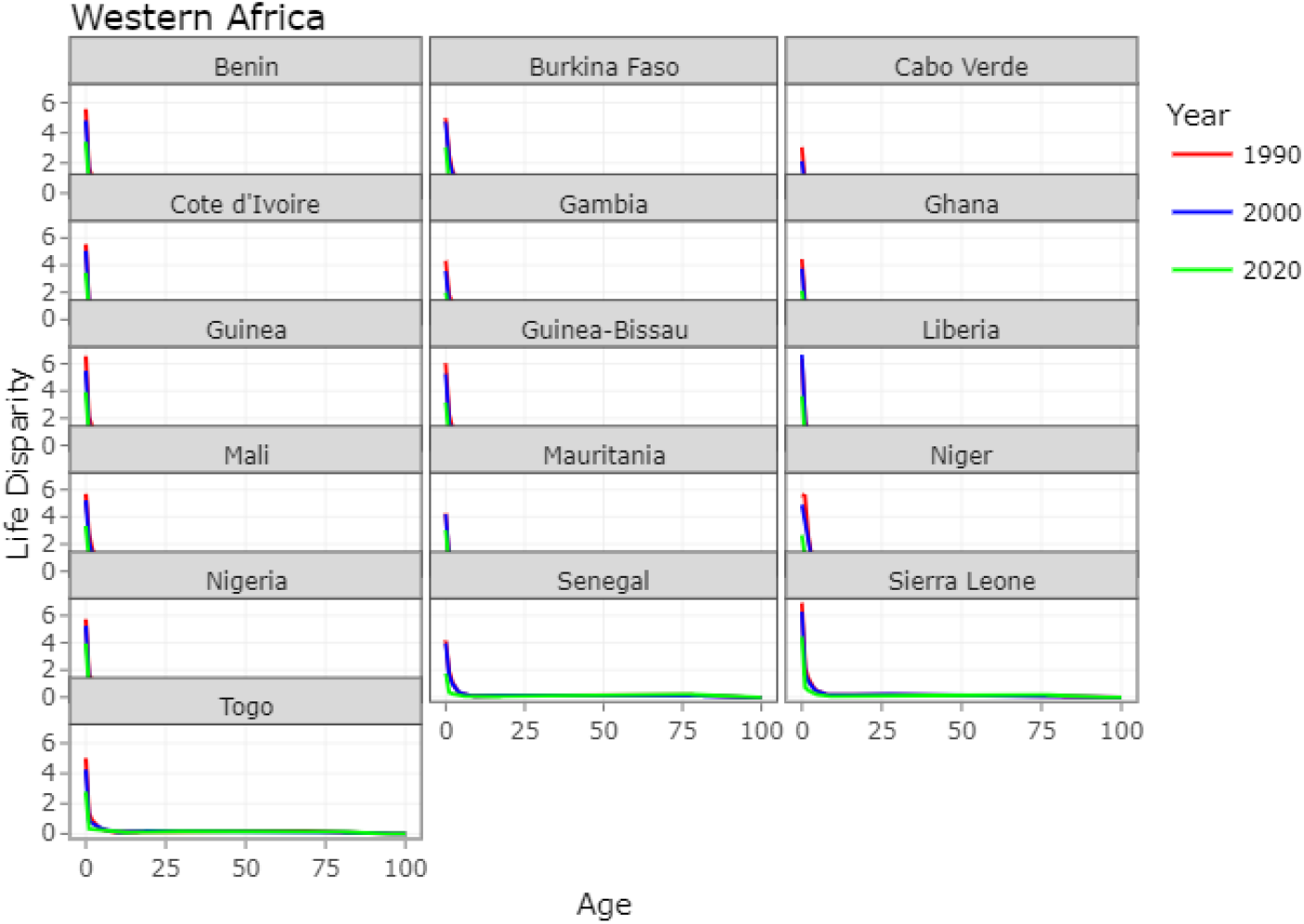
Life disparity among age groups in West Africa over 30 years (1990-2020). Figure 18 shows improving life disparity across all West African countries over 30 years. However, most life disparity was observed among children under ten years.

Figure 19 is a Gini coefficient of life expectancy in West Africa over 30 years (1990-2020), showing declining life inequality in all West African countries. The life inequality years between people within the same country were measured with the Gini coefficient. A high Gini coefficient meant a significant within-country inequalities in the number of years people live. Of note, life disparity and the Gini coefficient are the other life span measures used other than life expectancy (Figure 19).

**Figure 19:**
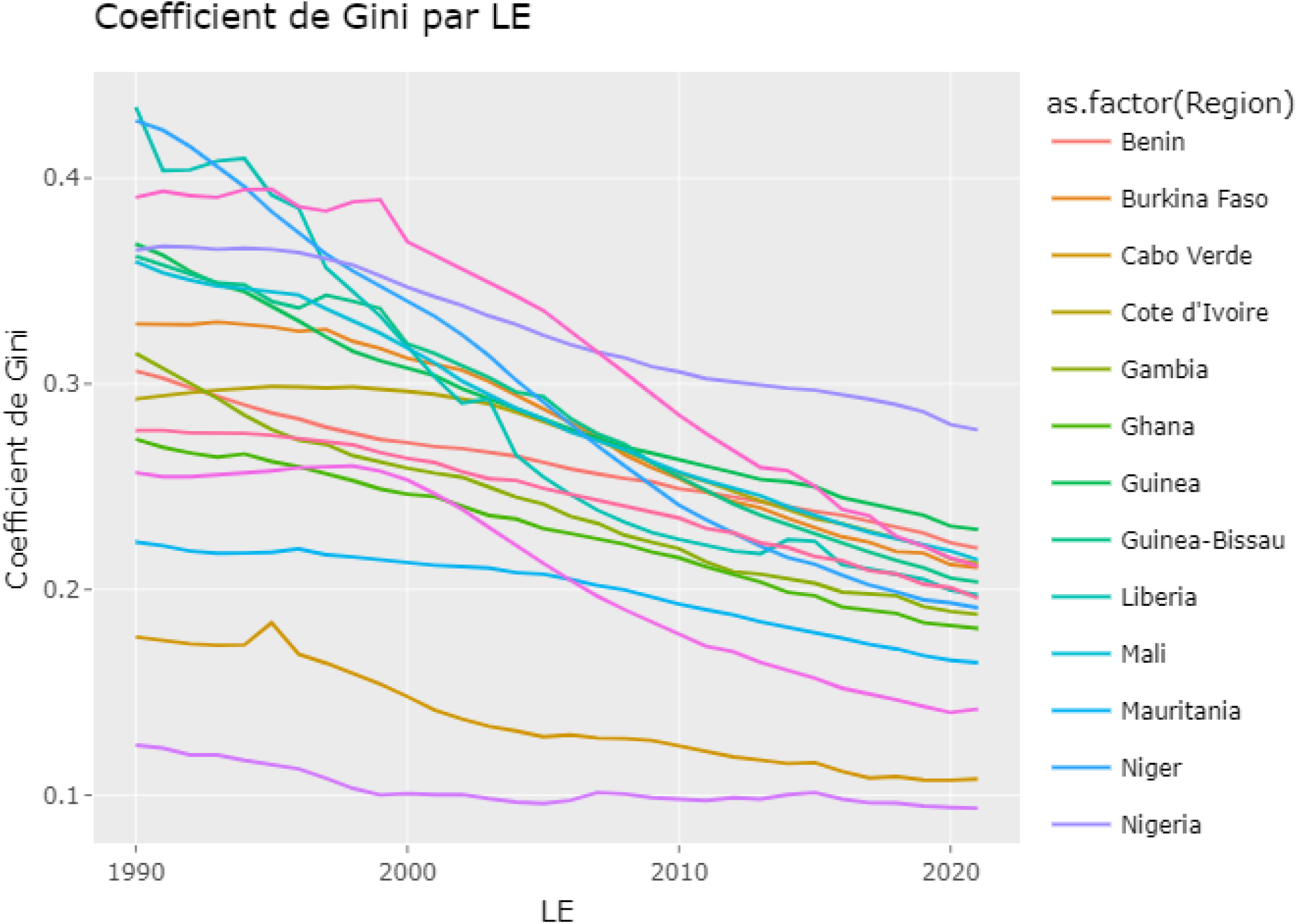
The Gini coefficient in West Africa over 31 years (1990-2021). Figure 19 is a Gini coefficient of West African countries over 31 years (1990-2021), showing improvement of the coefficient over the years among West African countries.

Life-table construction for all sixteen West African countries by sex for 1990–2021 was done following a piece-wise constant hazard model using all-cause mortality by single age, with the last age interval grouping deaths at ages 81-100 years, consistent with standard demographic techniques.^26^ Our choice of grouping deaths aged 81-100 years is common practice in demographic research, and it ensures comparability across countries, primarily because some countries used this last age group to report deaths for 2020. The choice of the last open-ended interval may impact life expectancy estimates depending on the proportion of the population surviving this last interval.^27^

From these life tables, life expectancy at birth and life expectancy conditional on surviving to age 60 years for males and females were extracted for each country by sex (Figures 4, Figure 7, Figure 14, Figure 16, Figure 17). We performed several sensitivity checks to ensure the robustness of our estimates, including comparisons with alternative sources such as the United Nations (UN) and the Human Mortality Database (HMD) for the period from 1990 to 2020 or their most recent year available, 2021.

## Discussion

Our study found substantial gains in life expectancy (LE_0_) among males and females in West African countries over the three decades (1990-2020) (Figure 1, Figure 2, Table 1). However, life expectancy gains were variable among W. African countries, but the highest gains were in Guinea-Bissau (28.32 years) and least in Mali (4.48 years). Females in Cape Verde had the highest life expectancy in 1990 (66.58 years) and (79.16 years) in 2020 among all West African countries (Table 1). With the emergence of the COVID-19 pandemic in 2020-2021, most West African countries experienced life expectancy losses, except Togo (Figure 3, Figure 4, Figure 5, Figure 6, Figure 7, Figure 8).

As already observed by many studies, in the absence of wars, new epidemics, or substantial economic reforms, lack of improvement or stagnation in life expectancy (LE_0_) gains is considered a cause for concern.^28^ Actual declines in life expectancy (LE_0_) are particularly alarming.^28^ In addition, stagnation or declines in life expectancy may signal a decline in the health profile of the population driven by adverse socioeconomic trends, a deterioration in the provision or quality of healthcare services, or worsening behavioral factors.^28^ A general pattern that emerges is that the more significant the decline in life expectancy at birth (LE_0_), the greater the role played by respiratory and cardiovascular diseases.^28^

Experts, specialists, and public health practitioners assert that life expectancy is a critical summary measure of the health and wellbeing of a population.^29,30^ A nation’s life expectancy reflects its social and economic conditions and the quality of its public health and healthcare infrastructures, among other factors.^29,30^ Monumental improvements in life expectancy (LE_0_) have been the predominant trend for high-income, developed countries over the 20^th^ and 21^st^ centuries.^29,30^

Of particular interest in the life expectancy study in West Africa were (LE_0_) gaps between males and females was negative in Benin, Cote d’Ivoire, Gambia, Liberia, Nigeria, and Togo from 1990-2020, an indication of the consistently higher life expectancy among females compared to males (Figure 9). In addition, the yearly changes in life expectancy over the 30 years among males were highest in Liberia and least in Mauritania (Figure 10).

Similar findings were observed among females, with the highest total yearly LE_0_ gains in Niger (21 years) and the least in Mauritania (3.0 years) (Figure 11). In the combined sexes, total yearly LE_0_ changes were highest in Liberia (24 years) and least in Mauritania (3.0 years) (Figure 12).

Several propositions have been advanced about the determinants of life expectancy. Shin et al. proposed that the impact of the pension system could make life expectancy longer or shorter, and it is only sometimes valid that the pension system improves the lifetime utility level.^31^ While Hazan (2012) proposes a positive correlation between the percentage change in schooling and the change in life expectancy.^32^ The life expectancy gains in West Africa were partly due to the introduction of universal primary and secondary education in the 2000s, as proposed by these authors.^31,32^

Further in our findings, LE_0_ over time in West Africa among males and females showed progressive gains over thirty years (Figure 13, Figure 15). In addition, life disparity among males and females also reported a progressive decrease, indicating a decrease in inequalities among the population in the region (Figure 14, Figure 16). These findings are consistent with studies that showed that the determinants of life expectancy (LE_0_) gains can be divided into economic, social, and environmental factors. These factors include wages, the number of beds in hospitals, the number of doctors, and the number of readers subscribed to libraries^33^, nutrition and food availability factors^34^, household incomes, nutritional intake, literacy, number of physicians in a country^35^, environmental quality dynamics^36^, per capita income, health expenditures, literacy rate and daily calorie intake^37,38^, human capital formation and life expectancy factors potentially reinforced each other due to advances in technological progress.^39,40^

Furthermore, our study found a survivorship curve showing improvement in life expectancy at birth among the West African population (Figure 17), consistent with the improving life disparity across all W. African countries in the 30 years (Figure 18). In addition, the Gini coefficient showed declining inequalities among the population of West Africa over the 30 years (1990-2020). All these findings are consistent with previous studies on life expectancy gains in the African continent over the period.^9,14,16,21,22,23^

This information that West African countries have been registering life expectancy gains is well-come information as an extension of life expectancy (LE_0_) has always been a primary interest of medical research and an indicator of national public health profiles.^41^

Globally, life expectancy has exhibited patterns of continuous growth over time, but it has also demonstrated persistently high variability between countries over the past half-century.^42,43^ Changes in life expectancy can result from long-term changes in many factors, including political regimes and socioeconomic status.^44,45^ Political regimes have been used as a distal determinant of life expectancy at country level,^46,47^ and it has consistently been associated with a negative impact on life expectancy; thus, a country with political instability displays life expectancy losses.^48,49^ Thus, investment in welfare and health policies, such as ensuring safe childbirth for mothers and babies, securing children’s right to nutrition, enhancing the education of women and children, and increasing accessibility of public health and medical services, could benefit population health by redistributing resources to more people who are in need.^47,50,51^ Many factors affect life expectancy of a population, for example, hygiene/sanitation status, health care systems, industrialization, technological progress, literacy rates, natural and manmade disasters, global changes, and HIV and AIDS pandemics.^52,53^

### COVID-19 and life expectancy (LE_0_) losses in West Africa

Since the COVID-19 outbreak at the end of 2019, the pandemic has evolved into the most significant public health crisis of the new millennium.^28^ As of August 2nd, 2023, there were 769 million confirmed COVID-19 cases, with 6.95 million deaths resulting from infection with the SARS-CoV-2 around the world.^12^ This estimate, although astounding, disguises the uneven impact of the pandemic across different countries and demographic characteristics like age and sex^13^, as well as its impact on population health, years of life lost^14^, and longevity.^1^

Globally, the outbreak of the COVID-19 pandemic has caused a profound mortality increase.^1-7^ Many scientists, experts, and researchers believe there is a need to comprehensively understand COVID-19’s effects on mortality to assess better the pandemic’s consequences on public and population health.^54,55,56,57^

This study finding provides a new assessment of COVID-19’s effects on mortality in sixteen West African countries, accounting for the year-on-year intrinsic variations of mortality, which have been largely overlooked in previous research. The results indicate that after considering the expected intrinsic changes to life expectancy between 2013-2019, 2020, and 2021, losses in life expectancy caused by COVID-19 amounted to about 1.0 year at age 60 years and below and less than one year at birth in all sixteen countries (Figure 5, Figure 6, Figure 7, Figure 8). These results suggest that the COVID-19 pandemic resulted in a considerable increase in mortality in 2020 and 2021 in Africa, even though previous studies had suggested that the effect of COVID-19 would be relatively lower because of the young demographic characteristics, warm climate, and less crowding of the population due to the vast geographical environment.^58-66^ Life expectancy losses during the COVID-19 pandemic occurred in (15/16, 93.8%) West African countries, except Togo. It was noted that the effects of COVID-19 on life expectancy in West African countries were more than initially expected when the year-on-year intrinsic variations of life expectancy were considered (Additional files (Tables 1-34).

This study captures better, life expectancy losses caused by COVID-19 in West African countries. It corrects previously underestimated effects of COVID-19 on life expectancy in Africa (Figure 5, Figure 6, Figure 7, Figure 8), Additional files (Tables 1-35). This similar occurrence has been observed elsewhere during the COVID-19 pandemic. For example, the loss of life expectancy at birth caused by COVID-19 in the USA was previously reported as ranging from 1.18 to 1.87 years without considering the intrinsic year-on-year mortality variations.^67,68,69,70^

### Life expectancy (LE_0_) in Nigeria

Our study found that the life expectancy of Nigerians (the most populous country in West Africa) in the period 2020-2021 compared to 2019 was marked with losses, with -0.40(CI:-0.94-0.13);p=0.139 in 2020 and -0.75(−1.29--0.22);p=0.006 in 2021 (Additional files Table 26). Many studies show that the coronavirus slashed living conditions by two years and reversed the trend of people living longer than their parents and grandparents.^68,69^ There have been reports worldwide that the Coronavirus disease (COVID-19) has led to a drop in life expectancy.^1-7,71^ Several studies suggest that the pandemic has led to the neglect of other diseases and, consequently, a rise in mortality and morbidity.^71^

According to studies, although lifespan has changed very little, if at all, global life expectancy has soared by more than 40 years since the beginning of the 20th century.^71^ This was achieved through scientific discoveries and public health measures that drove down infant mortality rates.^71^ However, medical experts are unanimous that the coronavirus pandemic is causing a substantial increase in mortality in Nigeria and worldwide populations because the pandemic has overwhelmed health systems, increased morbidity, mortality, and the increase in mortality has the potential of causing a decline in life expectancy (LE_0_) in Nigeria and elsewhere in the world.^71^

Medical experts argued that the COVID-19 pandemic caused substantial increases in mortality across populations worldwide and overwhelmed health systems in many countries, potentially leading to increases in morbidity and mortality beyond the direct impact of COVID-19 infection.^71^ These increases in mortality, either direct or indirect, have caused stagnations or declines in life expectancy.^71^

A consultant clinical pharmacist and public health specialist, Dr. Kingsley Amibor, told The Guardian, “The average life expectancy for Nigerians in 2020 was 54.81 years.^71^ This represents an increase of 0.58 percent from 2019. This finding is a projection of the United Nations (UN) Organization and does not reflect the impacts of the Coronavirus disease (COVID-19) pandemic”.^71^

In Nigeria, life expectancy (LE_0_) was calculated at 54.49 years in 2019, 54.18 years in 2018, and 53.73 years in 2017, showing a progressive increase in life expectancy from 2017 to 2019.^71^ The figures represent an increase since 2016. For instance, there was a 0.84% increase from the 2016 value, a 0.83% increase from 2017, and a 0.58% increase from 2018 and 2019, respectively. The final figures for 2020 may represent a slight decrease because of the impact of COVID-19.^71^ Luckily for Nigeria, the reduction may not be significant because of the low case fatality for COVID-19 recorded in Nigeria, which was about two percent.^71^ Further to this information, Nigeria is ranked one of the countries with the lowest life expectancy figures in Africa (54.81 years, while a country like Tunisia also in Africa records 76.79 years).^71^

Thus, life expectancy observed a steady rise in low-income countries, including Nigeria, by as much as 21% between 2000 and 2016, due mainly to improved access to services to prevent and treat HIV, malaria, and tuberculosis, as well as several neglected tropical diseases such as guinea worm.^71^ Additionally, improved maternal and child healthcare led to a reduction of child mortality by almost 50% between 2000 and 2018.^71^

Not just COVID-19, but there are other causes of mortality, including infant mortality, HIV and AIDS, malaria, tuberculosis, poverty, malnutrition, road traffic accidents, and other causes that afflict African countries.^71^

In addition, non-communicable diseases such as heart diseases, strokes, diabetes, hypertension, and cancers caused 70% of all deaths in 2016, and about 85% of non-communicable diseases deaths occurred in low-income countries, including Nigeria.^71^

### Life expectancy (LE_0_) in Togo

The research team reviewed charts and tables of Togo’s life expectancy from 1950 to 2023.^72^ The United Nations projections were also included through the year 2100. It shows that the current life expectancy for Togo in 2023 was 62.13 years, a 0.52% increase from 2022. In addition, the life expectancy for Togo in 2022 was 61.81 years, a 0.52% increase from 2021.^72^ In 2021, life expectancy for Togo was 61.49 years, a 0.52% increase from 2020, while the life expectancy for Togo in 2020 was 61.17 years, and a 0.53% increase from 2019.^72^ Furthermore, the COVID-19 crisis exacerbated the plight of vulnerable people, especially people without homes. Lockdowns and social distancing measures deprived them of support, and they experienced more isolation, insecurity, and malnourishment than ever.^72^

### Promoting shielding measures

However, a report from Togo showed that Humanity & Inclusion (HI) teams conducted outreach work to limit the spread of COVID-19 in Lomé and the city of Santee Condji.^72^ Between April 2021 and February 2022, several actions were implemented to limit the spread of COVID-19 in Togo. For example, HI installed public showers, and more than 80,000 people were taken, with an average of 210 people per day benefiting from their services.^72^ In addition, with the help of awareness-raising sessions, almost 16,000 homeless people were taught how to protect themselves and others from COVID-19.^72^ Furthermore, a thousand information posters on COVID-19 prevention and vaccination were produced and distributed with more than 25,000 facemasks and 15,000 containers of hydro-alcoholic gel distributed. More than 8,000 hygiene kits were distributed, containing hydro-alcoholic gel, face masks, toothpaste, a toothbrush, soap, a sponge, and sanitary pads. In addition, around 100 peer educators were trained to share good practices and raise the awareness to others, and more than 1,000 people received COVID-19 vaccines.^72^

### Providing medical and psychological support

The report from Togo showed that two health surveillance teams, each with a nurse, a psychologist, and a midwife, conducted night rounds in Lomé during the COVID-19 pandemic. They provided medical and psychosocial care to over 15,000 people.^72^ Also, whenever possible, medical conditions, such as headaches, sores, rashes, malaria, and sexually transmitted diseases, were treated directly on-site.^34^ Thanks to these activities, more than 8,500 people accessed healthcare between April 2021 and February 2022. As part of the outreach work organized by HI, more than 4,500 people were given psychosocial support, as there was a real need among homeless people to talk about their past bad experiences and their day-to-day lives on the street.^72^

> *“We provide homeless people with psychological support because they are a sector that feels vulnerable and neglected. They feel seen when we offer them specialized services that are otherwise inaccessible. It is part of HI’s mission to give hope to people who feel forgotten”, explains Issa Afo, HI psychologist*.^72^

Given the insufficient testing and the under-reporting of cases and deaths in many countries, the actual number of deaths from COVID-19 is still believed to be considerably underestimated. Additionally, inconsistencies in public health systems globally in defining and classifying COVID-19 deaths and the indirect effects of the pandemic on other causes of death make it difficult to estimate an accurate pandemic death toll.^1,67,73,74^

Therefore, as the COVID-19 pandemic is still ongoing and causing profound challenges for humanity, it is vital to understand its effects on life expectancy, including its relevant characteristics and patterns.^1,67,73,74^

Our analyses of life expectancy show that the pandemic exacted a striking toll on population health in 2020 and 2021 across most West African countries. Only males and females in Togo successfully avoided drops in life expectancy (LE_0_) in our cross-national comparison of sixteen West African countries (Figures 3, Figure 5, Figure 6, and Figure 7). Early non-pharmaceutical interventions coupled with a robust healthcare system may have helped to explain some of these successes in Togo, including no losses in life expectancy.^72^

In contrast to other studies worldwide, the USA, followed by Eastern European countries such as Lithuania, Bulgaria, and Poland, experienced the most considerable losses in life expectancy in 2020, with more considerable losses in most countries for males than females. Among Western European countries, life expectancy losses were also documented.^73,74^

Consistent with our study on West Africa, emerging evidence from low-middle-income countries (such as Brazil and Mexico) shows that the pandemic substantially devastated those countries^75^, suggesting that life-expectancy losses are more considerable in these populations than previously predicted. In addition, losses in life expectancy in the sixteen West African countries varied substantially between subgroups (age groups and sexes) within countries and among countries (Additional files). However, more data is needed to limit direct and disaggregated comparisons across various countries. However, these are urgently needed to understand the full mortality impacts of the COVID-19 pandemic.

This limitation is manifested in a study on the impact of COVID-19 in Europe, which included only two countries outside of Europe with reliable and complete information for 2020, whereas the analysis focused on national populations.^76^ It is crucial that in the future, analyses should be broadened to include a set of countries, data on all-cause mortality with detailed disaggregation by age and sex, and deeper analysis of other characteristics, for example, socioeconomic status and ethnicity, which are needed to assess the uneven mortality impacts of the pandemic. However, we are cognizant of deficiencies and limited vital registrations (Birth and Death rates), the actual mortality impacts of the COVID-19 pandemic that may never fully be known. The minimum can be data from contexts requiring further adjustments, for example, undercounting and age stacking, before it can be used. As much as COVID-19 might have been seen as a transient shock to life expectancy, the evidence of potential long-term morbidity due to long COVID and impacts of delayed care for other illnesses is likely to be seen in the future.^40,41^

In addition, the health effects and widening inequalities stemming from the social, mental health, psychosocial, and economic disruptions of the COVID-19 pandemic^42^ suggest that the scars of the COVID-19 pandemic on the population’s health will be long-lasting.

In summary, life expectancy is primarily a public health measure that can be compared across nations and subpopulations. It has been used to indicate economic successes and medical care’s effectiveness. Life expectancy at birth (LE_0_) is a commonly used indicator and a critical summary measure of the health and wellbeing of a population. It is one of the most widely used metrics for measuring population health and longevity. It refers to the average number of years a hypothetical cohort of people would live if they were to experience the death rates observed in a given period throughout their lifespan. Thus, life expectancy (LE_0_) is estimated, based on the death rates for a given period. Over a period, life expectancy at birth has increased significantly in most countries, globally.^77^

Also, life disparity is one measure of lifespan variation, representing the average remaining life expectancy at the age when death occurs. It is a measure of life years lost due to death and it moves in the opposite direction to life expectancy (LE_0_).^24^

Furthermore, the Gini Coefficient (Gini index) is a synthetic indicator that captures the level of inequality for a given variable and population. It varies between 0 (perfect equality) and 1 (extreme inequality). Between 0 and 1, where the higher the Gini index, the greater the inequality.^25^ So, life inequality years between people within the same country are measured with the Gini coefficient.^25^ A high Gini coefficient means there is a significant within-country inequalities exist in the number of years people live. (Life disparity and Gini coefficient are the other life span measures that can be used other than LE).^24,25^ In the case of the West African countries, life disparity and Gini coefficient have shown improvement over the years of study (1990-2021).

## Conclusion

Although most West African countries posted progressive LE_0_ gains from 1990 to 2021, there were LE_0_ losses in 2020 and 2021 when the COVID-19 pandemic emerged. The West African region has the lowest LE_0_ of all African regions probably due to lower socio-economic indicators compared to all other African regions. Also, during the COVID-19 pandemic in 2020 and 2021, there were LE_0_ losses in all West African countries, except Togo. In addition, LE_0_ gaps between males and females were highest in the late 1990s and least during the late 2000s. Even though several studies reported that morbidity and mortality rates of COVID-19 were lower in Africa than in the rest of the world, a more comprehensive study is warranted to assess the actual impact of COVID-19 on West African countries.

## Data Availability

All data produced in the present study are available upon reasonable request to the authors

## Abbreviations

AIDS: Acquired immunodeficiency syndrome
COVID-19: Coronavirus disease-19
HIV: Human immunodeficiency virus
(LE_0_): Life expectancy at birth
SARS-CoV-2: Severe acute respiratory coronavirus-2
WHO: World Health Organization.

## Declarations

### Ethics approval and consent to participate

This study on life expectancy in the population of West Africa was obtained from a public repository, and we conducted secondary data analysis. In addition, the study followed relevant institutional guidelines and regulations on managing open data sources.

### Consent to publish

Not applicable.

### Availability of data and material

All datasets supporting this article’s conclusion are within this paper and are accessible by a reasonable request to the corresponding author.

## Authors’ contributions

DLK, GB, and JA designed this study. JA, GB, and DLK supervised data management. JA, GB, and DLK analyzed and interpreted the data. JA, GB, EO, RN, CL, and DLK wrote and revised the manuscript. All Authors approved the manuscript.

## Authors’ Information

Prof. David Lagoro Kitara (DLK) is a Takemi fellow of Harvard University and a Professor at Gulu University, Faculty of Medicine, Department of Surgery, Gulu City, Uganda; Dr. Gaye Bamba (GB) is a founder member of the African Research Network (ARN), Dakar, Senegal; Joelle Abi Abboud (JA) is Research manager at the African Research Network (ARN), Paris, France; Dr. Ritah Nantale (RN) is Manager at Busitema University, Faculty of Health Sciences, Department of Maternal and Child health, Mbale City, Uganda; Camille Lassale (CL) is at Barcelona Institute for Global Health (ISGlobal), Barcelona, Spain. Dr. Emmanuel Olal (EO) is a public health specialist and a physician a YotKom medical Centre, Kitgum Municipality, Uganda.

## Acknowledgment

We thank the assistance from WPP for the datasets obtained and Joelle Abi Abboud for a comprehensive data analysis of this study.

In 2022, 67.4 percent of people aged 15 years and above in Africa were able to read and write a simple statement and understand it. Regionally, Southern Africa presented the highest literacy rate, at 80 percent. North and East Africa had similar shares of literate people, at over 71 percent. In contrast, 67.5 percent and 54 percent of the adult population in Central and West Africa could read and write.

**Saifaddin Galal. Adult literacy rate in Africa as of 2022, by region. Statistica. 2023**. https://www.statista.com/statistics/1233204/adult-literacy-rate-in-africa-by-region/#:~:text=North%20and%20East%20Africa%20had,Africa%20could%20read%20and%20write.

## References

1. Aburto JM, Kashyap R, Scholey J. Estimating the burden of the COVID-19 pandemic on mortality, life expectancy and lifespan inequality in England and Wales: a population-level analysis. J Epidemiol Community Health 2021;75:735–40.

2. Karlinsky A, Kobak D. Tracking excess mortality across countries during the COVID-19 pandemic with the world mortality dataset. ELife 2021;10:e69336.

3. Goldstein JR, Lee RD. Demographic perspectives on the mortality of COVID-19 and other epidemics. Proc Natl Acad Sci USA. 2020;117:22035–41.

4. Ortiz-Ospina Esteban. “Life expectancy”-What does this actually mean? Our World in Data. 2017. Available Online: https://ourworldindata.org, Accessed on March 2, 2019.

5. Aboriginal and Torres Strait Islander Health Performance Framework (HPF). Tier 1-Life expectancy and wellbeing. 2012. Available Online: https://www.health.gov.au. Accessed on March 2, 2019.

6. Statistica. Life Expectancy in Africa. 2018. Available Online: https://www.statista.com, Accessed June 22, 2020.

7. Nkalu CN, Edeme RK. Environmental Hazards and Life Expectancy in Africa: Evidence from GARCH Model, SAGE Open. 2019:1–8. DOI:10.1177/2158244019830500.

8. Otekunrin OA, Otekunrin OA, Momoh S and Ayinde IA. How far has Africa gone in achieving the zero-hunger target? Evidence from Nigeria. Global Food Security. 2019a;22:1–12.

9. Uchendu FN. Hunger influenced life expectancy in war-torn Sub-Saharan African countries. Journal of Health, Population and Nutrition. 2018;37:11. 10.1186/s41043-018-0143-3.

10. Mondal NI, Shitan M. Impact of Socio-Health Factors on Life Expectancy in the Low and Lower Middle-Income Countries. Iranian Journal of Public Health. 2013;42(12):1354–1362.

11. Wiysonge CS. People are living longer in Africa, but the rise of lifestyle diseases threatens progress. Quartz Africa. 2018. https://qz.com/africa/1475911/, Accessed June 22, 2020.

12. WHO. COVID-19 situations by WHO regions. 2023.https://covid19.who.int/?mapFilter=vaccinations

13. Dowd JB, Andriano L, Brazel DM et al. Demographic science aids in understanding the spread and fatality rates of COVID-19. Proc Natl Acad Sci USA. 2020;117:9696–98.

14. Pifarré I Arolas H, Acosta E, Ló pez-Casasnovas G. Years of life lost to COVID-19 in 81 countries. Sci Rep 2021;11:3504.

15. Otekunrin OA, Momoh S, Ayinde IA and Otekunrin OA. How far has Africa gone in achieving sustainable development goals? Exploring African dataset, Data in Brief. 2019b;27:104647.

16. United Nations. World Population Prospects—Population Division—United Nations. 2021. https://population.un.org/wpp/Download/Standard/Population/ (26 January 2021, date last accessed).

17. Aburto JM, Villavicencio F, Basellini U, Kjærgaard S, Vaupel JW. Dynamics of life expectancy and life span equality. Proc Natl Acad Sci USA. 2020;117:5250–59.

18. Mehta NK, Abrams LR, Myrskyla M. US life expectancy stalls due to cardiovascular disease, not drug deaths. Proc Natl Acad Sci USA. 2020;117:6998–7000.

19. Leon DA, Jdanov DA, Shkolnikov VM. Trends in life expectancy and age-specific mortality in England and Wales, 1970–2016, in comparison with a set of 22 high-income countries: an analysis of vital statistics data. Lancet Public Health 2019;4:e575–82.

20. Fenton L, Minton J, Ramsay J. Recent adverse mortality trends in Scotland: comparison with other high-income countries. BMJ Open 2019;9:e029936.

21. OCHA services. World Population Prospects 2022: Summary of Results. Reliefweb. 2022. https://reliefweb.int/report/world/world-population-prospects-2022-summary-results?

22. World Population Prospects - Population Division - United Nations [Internet]. [cited 2023 Aug 1]. Available from: https://population.un.org/wpp/Download/Standard/Mortality/

23. Aburto JM, Schöley J, Kashnitsky I, Zhang L, Rahal C, Missov TI, et al. Quantifying impacts of the COVID-19 pandemic through life-expectancy losses: a population-level study of 29 countries. Int J Epidemiol. 2022;51(1):63–74.

24. Vaupel JW, Zhang Z, van Raalte AA. Life expectancy and disparity: an international comparison of life table data. BMJ Open. 2011;1(1):e000128. doi:10.1136/bmjopen-2011-000128.

25. Adam Hayes. Gini Index Explained and Gini Co-efficient Around the World. Investopedia. 2023. https://www.investopedia.com/terms/g/gini-index.asp

26. Softonic [Internet]. [cited 2023 Aug 7]. Download RStudio Desktop - free - latest version. Available from: https://rstudio-desktop.en.softonic.com.

27. Timothy C, Jacqueline N Milton. Basic Statistical Analysis Using the R Statistical Package. RStudio Team. 2021. https://sphweb.bumc.bu.edu/otlt/MPH-Modules/BS/R/R-Manual/R-Manual_print.html.

28. Jessica Y Ho, Arun S Hendi. Recent trends in life expectancy across high income countries: retrospective observational study. BMJ. 2018;362:k2562. |doi:10.1136/bmj.k2562.

29. Bongaarts J. How long will we live? Popul Dev Rev 2006;32:605–28. doi:10.1111/j.1728-4457.2006.00144.x.

30. Wilmoth JR. Demography of longevity: past, present, and future trends. Exp Gerontol. 2000;35:1111–29. doi:10.1016/S0531-5565(00)00194-7.

31. Shin I. The effect of pension on the optimized life expectancy and lifetime utility level. MPRA Paper, No. 2013;41374: 1–27.

32. Hazan M. Life expectancy and schooling: New insights from cross-country data. Journal of Population Economics. 2012;25(4):1237–1248.

33. Balan C and E Jaba. Statistical analysis of the determinants of life expectancy in Romania. Romanian Journal of Regional Science. 2011;5(2):28–38.

34. Halicioglu F. Modeling life expectancy in Turkey. MPRA Paper, No. 30840: 2010:1–31.

35. Bergh A and TH Nilsson. Good for living? On the relation between globalization and life expectancy. World Development. 2009;38(9):1191–1203.

36. Mariani F, P Barahona and N Raffin. Life expectancy and the environment. CES Working Papers, No.48: 2008:1–23.

37. Yavari K and M Mehrnoosh. Determinants of life expectancy: A cross – country. Iranian Economic Review. 2006;11(15):13–142.

38. Leung MCM and Y Wang. Endogenous health care and life expectancy in a neoclassical growth model. Singapore: World Scientific Publishing Co. 2003.

39. Cervellati M and U Sunde. Human capital formation, life expectancy and the process of economic development. IZA Discussion Paper, No. 585. 2002;1–29.

40. Castello A and Domenech R. Human capital inequality and economic growth: Some new evidence. Economic Journal. 2002;112(478):187–200.

41. Wilmoth JR. Demography of longevity: past, present, and future trends. Exp Gerontol. 2000;35(9-10):1111–1129.

42. Vagero D. Health inequalities across the globe demand new global policies. Scand J Public Health. 2007;35(2):113–115.

43. Moser K, Shkolnikov V, Leon DA. World mortality 1950-2000: divergence replaces convergence from the late 1980s. Bull World Health Organ. 2005;83(3):202–209.

44. Shen CE, Williamson JB. Child mortality, women’s status, economic dependency, and state strength: a cross-national study of less developed countries. Soc Forces. 1997;76(2):667–694.

45. Mahfuz K. Determinants of life expectancy in developing countries. J Dev Areas 2008, 41(2):185–204.

46. Franco A, Alvarez-Dardet C, Ruiz MT. Effect of democracy on health: ecological study. BMJ. 2004;329(7480):1421–1423.

47. Navarro V, Muntaner C, Borrell C, Benach J, Quiroga A, Rodriguez-Sanz M, Verges N, Pasarin MI: Politics and health outcomes. Lancet. 2006;368(9540):1033–1037.

48. Lake DA, Baum MA: The invisible hand of democracy: political control and the provision of public services. Comp Polit Stud. 2001;34(6):587–621.

49. Ro-Ting Lin, Ya-Mei Chen, Lung-Chang Chien, and Chang-Chuan Chan. Political and social determinants of life expectancy in less developed countries: a longitudinal study. BMC Public Health. 2012;12:85.

50. Flegg AT. Inequality of income, illiteracy, and medical care as determinants of infant mortality in underdeveloped countries. Popul Stud. 1982;36(3):441–458.

51. Ruger JP, Kim HJ: Global health inequalities: an international comparison. J Epidemiol Community Health. 2006;60(11):928–936.

52. McMichael AJ, McKee M, Shkolnikov V, Valkonen T. Mortality trends and setbacks: global convergence or divergence? Lancet. 2004;363(9415):1155–1159.

53. McMichael AJ, Butler CD. Emerging health issues: the widening challenge for population health promotion. Health Promot Int. 2006;21(Suppl 1):15–24.

54. Guogui Huang, Fei Guo, Klaus F Zimmermann, Lihua Liu, Lucy Taksa, Zhiming Cheng, et al. The effect of COVID-19 pandemic in 27 countries. Scientific Reports. 2023;13:8911.

55. David Lagoro Kitara, Eric Nzirakaindi Ikoona. COVID-19 pandemic, Uganda’s Story. Pan Afr Med J. 2020; 35:51.

56. David Lagoro Kitara, Eric Nzirakaindi Ikoona. Proposed strategies for easing COVID-19 lockdown measures in Africa. Pan Afr Med J, 2020; 36(179): 10.11604/pamj.2020.36.179.24194.

57. David Lagoro Kitara, Eric Nzirakaindi Ikoona. A proposed framework to limit post-lockdown community transmission of COVID-19 in Africa. Pan Afr Med J. 2021;38:303.

58. Adams J, MacKenzie MJ, Amegah AK, Ezeh A, Gadanya MA, Omigbodun A, et al. The Conundrum of Low COVID-19 Mortality Burden in sub-Saharan Africa: Myth or Reality? Glob Health Sci Pract. 2021;9(3):433–443.

59. Williamson EJ, Walker AJ, Bhaskaran K. Factors associated with COVID-19-related death using OpenSAFELY. Nature. 2020;584(7821):430–436.

60. Osei SA, Biney RP, Anning AS. Low incidence of COVID-19 case severity and mortality in Africa; Could malaria co-infection provide the missing link? BMC Infect Dis. 2022; 22:78.

61. Jyoti Dalal, Benedict Nguimbis, Gabriela Guizzo Dri, Akarsh Venkatasubramanian, Lucie Noubi Tchoupopnou Royd, Sara Botero Mesa, et al. COVID-19 mortality in women and men in sub-Saharan Africa: a cross-sectional study. BMJ Global Health 2021;6:e007225.

62. Steven Baguma, Christopher Okot, Nelson Alema Onira, Judith Aloyo, Eric Nzirakaindi Ikoona, David Lagoro Kitara et al. Factors associated with mortality among the COVID-19 patients treated at Gulu Regional Referral Hospital: A retrospective study. Research Square. 2021. DOI:10.21203/rs.3.rs-1191937/v1.

63. Okot C, Baguma S, Onira NA, Judith Aloyo, Eric Nzirakaindi Ikoona, David Lagoro Kitara et al. Characteristics of the COVID-19 patients treated at Gulu Regional Referral Hospital, Northern Uganda: A cross-sectional study. J Infect Dis Ther. 2022; S1:004.

64. Eric Nzirakaindi Ikoona, Denis Acullu, Johnson Oloya Nyeko, Judith Aloyo, David Lagoro Kitara, et al. COVID-19 pandemic, challenges, and opportunities in Northern Uganda; Community overview and perspectives: A qualitative study using informant interviews. Advance Journal of virology, epidemic, and pandemic diseases. 2022;7(1):63–71.

65. Steven Baguma, Christopher Okot, Judith Aloyo, Eric Nzirakaindi Ikoona, David Lagoro Kitara, et al. Factors associated with mortality among the COVID-19 patients treated at Gulu Regional Referral Hospital: A retrospective study. Front. Public Health. 2022. 10:841906. Doi:10.3389/pubh.2022.841906.

66. Judith Aloyo, Denis Acullu, Freddy Wathum Drinkwater Oyat, Lawence Obalim, Eric Nzirakaindi Ikoona, and David Lagoro Kitara, et al. Distinct Characteristics of the COVID-19 among Children and Young Adolescents Treated at Gulu Regional Referral Hospital; Northern Uganda: A Cross-Sectional Study. J Infect Dis Ther. 2022, 10:S1:1000004.

67. Aburto JM. Quantifying impacts of the COVID-19 pandemic through life expectancy losses. Int. J. Epidemiol. 2021;51:63–74.

68. Heuveline P & Tzen M. Beyond deaths per capita: Comparative COVID-19 mortality indicators. BMJ Open. 2021;11:e042934.

69. Woolf SH, Masters RK & Aron LY. Effect of the covid-19 pandemic in 2020 on life expectancy across populations in the USA and other high-income countries: simulations of provision high-income data. BMJ. 2021;373:n1343.

70. Andrasfay T & Goldman N. Association of the COVID-19 pandemic with estimated life expectancy by race/ethnicity in the United States, 2020. JAMA Netw. Open. 2021;4:e2114520–e2114520.

71. Chukwuma Muanya. Probing impact of COVID-19 on life expectancy. The Guardian. 2020. https://guardian.ng/features/health/probing-impact-of-covid-19-on-life-expectancy/

72. Togo: helping homeless people cope with COVID-19. Humanity and Inclusion. 2022. https://www.hi-us.org/en/news/togo-helping-homeless-people-cope-with-covid-19

73. Modig K, Ahlbom A & Ebeling M. Excess mortality from COVID-19: Weekly excess death rates by age and sex for Sweden and its most affected region. Eur. J. Public Health. 2021;31:17–22.

74. Karanikolos M & McKee M. How comparable is COVID-19 mortality across countries? Eurohealth. 2020;26:45–50.

75. Conyon MJ, He L, Thomsen S. Lockdowns and COVID-19 Deaths in Scandinavia. Rochester, NY: Social Science Research Network, 2020.

76. Juranek S, Zoutman F. The Effect of Non-pharmaceutical Interventions on the Demand for Health Care and Mortality: Evidence on COVID-19 in Scandinavia. Rochester, NY: Social Science Research Network, 2020.

77. Muniyandi M, Singh PK, Aanandh Y, Karikalan N and Padmapriyadarsini C. A national-level analysis of life expectancy associated with the COVID-19 pandemic in India. Front. Public Health. 2022;10:1000933.

