## Additional Table files 1-34 for "Quantifying life-expectancy Losses and Gains over 31 years (1990-2021): A population-level study on West African Countries"

**Additional files: Linear regression analyses on LE_0_ for West African countries across 31 years (1990-2021)**

**Table 1: Linear regression table of LE_0_ in Benin across 31 years (1990-2021).**

|  | **^ex^** | | |
| --- | --- | --- | --- |
| **^Predictors^** | **^Estimates^** | **^CI^** | **^p^** |
| **^(Intercept)^** | **^60.31^** | **^59.86 – 60.75^** | **^<0.001^** |
| **^Year [1991]^** | **^0.06^** | **^-0.53 – 0.65^** | **^0.844^** |
| **^Year [1992]^** | **^0.16^** | **^-0.43 – 0.75^** | **^0.606^** |
| **^Year [1993]^** | **^0.25^** | **^-0.34 – 0.84^** | **^0.399^** |
| **^Year [1994]^** | **^0.33^** | **^-0.26 – 0.91^** | **^0.280^** |
| **^Year [1995]^** | **^0.40^** | **^-0.19 – 0.99^** | **^0.180^** |
| **^Year [1996]^** | **^0.44^** | **^-0.15 – 1.03^** | **^0.145^** |
| **^Year [1997]^** | **^0.54^** | **^-0.05 – 1.13^** | **^0.073^** |
| **^Year [1998]^** | **^0.56^** | **^-0.03 – 1.15^** | **^0.065^** |
| **^Year [1999]^** | **^0.61^** | **^0.02 – 1.20^** | **^0.042^** |
| **^Year [2000]^** | **^0.54^** | **^-0.05 – 1.13^** | **^0.075^** |
| **^Year [2001]^** | **^0.48^** | **^-0.11 – 1.07^** | **^0.111^** |
| **^Year [2002]^** | **^0.36^** | **^-0.23 – 0.95^** | **^0.236^** |
| **^Year [2003]^** | **^0.33^** | **^-0.26 – 0.92^** | **^0.273^** |
| **^Year [2004]^** | **^0.28^** | **^-0.30 – 0.87^** | **^0.344^** |
| **^Year [2005]^** | **^0.34^** | **^-0.25 – 0.93^** | **^0.252^** |
| **^Year [2006]^** | **^0.42^** | **^-0.17 – 1.01^** | **^0.160^** |
| **^Year [2007]^** | **^0.45^** | **^-0.14 – 1.04^** | **^0.131^** |
| **^Year [2008]^** | **^0.47^** | **^-0.12 – 1.06^** | **^0.115^** |
| **^Year [2009]^** | **^0.44^** | **^-0.15 – 1.03^** | **^0.140^** |
| **^Year [2010]^** | **^0.58^** | **^-0.01 – 1.17^** | **^0.055^** |
| **^Year [2011]^** | **^0.55^** | **^-0.04 – 1.14^** | **^0.066^** |
| **^Year [2012]^** | **^0.61^** | **^0.02 – 1.20^** | **^0.041^** |
| **^Year [2013]^** | **^0.66^** | **^0.07 – 1.25^** | **^0.028^** |
| **^Year [2014]^** | **^0.67^** | **^0.08 – 1.26^** | **^0.026^** |
| **^Year [2015]^** | **^0.72^** | **^0.13 – 1.31^** | **^0.016^** |
| **^Year [2016]^** | **^0.74^** | **^0.15 – 1.33^** | **^0.014^** |
| **^Year [2017]^** | **^0.83^** | **^0.24 – 1.42^** | **^0.006^** |
| **^Year [2018]^** | **^0.89^** | **^0.30 – 1.48^** | **^0.003^** |
| **^Year [2019]^** | **^0.97^** | **^0.38 – 1.56^** | **^0.001^** |
| **^Year [2020]^** | **^0.36^** | **^-0.23 – 0.95^** | **^0.225^** |
| **^Year [2021]^** | **^-0.02^** | **^-0.61 – 0.57^** | **^0.956^** |
| **^Age^** | **^-0.67^** | **^-0.67 – -0.66^** | **^<0.001^** |
| **^Gender [Male]^** | **^-1.56^** | **^-1.70 – -1.41^** | **^<0.001^** |
| **^Observations^** | **^6464^** | | |
| **^R2 / R2 adjusted^** | **^0.977 / 0.976^** | | |

Table 1 shows the linear regression of LE for Benin taking into consideration the factor, years (1990 to 2021), age (from 0-100), and gender (Male and Female). The formula for all tables: LE=β_0_+β_1_⋅Year+β_2_⋅age+β_3_⋅sexe+e.
In these tables, we compared factors; years and female gender to the year 1990. LE in Benin increased compared to 1990 in all the years (estimates positives) and became significant from 2012 till 2019. In 2020 and 2021 (during the COVID-19 pandemic), LE decreased although insignificantly compared to 1990. From the estimate observed, LE in 2019 was the highest in the 30 year LE estimates from 1990.

**Table 2: Comparison of LE_0_ between males and females in Benin (1990-2021).**

|  | **ex** | | |
| --- | --- | --- | --- |
| **Predictors** | **Estimates** | **CI** | **p** |
| (Intercept) | 61.28 | 60.84 – 61.72 | **<0.001** |
| Year [1990] | -0.97 | -1.56 – -0.38 | **0.001** |
| Year [1991] | -0.91 | -1.50 – -0.32 | **0.002** |
| Year [1992] | -0.82 | -1.41 – -0.23 | **0.007** |
| Year [1993] | -0.72 | -1.31 – -0.13 | **0.017** |
| Year [1994] | -0.65 | -1.24 – -0.06 | **0.031** |
| Year [1995] | -0.57 | -1.16 – 0.02 | 0.058 |
| Year [1996] | -0.53 | -1.12 – 0.06 | 0.076 |
| Year [1997] | -0.43 | -1.02 – 0.16 | 0.149 |
| Year [1998] | -0.42 | -1.01 – 0.17 | 0.165 |
| Year [1999] | -0.36 | -0.95 – 0.23 | 0.228 |
| Year [2000] | -0.44 | -1.03 – 0.15 | 0.147 |
| Year [2001] | -0.49 | -1.08 – 0.10 | 0.101 |
| Year [2002] | -0.62 | -1.21 – -0.03 | **0.040** |
| Year [2003] | -0.64 | -1.23 – -0.05 | **0.033** |
| Year [2004] | -0.69 | -1.28 – -0.10 | **0.022** |
| Year [2005] | -0.63 | -1.22 – -0.04 | **0.037** |
| Year [2006] | -0.55 | -1.14 – 0.04 | 0.067 |
| Year [2007] | -0.52 | -1.11 – 0.07 | 0.085 |
| Year [2008] | -0.50 | -1.09 – 0.09 | 0.097 |
| Year [2009] | -0.53 | -1.12 – 0.06 | 0.078 |
| Year [2010] | -0.40 | -0.99 – 0.19 | 0.187 |
| Year [2011] | -0.42 | -1.01 – 0.17 | 0.163 |
| Year [2012] | -0.36 | -0.95 – 0.23 | 0.232 |
| Year [2013] | -0.31 | -0.90 – 0.28 | 0.302 |
| Year [2014] | -0.30 | -0.89 – 0.29 | 0.314 |
| Year [2015] | -0.25 | -0.84 – 0.34 | 0.407 |
| Year [2016] | -0.24 | -0.83 – 0.35 | 0.434 |
| Year [2017] | -0.15 | -0.74 – 0.44 | 0.625 |
| Year [2018] | -0.08 | -0.67 – 0.51 | 0.788 |
| Year [2020] | -0.61 | -1.20 – -0.02 | **0.043** |
| Year [2021] | -0.99 | -1.58 – -0.40 | **0.001** |
| Age | -0.67 | -0.67 – -0.66 | **<0.001** |
| Gender [Male] | -1.56 | -1.70 – -1.41 | **<0.001** |
| Observations | 6464 | | |
| R^2^ / R^2^ adjusted | 0.977 / 0.976 | | |

Table 2 shows that 2019 had the highest LE (estimates of all years were negative meaning that LE was lower compared to 2019). It shows that from (1990 till 1994, 200-2005 and from 2020-2021), LE was significantly lower than 2019 and can be seen in age and gender.

**Table 3: Linear regression table of LE_0_ for Burkina Faso (1990-2021).**

|  | **ex** | | |
| --- | --- | --- | --- |
| **Predictors** | **Estimates** | **CI** | **p** |
| (Intercept) | 57.41 | 56.93 – 57.89 | **<0.001** |
| Year [1991] | -0.01 | -0.65 – 0.63 | 0.971 |
| Year [1992] | -0.03 | -0.67 – 0.61 | 0.926 |
| Year [1993] | -0.16 | -0.80 – 0.48 | 0.618 |
| Year [1994] | -0.14 | -0.78 – 0.50 | 0.676 |
| Year [1995] | -0.12 | -0.76 – 0.52 | 0.721 |
| Year [1996] | -0.11 | -0.75 – 0.53 | 0.732 |
| Year [1997] | -0.26 | -0.90 – 0.38 | 0.428 |
| Year [1998] | -0.11 | -0.75 – 0.53 | 0.731 |
| Year [1999] | -0.05 | -0.69 – 0.59 | 0.886 |
| Year [2000] | 0.11 | -0.53 – 0.75 | 0.740 |
| Year [2001] | 0.10 | -0.54 – 0.74 | 0.752 |
| Year [2002] | 0.10 | -0.54 – 0.74 | 0.757 |
| Year [2003] | 0.21 | -0.44 – 0.85 | 0.530 |
| Year [2004] | 0.33 | -0.31 – 0.97 | 0.307 |
| Year [2005] | 0.47 | -0.17 – 1.11 | 0.149 |
| Year [2006] | 0.61 | -0.03 – 1.25 | 0.062 |
| Year [2007] | 0.74 | 0.10 – 1.38 | **0.024** |
| Year [2008] | 1.00 | 0.36 – 1.64 | **0.002** |
| Year [2009] | 1.13 | 0.49 – 1.77 | **0.001** |
| Year [2010] | 1.23 | 0.59 – 1.87 | **<0.001** |
| Year [2011] | 1.38 | 0.74 – 2.02 | **<0.001** |
| Year [2012] | 1.49 | 0.85 – 2.13 | **<0.001** |
| Year [2013] | 1.47 | 0.83 – 2.11 | **<0.001** |
| Year [2014] | 1.62 | 0.98 – 2.26 | **<0.001** |
| Year [2015] | 1.74 | 1.10 – 2.38 | **<0.001** |
| Year [2016] | 1.87 | 1.23 – 2.51 | **<0.001** |
| Year [2017] | 1.87 | 1.23 – 2.51 | **<0.001** |
| Year [2018] | 2.02 | 1.38 – 2.66 | **<0.001** |
| Year [2019] | 1.95 | 1.31 – 2.59 | **<0.001** |
| Year [2020] | 1.37 | 0.73 – 2.01 | **<0.001** |
| Year [2021] | 0.90 | 0.26 – 1.54 | **0.006** |
| Age | -0.65 | -0.65 – -0.65 | **<0.001** |
| Gender [Male] | -1.61 | -1.77 – -1.45 | **<0.001** |
| Observations | 6464 | | |
| R^2^ / R^2^ adjusted | 0.971 / 0.971 | | |

Table 3 shows that LE decreased insignificantly from 1990 until 1999. In 2000, LE increased insignificantly until 2006. But from 2007, it increased significantly to the highest level in 2018 and decreased significantly from 2019 to 2021.

**Table 4: Comparison of LE_0_ between males and females in Burkina Faso (1990-2021).**

|  | **ex** | | |
| --- | --- | --- | --- |
| **Predictors** | **Estimates** | **CI** | **p** |
| (Intercept) | 59.37 | 58.89 – 59.85 | **<0.001** |
| Year [1990] | -1.95 | -2.59 – -1.31 | **<0.001** |
| Year [1991] | -1.97 | -2.61 – -1.33 | **<0.001** |
| Year [1992] | -1.98 | -2.63 – -1.34 | **<0.001** |
| Year [1993] | -2.12 | -2.76 – -1.48 | **<0.001** |
| Year [1994] | -2.09 | -2.73 – -1.45 | **<0.001** |
| Year [1995] | -2.07 | -2.71 – -1.43 | **<0.001** |
| Year [1996] | -2.07 | -2.71 – -1.43 | **<0.001** |
| Year [1997] | -2.21 | -2.85 – -1.57 | **<0.001** |
| Year [1998] | -2.07 | -2.71 – -1.43 | **<0.001** |
| Year [1999] | -2.00 | -2.64 – -1.36 | **<0.001** |
| Year [2000] | -1.85 | -2.49 – -1.21 | **<0.001** |
| Year [2001] | -1.85 | -2.49 – -1.21 | **<0.001** |
| Year [2002] | -1.85 | -2.49 – -1.21 | **<0.001** |
| Year [2003] | -1.75 | -2.39 – -1.11 | **<0.001** |
| Year [2004] | -1.62 | -2.26 – -0.98 | **<0.001** |
| Year [2005] | -1.48 | -2.12 – -0.84 | **<0.001** |
| Year [2006] | -1.34 | -1.98 – -0.70 | **<0.001** |
| Year [2007] | -1.22 | -1.86 – -0.58 | **<0.001** |
| Year [2008] | -0.95 | -1.59 – -0.31 | **0.004** |
| Year [2009] | -0.82 | -1.46 – -0.18 | **0.012** |
| Year [2010] | -0.73 | -1.37 – -0.09 | **0.026** |
| Year [2011] | -0.57 | -1.21 – 0.07 | 0.081 |
| Year [2012] | -0.46 | -1.10 – 0.18 | 0.159 |
| Year [2013] | -0.48 | -1.12 – 0.16 | 0.141 |
| Year [2014] | -0.34 | -0.98 – 0.30 | 0.301 |
| Year [2015] | -0.21 | -0.85 – 0.43 | 0.516 |
| Year [2016] | -0.08 | -0.72 – 0.56 | 0.797 |
| Year [2017] | -0.08 | -0.72 – 0.56 | 0.796 |
| Year [2018] | 0.07 | -0.57 – 0.71 | 0.836 |
| Year [2020] | -0.58 | -1.22 – 0.06 | 0.075 |
| Year [2021] | -1.06 | -1.70 – -0.42 | **0.001** |
| Age | -0.65 | -0.65 – -0.65 | **<0.001** |
| Gender [Male] | -1.61 | -1.77 – -1.45 | **<0.001** |
| Observations | 6464 | | |
| R^2^ / R^2^ adjusted | 0.971 / 0.971 | | |

Table 4 compares the LE between males and females from 1990 to 2021. Differences in LE between males and females were insignificant until 2020. In all the years, except 2018, LE among males was lower (estimates negative) although it was not statistically significant. However, in 2021 differences in LE between males and females became statistically significant compared to 2019.

**Table 6: Linear regression table of LE_0_ for Cape Verde (1990-2021).**

|  | **ex** | | |
| --- | --- | --- | --- |
| **Predictors** | **Estimates** | **CI** | **p** |
| (Intercept) | 67.82 | 67.29 – 68.35 | **<0.001** |
| Year [1991] | -0.01 | -0.72 – 0.70 | 0.982 |
| Year [1992] | 0.02 | -0.69 – 0.73 | 0.953 |
| Year [1993] | -0.03 | -0.74 – 0.68 | 0.933 |
| Year [1994] | -0.08 | -0.79 – 0.62 | 0.816 |
| Year [1995] | -0.44 | -1.15 – 0.27 | 0.221 |
| Year [1996] | 0.03 | -0.68 – 0.74 | 0.928 |
| Year [1997] | 0.26 | -0.45 – 0.96 | 0.480 |
| Year [1998] | 0.44 | -0.27 – 1.15 | 0.224 |
| Year [1999] | 0.58 | -0.13 – 1.29 | 0.109 |
| Year [2000] | 1.06 | 0.35 – 1.77 | **0.003** |
| Year [2001] | 1.75 | 1.04 – 2.46 | **<0.001** |
| Year [2002] | 2.14 | 1.43 – 2.85 | **<0.001** |
| Year [2003] | 2.53 | 1.83 – 3.24 | **<0.001** |
| Year [2004] | 2.88 | 2.17 – 3.59 | **<0.001** |
| Year [2005] | 3.30 | 2.59 – 4.01 | **<0.001** |
| Year [2006] | 3.00 | 2.29 – 3.71 | **<0.001** |
| Year [2007] | 3.45 | 2.75 – 4.16 | **<0.001** |
| Year [2008] | 3.15 | 2.44 – 3.85 | **<0.001** |
| Year [2009] | 3.64 | 2.94 – 4.35 | **<0.001** |
| Year [2010] | 3.85 | 3.14 – 4.56 | **<0.001** |
| Year [2011] | 4.29 | 3.58 – 5.00 | **<0.001** |
| Year [2012] | 4.26 | 3.55 – 4.96 | **<0.001** |
| Year [2013] | 4.64 | 3.93 – 5.35 | **<0.001** |
| Year [2014] | 4.66 | 3.95 – 5.36 | **<0.001** |
| Year [2015] | 4.13 | 3.42 – 4.84 | **<0.001** |
| Year [2016] | 4.78 | 4.07 – 5.49 | **<0.001** |
| Year [2017] | 5.28 | 4.57 – 5.99 | **<0.001** |
| Year [2018] | 4.59 | 3.88 – 5.30 | **<0.001** |
| Year [2019] | 4.71 | 4.00 – 5.42 | **<0.001** |
| Year [2020] | 3.73 | 3.02 – 4.44 | **<0.001** |
| Year [2021] | 3.16 | 2.45 – 3.87 | **<0.001** |
| Age | -0.76 | -0.76 – -0.76 | **<0.001** |
| Gender [Male] | -4.25 | -4.42 – -4.07 | **<0.001** |
| Observations | 6464 | | |
| R^2^ / R^2^ adjusted | 0.974 / 0.974 | | |

Table 5 shows that LE of the population of Cape Verde decreased in 1991 compared to 1990 and from 1993 till 1995. In all the remaining years till 2000, it increased significantly and attained a highest LE in 2017 (estimate). However, it decreased in 2018 and again increased in 2019 but decreased from 2020 to 2021 during the COVID-19 pandemic.

**Table 6: Comparison of LE_0_ between males and females in Cape Verde (1990-2021).**

|  | ***ex*** | | |
| --- | --- | --- | --- |
| ***Predictors*** | ***Estimates*** | ***CI*** | ***p*** |
| (Intercept) | 72.53 | 72.00 – 73.07 | **<0.001** |
| Year [1990] | -4.71 | -5.42 – -4.00 | **<0.001** |
| Year [1991] | -4.72 | -5.43 – -4.01 | **<0.001** |
| Year [1992] | -4.69 | -5.40 – -3.98 | **<0.001** |
| Year [1993] | -4.74 | -5.45 – -4.03 | **<0.001** |
| Year [1994] | -4.80 | -5.50 – -4.09 | **<0.001** |
| Year [1995] | -5.15 | -5.86 – -4.44 | **<0.001** |
| Year [1996] | -4.68 | -5.39 – -3.97 | **<0.001** |
| Year [1997] | -4.46 | -5.16 – -3.75 | **<0.001** |
| Year [1998] | -4.27 | -4.98 – -3.56 | **<0.001** |
| Year [1999] | -4.13 | -4.84 – -3.42 | **<0.001** |
| Year [2000] | -3.65 | -4.36 – -2.94 | **<0.001** |
| Year [2001] | -2.96 | -3.67 – -2.25 | **<0.001** |
| Year [2002] | -2.57 | -3.28 – -1.86 | **<0.001** |
| Year [2003] | -2.18 | -2.89 – -1.47 | **<0.001** |
| Year [2004] | -1.83 | -2.54 – -1.12 | **<0.001** |
| Year [2005] | -1.41 | -2.12 – -0.70 | **<0.001** |
| Year [2006] | -1.71 | -2.42 – -1.00 | **<0.001** |
| Year [2007] | -1.26 | -1.97 – -0.55 | **0.001** |
| Year [2008] | -1.57 | -2.27 – -0.86 | **<0.001** |
| Year [2009] | -1.07 | -1.78 – -0.36 | **0.003** |
| Year [2010] | -0.86 | -1.57 – -0.15 | **0.017** |
| Year [2011] | -0.42 | -1.13 – 0.29 | 0.245 |
| Year [2012] | -0.45 | -1.16 – 0.25 | 0.208 |
| Year [2013] | -0.07 | -0.78 – 0.64 | 0.848 |
| Year [2014] | -0.05 | -0.76 – 0.65 | 0.879 |
| Year [2015] | -0.58 | -1.29 – 0.13 | 0.109 |
| Year [2016] | 0.07 | -0.64 – 0.78 | 0.847 |
| Year [2017] | 0.57 | -0.14 – 1.27 | 0.118 |
| Year [2018] | -0.12 | -0.83 – 0.59 | 0.737 |
| Year [2020] | -0.98 | -1.69 – -0.28 | **0.006** |
| Year [2021] | -1.55 | -2.26 – -0.84 | **<0.001** |
| Age | -0.76 | -0.76 – -0.76 | **<0.001** |
| Gender [Male] | -4.25 | -4.42 – -4.07 | **<0.001** |
| Observations | 6464 | | |
| R^2^ / R^2^ adjusted | 0.974 / 0.974 | | |

Table 6 shows that compared to 2019, LE differences between males and females was at its highest in 2017 followed by 2016 and then 2019 (which posted positive estimates) but LE between males and females decreased in 2020 and continued decreasing in 2021.

**Table 7: Linear regression analysis table of LE_0_ in Cote d’Ivoire (1990-2021).**

|  | **ex** | | |
| --- | --- | --- | --- |
| **Predictors** | **Estimates** | **CI** | **p** |
| (Intercept) | 55.62 | 55.16 – 56.08 | **<0.001** |
| Year [1991] | -0.12 | -0.74 – 0.49 | 0.698 |
| Year [1992] | -0.20 | -0.82 – 0.42 | 0.527 |
| Year [1993] | -0.30 | -0.92 – 0.32 | 0.341 |
| Year [1994] | -0.39 | -1.00 – 0.23 | 0.217 |
| Year [1995] | -0.46 | -1.08 – 0.16 | 0.144 |
| Year [1996] | -0.55 | -1.16 – 0.07 | 0.082 |
| Year [1997] | -0.66 | -1.27 – -0.04 | **0.036** |
| Year [1998] | -0.93 | -1.55 – -0.32 | **0.003** |
| Year [1999] | -1.12 | -1.74 – -0.51 | **<0.001** |
| Year [2000] | -1.28 | -1.90 – -0.67 | **<0.001** |
| Year [2001] | -1.40 | -2.01 – -0.78 | **<0.001** |
| Year [2002] | -1.47 | -2.09 – -0.85 | **<0.001** |
| Year [2003] | -1.54 | -2.15 – -0.92 | **<0.001** |
| Year [2004] | -1.45 | -2.07 – -0.84 | **<0.001** |
| Year [2005] | -1.36 | -1.98 – -0.75 | **<0.001** |
| Year [2006] | -1.20 | -1.82 – -0.58 | **<0.001** |
| Year [2007] | -0.97 | -1.58 – -0.35 | **0.002** |
| Year [2008] | -0.73 | -1.35 – -0.12 | **0.020** |
| Year [2009] | -0.48 | -1.09 – 0.14 | 0.130 |
| Year [2010] | -0.27 | -0.88 – 0.35 | 0.397 |
| Year [2011] | -0.10 | -0.71 – 0.52 | 0.761 |
| Year [2012] | 0.13 | -0.49 – 0.74 | 0.683 |
| Year [2013] | 0.33 | -0.28 – 0.95 | 0.289 |
| Year [2014] | 0.46 | -0.16 – 1.07 | 0.145 |
| Year [2015] | 0.65 | 0.04 – 1.27 | **0.038** |
| Year [2016] | 0.73 | 0.12 – 1.35 | **0.020** |
| Year [2017] | 0.88 | 0.27 – 1.50 | **0.005** |
| Year [2018] | 0.96 | 0.34 – 1.57 | **0.002** |
| Year [2019] | 1.13 | 0.51 – 1.74 | **<0.001** |
| Year [2020] | 0.56 | -0.06 – 1.18 | 0.075 |
| Year [2021] | 0.08 | -0.53 – 0.70 | 0.790 |
| Age | -0.60 | -0.61 – -0.60 | **<0.001** |
| Gender [Male] | -1.23 | -1.38 – -1.07 | **<0.001** |
| Observations | 6464 | | |
| R^2^ / R^2^ adjusted | 0.969 / 0.969 | | |

Table 7 shows that LE decreased from 1991 till 2011 compared to 1990 and this decrease was statistically significant from 1997 to 2008. LE increased to 2012 after which it began increasing significantly from 2015 till 2019 where LE was at it highest value. In 2020 and 2021, LE decreased insignificantly to a minimum of 0.08.

**Table 8: Comparison of LE_0_ between males and females in Cote d’Ivoire (1990-2021).**

|  | **ex** | | |
| --- | --- | --- | --- |
| **Predictors** | **Estimates** | **CI** | **p** |
| (Intercept) | 56.75 | 56.29 – 57.21 | **<0.001** |
| Year [1990] | -1.13 | -1.74 – -0.51 | **<0.001** |
| Year [1991] | -1.25 | -1.87 – -0.63 | **<0.001** |
| Year [1992] | -1.33 | -1.94 – -0.71 | **<0.001** |
| Year [1993] | -1.43 | -2.04 – -0.81 | **<0.001** |
| Year [1994] | -1.52 | -2.13 – -0.90 | **<0.001** |
| Year [1995] | -1.59 | -2.20 – -0.97 | **<0.001** |
| Year [1996] | -1.67 | -2.29 – -1.06 | **<0.001** |
| Year [1997] | -1.79 | -2.40 – -1.17 | **<0.001** |
| Year [1998] | -2.06 | -2.68 – -1.45 | **<0.001** |
| Year [1999] | -2.25 | -2.87 – -1.64 | **<0.001** |
| Year [2000] | -2.41 | -3.03 – -1.80 | **<0.001** |
| Year [2001] | -2.53 | -3.14 – -1.91 | **<0.001** |
| Year [2002] | -2.60 | -3.22 – -1.98 | **<0.001** |
| Year [2003] | -2.66 | -3.28 – -2.05 | **<0.001** |
| Year [2004] | -2.58 | -3.20 – -1.96 | **<0.001** |
| Year [2005] | -2.49 | -3.11 – -1.88 | **<0.001** |
| Year [2006] | -2.33 | -2.95 – -1.71 | **<0.001** |
| Year [2007] | -2.10 | -2.71 – -1.48 | **<0.001** |
| Year [2008] | -1.86 | -2.48 – -1.24 | **<0.001** |
| Year [2009] | -1.60 | -2.22 – -0.99 | **<0.001** |
| Year [2010] | -1.39 | -2.01 – -0.78 | **<0.001** |
| Year [2011] | -1.22 | -1.84 – -0.61 | **<0.001** |
| Year [2012] | -1.00 | -1.62 – -0.38 | **0.001** |
| Year [2013] | -0.80 | -1.41 – -0.18 | **0.011** |
| Year [2014] | -0.67 | -1.29 – -0.05 | **0.033** |
| Year [2015] | -0.47 | -1.09 – 0.14 | 0.131 |
| Year [2016] | -0.40 | -1.01 – 0.22 | 0.208 |
| Year [2017] | -0.24 | -0.86 – 0.37 | 0.436 |
| Year [2018] | -0.17 | -0.79 – 0.45 | 0.588 |
| Year [2020] | -0.57 | -1.19 – 0.05 | 0.070 |
| Year [2021] | -1.04 | -1.66 – -0.43 | **0.001** |
| Age | -0.60 | -0.61 – -0.60 | **<0.001** |
| Gender [Male] | -1.23 | -1.38 – -1.07 | **<0.001** |
| Observations | 6464 | | |
| R^2^ / R^2^ adjusted | 0.969 / 0.969 | | |

Table 8 shows that compared with 2019 LE between males and females decreased in 2020 but decreased significantly in 2021.

**Table 9: Linear regression table of LE_0_ for Gambia (1990-2021).**

|  | **ex** | | |
| --- | --- | --- | --- |
| **Predictors** | **Estimates** | **CI** | **p** |
| (Intercept) | 58.91 | 58.46 – 59.35 | **<0.001** |
| Year [1991] | 0.24 | -0.35 – 0.84 | 0.425 |
| Year [1992] | 0.49 | -0.11 – 1.08 | 0.108 |
| Year [1993] | 0.73 | 0.14 – 1.32 | **0.016** |
| Year [1994] | 1.05 | 0.46 – 1.65 | **0.001** |
| Year [1995] | 1.26 | 0.67 – 1.85 | **<0.001** |
| Year [1996] | 1.37 | 0.77 – 1.96 | **<0.001** |
| Year [1997] | 1.27 | 0.67 – 1.86 | **<0.001** |
| Year [1998] | 1.33 | 0.73 – 1.92 | **<0.001** |
| Year [1999] | 1.33 | 0.73 – 1.92 | **<0.001** |
| Year [2000] | 1.23 | 0.64 – 1.82 | **<0.001** |
| Year [2001] | 1.14 | 0.55 – 1.74 | **<0.001** |
| Year [2002] | 1.02 | 0.43 – 1.62 | **0.001** |
| Year [2003] | 1.14 | 0.55 – 1.73 | **<0.001** |
| Year [2004] | 1.26 | 0.66 – 1.85 | **<0.001** |
| Year [2005] | 1.29 | 0.70 – 1.89 | **<0.001** |
| Year [2006] | 1.48 | 0.89 – 2.07 | **<0.001** |
| Year [2007] | 1.54 | 0.94 – 2.13 | **<0.001** |
| Year [2008] | 1.76 | 1.16 – 2.35 | **<0.001** |
| Year [2009] | 1.82 | 1.23 – 2.42 | **<0.001** |
| Year [2010] | 1.86 | 1.27 – 2.46 | **<0.001** |
| Year [2011] | 2.15 | 1.56 – 2.75 | **<0.001** |
| Year [2012] | 2.33 | 1.74 – 2.93 | **<0.001** |
| Year [2013] | 2.27 | 1.67 – 2.86 | **<0.001** |
| Year [2014] | 2.26 | 1.67 – 2.85 | **<0.001** |
| Year [2015] | 2.29 | 1.70 – 2.88 | **<0.001** |
| Year [2016] | 2.46 | 1.87 – 3.05 | **<0.001** |
| Year [2017] | 2.38 | 1.79 – 2.98 | **<0.001** |
| Year [2018] | 2.34 | 1.75 – 2.94 | **<0.001** |
| Year [2019] | 2.62 | 2.03 – 3.22 | **<0.001** |
| Year [2020] | 1.57 | 0.97 – 2.16 | **<0.001** |
| Year [2021] | 1.04 | 0.45 – 1.64 | **0.001** |
| Age | -0.66 | -0.67 – -0.66 | **<0.001** |
| Gender [Male] | -1.52 | -1.67 – -1.37 | **<0.001** |
| Observations | 6464 | | |
| R^2^ / R^2^ adjusted | 0.976 / 0.976 | | |

Table 9 shows that compared to 1990, LE increased in all the years and became significant between 1997 to 2021 with the highest value in 2019. There was a gradual decrease in LE in Gambia between 2020 to 2021 during the COVID-19 pandemic.

**Table 10: Comparison of LE_0_ between males and females in Gambia (1990-2021).**

|  | **ex** | | |
| --- | --- | --- | --- |
| **Predictors** | **Estimates** | **CI** | **p** |
| (Intercept) | 61.53 | 61.09 – 61.98 | **<0.001** |
| Year [1990] | -2.62 | -3.22 – -2.03 | **<0.001** |
| Year [1991] | -2.38 | -2.98 – -1.79 | **<0.001** |
| Year [1992] | -2.14 | -2.73 – -1.54 | **<0.001** |
| Year [1993] | -1.89 | -2.49 – -1.30 | **<0.001** |
| Year [1994] | -1.57 | -2.17 – -0.98 | **<0.001** |
| Year [1995] | -1.37 | -1.96 – -0.77 | **<0.001** |
| Year [1996] | -1.26 | -1.85 – -0.66 | **<0.001** |
| Year [1997] | -1.36 | -1.95 – -0.76 | **<0.001** |
| Year [1998] | -1.30 | -1.89 – -0.70 | **<0.001** |
| Year [1999] | -1.30 | -1.89 – -0.70 | **<0.001** |
| Year [2000] | -1.40 | -1.99 – -0.80 | **<0.001** |
| Year [2001] | -1.48 | -2.07 – -0.89 | **<0.001** |
| Year [2002] | -1.60 | -2.19 – -1.01 | **<0.001** |
| Year [2003] | -1.49 | -2.08 – -0.89 | **<0.001** |
| Year [2004] | -1.37 | -1.96 – -0.78 | **<0.001** |
| Year [2005] | -1.33 | -1.93 – -0.74 | **<0.001** |
| Year [2006] | -1.14 | -1.74 – -0.55 | **<0.001** |
| Year [2007] | -1.09 | -1.68 – -0.49 | **<0.001** |
| Year [2008] | -0.87 | -1.46 – -0.27 | **0.004** |
| Year [2009] | -0.80 | -1.40 – -0.21 | **0.008** |
| Year [2010] | -0.76 | -1.35 – -0.17 | **0.012** |
| Year [2011] | -0.47 | -1.06 – 0.12 | 0.120 |
| Year [2012] | -0.29 | -0.88 – 0.30 | 0.339 |
| Year [2013] | -0.36 | -0.95 – 0.23 | 0.236 |
| Year [2014] | -0.37 | -0.96 – 0.23 | 0.228 |
| Year [2015] | -0.33 | -0.93 – 0.26 | 0.269 |
| Year [2016] | -0.16 | -0.76 – 0.43 | 0.586 |
| Year [2017] | -0.24 | -0.83 – 0.35 | 0.429 |
| Year [2018] | -0.28 | -0.87 – 0.31 | 0.353 |
| Year [2020] | -1.06 | -1.65 – -0.47 | **<0.001** |
| Year [2021] | -1.58 | -2.17 – -0.99 | **<0.001** |
| Age | -0.66 | -0.67 – -0.66 | **<0.001** |
| Gender [Male] | -1.52 | -1.67 – -1.37 | **<0.001** |
| Observations | 6464 | | |
| R^2^ / R^2^ adjusted | 0.976 / 0.976 | | |

Table 10 shows that LE increased from 1990 to 2019 which had the highest value and thus explains the negative LE values observed for the years before 2019. Using 2019 as the reference year, LE in 2020 and 2021 decreased significantly.

**Table 11: Linear regression table of LE_0_ for Ghana (1990-2021).**

|  | **ex** | | |
| --- | --- | --- | --- |
| **Predictors** | **Estimates** | **CI** | **p** |
| (Intercept) | 60.16 | 59.72 – 60.61 | **<0.001** |
| Year [1991] | 0.07 | -0.52 – 0.67 | 0.804 |
| Year [1992] | 0.08 | -0.51 – 0.67 | 0.794 |
| Year [1993] | 0.10 | -0.49 – 0.70 | 0.731 |
| Year [1994] | -0.07 | -0.66 – 0.52 | 0.815 |
| Year [1995] | 0.08 | -0.51 – 0.68 | 0.784 |
| Year [1996] | 0.14 | -0.45 – 0.74 | 0.633 |
| Year [1997] | 0.26 | -0.33 – 0.86 | 0.384 |
| Year [1998] | 0.38 | -0.22 – 0.97 | 0.214 |
| Year [1999] | 0.48 | -0.11 – 1.08 | 0.111 |
| Year [2000] | 0.44 | -0.15 – 1.04 | 0.142 |
| Year [2001] | 0.28 | -0.31 – 0.88 | 0.350 |
| Year [2002] | 0.41 | -0.18 – 1.00 | 0.176 |
| Year [2003] | 0.55 | -0.05 – 1.14 | 0.070 |
| Year [2004] | 0.50 | -0.09 – 1.09 | 0.098 |
| Year [2005] | 0.69 | 0.10 – 1.29 | **0.022** |
| Year [2006] | 0.73 | 0.14 – 1.32 | **0.016** |
| Year [2007] | 0.77 | 0.17 – 1.36 | **0.011** |
| Year [2008] | 0.81 | 0.22 – 1.41 | **0.007** |
| Year [2009] | 0.94 | 0.35 – 1.54 | **0.002** |
| Year [2010] | 0.94 | 0.35 – 1.54 | **0.002** |
| Year [2011] | 1.08 | 0.49 – 1.67 | **<0.001** |
| Year [2012] | 1.18 | 0.59 – 1.78 | **<0.001** |
| Year [2013] | 1.25 | 0.66 – 1.85 | **<0.001** |
| Year [2014] | 1.47 | 0.88 – 2.07 | **<0.001** |
| Year [2015] | 1.46 | 0.86 – 2.05 | **<0.001** |
| Year [2016] | 1.72 | 1.13 – 2.31 | **<0.001** |
| Year [2017] | 1.71 | 1.12 – 2.31 | **<0.001** |
| Year [2018] | 1.71 | 1.11 – 2.30 | **<0.001** |
| Year [2019] | 1.95 | 1.35 – 2.54 | **<0.001** |
| Year [2020] | 1.35 | 0.75 – 1.94 | **<0.001** |
| Year [2021] | 1.00 | 0.41 – 1.60 | **0.001** |
| Age | -0.67 | -0.67 – -0.67 | **<0.001** |
| Gender [Male] | -1.35 | -1.50 – -1.20 | **<0.001** |
| Observations | 6464 | | |
| R^2^ / R^2^ adjusted | 0.977 / 0.976 | | |

Table 11 shows that compared to 1990, LE increased up to 1993 but decreased in 1994. LE in Ghana increased significantly from 2005 until it attained the highest value in 2019. However, LE significantly decreased in 2020 and 2021.

**Table 12: Comparison of LE_0_ between males and females in Ghana (1990-2021).**

|  | **ex** | | |
| --- | --- | --- | --- |
| **Predictors** | **Estimates** | **CI** | **p** |
| (Intercept) | 62.11 | 61.67 – 62.55 | **<0.001** |
| Year [1990] | -1.95 | -2.54 – -1.35 | **<0.001** |
| Year [1991] | -1.87 | -2.46 – -1.28 | **<0.001** |
| Year [1992] | -1.87 | -2.46 – -1.27 | **<0.001** |
| Year [1993] | -1.84 | -2.44 – -1.25 | **<0.001** |
| Year [1994] | -2.02 | -2.61 – -1.42 | **<0.001** |
| Year [1995] | -1.86 | -2.46 – -1.27 | **<0.001** |
| Year [1996] | -1.80 | -2.39 – -1.21 | **<0.001** |
| Year [1997] | -1.68 | -2.28 – -1.09 | **<0.001** |
| Year [1998] | -1.57 | -2.16 – -0.98 | **<0.001** |
| Year [1999] | -1.46 | -2.06 – -0.87 | **<0.001** |
| Year [2000] | -1.50 | -2.10 – -0.91 | **<0.001** |
| Year [2001] | -1.66 | -2.26 – -1.07 | **<0.001** |
| Year [2002] | -1.54 | -2.13 – -0.94 | **<0.001** |
| Year [2003] | -1.40 | -1.99 – -0.80 | **<0.001** |
| Year [2004] | -1.44 | -2.04 – -0.85 | **<0.001** |
| Year [2005] | -1.25 | -1.85 – -0.66 | **<0.001** |
| Year [2006] | -1.22 | -1.81 – -0.62 | **<0.001** |
| Year [2007] | -1.18 | -1.77 – -0.59 | **<0.001** |
| Year [2008] | -1.13 | -1.73 – -0.54 | **<0.001** |
| Year [2009] | -1.00 | -1.60 – -0.41 | **0.001** |
| Year [2010] | -1.00 | -1.59 – -0.41 | **0.001** |
| Year [2011] | -0.87 | -1.46 – -0.27 | **0.004** |
| Year [2012] | -0.76 | -1.35 – -0.17 | **0.012** |
| Year [2013] | -0.69 | -1.29 – -0.10 | **0.022** |
| Year [2014] | -0.47 | -1.07 – 0.12 | 0.119 |
| Year [2015] | -0.49 | -1.08 – 0.10 | 0.105 |
| Year [2016] | -0.23 | -0.82 – 0.37 | 0.455 |
| Year [2017] | -0.23 | -0.83 – 0.36 | 0.440 |
| Year [2018] | -0.24 | -0.83 – 0.35 | 0.429 |
| Year [2020] | -0.60 | -1.19 – -0.01 | **0.047** |
| Year [2021] | -0.94 | -1.53 – -0.35 | **0.002** |
| Age | -0.67 | -0.67 – -0.67 | **<0.001** |
| Gender [Male] | -1.35 | -1.50 – -1.20 | **<0.001** |
| Observations | 6464 | | |
| R^2^ / R^2^ adjusted | 0.977 / 0.976 | | |

Table 12 shows that compared with 2019, LE between males and females were significantly different, except between 2014 to 2018. Also, LE_0_ between females and males decreased significantly in 2020 and 2021 (during the COVID-19 pandemic).

**Table 13: Linear regression table of LE_0_ for Guinea (1990-2021).**

|  | **ex** | | |
| --- | --- | --- | --- |
| **Predictors** | **Estimates** | **CI** | **p** |
| (Intercept) | 57.51 | 57.07 – 57.94 | **<0.001** |
| Year [1991] | 0.10 | -0.48 – 0.68 | 0.731 |
| Year [1992] | 0.33 | -0.25 – 0.92 | 0.259 |
| Year [1993] | 0.44 | -0.14 – 1.02 | 0.135 |
| Year [1994] | 0.37 | -0.21 – 0.95 | 0.216 |
| Year [1995] | 0.51 | -0.07 – 1.09 | 0.083 |
| Year [1996] | 0.63 | 0.05 – 1.21 | **0.032** |
| Year [1997] | 0.84 | 0.26 – 1.42 | **0.005** |
| Year [1998] | 0.99 | 0.41 – 1.57 | **0.001** |
| Year [1999] | 0.94 | 0.36 – 1.52 | **0.001** |
| Year [2000] | 0.79 | 0.21 – 1.37 | **0.008** |
| Year [2001] | 0.69 | 0.11 – 1.27 | **0.020** |
| Year [2002] | 0.79 | 0.21 – 1.37 | **0.008** |
| Year [2003] | 0.81 | 0.23 – 1.39 | **0.006** |
| Year [2004] | 0.87 | 0.29 – 1.45 | **0.003** |
| Year [2005] | 0.97 | 0.39 – 1.55 | **0.001** |
| Year [2006] | 1.08 | 0.50 – 1.66 | **<0.001** |
| Year [2007] | 1.19 | 0.61 – 1.77 | **<0.001** |
| Year [2008] | 1.32 | 0.73 – 1.90 | **<0.001** |
| Year [2009] | 1.38 | 0.80 – 1.96 | **<0.001** |
| Year [2010] | 1.48 | 0.90 – 2.06 | **<0.001** |
| Year [2011] | 1.55 | 0.97 – 2.13 | **<0.001** |
| Year [2012] | 1.67 | 1.09 – 2.25 | **<0.001** |
| Year [2013] | 1.80 | 1.22 – 2.38 | **<0.001** |
| Year [2014] | 1.80 | 1.22 – 2.38 | **<0.001** |
| Year [2015] | 1.87 | 1.29 – 2.45 | **<0.001** |
| Year [2016] | 2.08 | 1.50 – 2.66 | **<0.001** |
| Year [2017] | 2.19 | 1.61 – 2.77 | **<0.001** |
| Year [2018] | 2.24 | 1.66 – 2.82 | **<0.001** |
| Year [2019] | 2.35 | 1.76 – 2.93 | **<0.001** |
| Year [2020] | 1.69 | 1.11 – 2.27 | **<0.001** |
| Year [2021] | 1.20 | 0.61 – 1.78 | **<0.001** |
| Age | -0.65 | -0.65 – -0.64 | **<0.001** |
| Gender [Male] | -0.80 | -0.95 – -0.66 | **<0.001** |
| Observations | 6464 | | |
| R^2^ / R^2^ adjusted | 0.976 / 0.976 | | |

Table 13 shows that in the last 30 years LE_0_ in Guinea increased compared to the 1990 value and it became significant in 1996 reaching the highest value in 2019. Contrary to findings in other West African Countries, LE in Guinea population significantly increased 2020 and insignificantly in 2021.

**Table 14: Comparison of LE_0_ between males and females in Guinea (1990-2021).**

|  | **ex** | | |
| --- | --- | --- | --- |
| **Predictors** | **Estimates** | **CI** | **p** |
| (Intercept) | 59.86 | 59.42 – 60.29 | **<0.001** |
| Year [1990] | -2.35 | -2.93 – -1.76 | **<0.001** |
| Year [1991] | -2.24 | -2.82 – -1.66 | **<0.001** |
| Year [1992] | -2.01 | -2.59 – -1.43 | **<0.001** |
| Year [1993] | -1.90 | -2.48 – -1.32 | **<0.001** |
| Year [1994] | -1.98 | -2.56 – -1.40 | **<0.001** |
| Year [1995] | -1.83 | -2.41 – -1.25 | **<0.001** |
| Year [1996] | -1.71 | -2.29 – -1.13 | **<0.001** |
| Year [1997] | -1.50 | -2.09 – -0.92 | **<0.001** |
| Year [1998] | -1.35 | -1.93 – -0.77 | **<0.001** |
| Year [1999] | -1.40 | -1.98 – -0.82 | **<0.001** |
| Year [2000] | -1.55 | -2.13 – -0.97 | **<0.001** |
| Year [2001] | -1.66 | -2.24 – -1.08 | **<0.001** |
| Year [2002] | -1.55 | -2.13 – -0.97 | **<0.001** |
| Year [2003] | -1.54 | -2.12 – -0.96 | **<0.001** |
| Year [2004] | -1.48 | -2.06 – -0.90 | **<0.001** |
| Year [2005] | -1.37 | -1.95 – -0.79 | **<0.001** |
| Year [2006] | -1.26 | -1.84 – -0.68 | **<0.001** |
| Year [2007] | -1.16 | -1.74 – -0.58 | **<0.001** |
| Year [2008] | -1.03 | -1.61 – -0.45 | **0.001** |
| Year [2009] | -0.96 | -1.54 – -0.38 | **0.001** |
| Year [2010] | -0.87 | -1.45 – -0.28 | **0.003** |
| Year [2011] | -0.79 | -1.37 – -0.21 | **0.007** |
| Year [2012] | -0.67 | -1.25 – -0.09 | **0.023** |
| Year [2013] | -0.55 | -1.13 – 0.03 | 0.064 |
| Year [2014] | -0.54 | -1.12 – 0.04 | 0.067 |
| Year [2015] | -0.47 | -1.05 – 0.11 | 0.111 |
| Year [2016] | -0.26 | -0.84 – 0.32 | 0.376 |
| Year [2017] | -0.16 | -0.74 – 0.42 | 0.592 |
| Year [2018] | -0.10 | -0.68 – 0.48 | 0.726 |
| Year [2020] | -0.66 | -1.24 – -0.08 | **0.026** |
| Year [2021] | -1.15 | -1.73 – -0.57 | **<0.001** |
| Age | -0.65 | -0.65 – -0.64 | **<0.001** |
| sex [Male] | -0.80 | -0.95 – -0.66 | **<0.001** |
| Observations | 6464 | | |
| R^2^ / R^2^ adjusted | 0.976 / 0.976 | | |

Table 14 shows that compared to 2019, LE_0_ between males and females increased significantly up to 2012 but significantly decreased in 2020 and 2021.

**Table 15: Linear regression table of LE_0_ for Guinea-Bissau (1990-2021).**

|  | **ex** | | |
| --- | --- | --- | --- |
| **Predictors** | **Estimates** | **CI** | **p** |
| (Intercept) | 56.71 | 56.23 – 57.19 | **<0.001** |
| Year [1991] | 0.05 | -0.58 – 0.69 | 0.867 |
| Year [1992] | 0.15 | -0.49 – 0.78 | 0.649 |
| Year [1993] | 0.13 | -0.50 – 0.77 | 0.683 |
| Year [1994] | -0.01 | -0.64 – 0.63 | 0.980 |
| Year [1995] | 0.16 | -0.47 – 0.80 | 0.617 |
| Year [1996] | 0.03 | -0.60 – 0.67 | 0.925 |
| Year [1997] | -0.56 | -1.19 – 0.08 | 0.086 |
| Year [1998] | -1.02 | -1.65 – -0.38 | **0.002** |
| Year [1999] | -1.00 | -1.63 – -0.36 | **0.002** |
| Year [2000] | -0.24 | -0.88 – 0.39 | 0.454 |
| Year [2001] | -0.28 | -0.92 – 0.35 | 0.382 |
| Year [2002] | -0.20 | -0.84 – 0.44 | 0.538 |
| Year [2003] | -0.21 | -0.85 – 0.42 | 0.509 |
| Year [2004] | -0.02 | -0.66 – 0.61 | 0.941 |
| Year [2005] | -0.09 | -0.73 – 0.54 | 0.772 |
| Year [2006] | 0.20 | -0.44 – 0.83 | 0.547 |
| Year [2007] | 0.38 | -0.25 – 1.02 | 0.236 |
| Year [2008] | 0.49 | -0.15 – 1.12 | 0.132 |
| Year [2009] | 0.67 | 0.04 – 1.31 | **0.038** |
| Year [2010] | 0.82 | 0.19 – 1.46 | **0.011** |
| Year [2011] | 1.02 | 0.39 – 1.66 | **0.002** |
| Year [2012] | 1.23 | 0.60 – 1.87 | **<0.001** |
| Year [2013] | 1.34 | 0.71 – 1.98 | **<0.001** |
| Year [2014] | 1.37 | 0.74 – 2.01 | **<0.001** |
| Year [2015] | 1.51 | 0.87 – 2.14 | **<0.001** |
| Year [2016] | 1.67 | 1.04 – 2.31 | **<0.001** |
| Year [2017] | 1.75 | 1.11 – 2.39 | **<0.001** |
| Year [2018] | 1.87 | 1.23 – 2.50 | **<0.001** |
| Year [2019] | 1.95 | 1.32 – 2.59 | **<0.001** |
| Year [2020] | 0.95 | 0.31 – 1.59 | **0.003** |
| Year [2021] | 0.56 | -0.07 – 1.20 | 0.082 |
| Age | -0.62 | -0.63 – -0.62 | **<0.001** |
| Gender [Male] | -2.05 | -2.21 – -1.89 | **<0.001** |
| Observations | 6464 | | |
| R^2^ / R^2^ adjusted | 0.969 / 0.969 | | |

Table 15 shows that compared to 1990, LE_0_ in Guinea-Bissau, decreased in 1994 and between 1997 to 2005 and was significant between 1998-1999. LE_0_ also increased in 2006 till 2021 and was significant in 2009. The highest value of life expectancy was in 2019. In 2020 and 2021, LE_0_ decreased compared to the 1990 value.

**Table 16: Comparison of LE_0_ between males and females in Guinea-Bissau (1990-2021).**

|  | **ex** | | |
| --- | --- | --- | --- |
| **Predictors** | **Estimates** | **CI** | **p** |
| (Intercept) | 58.66 | 58.19 – 59.14 | **<0.001** |
| Year [1990] | -1.95 | -2.59 – -1.32 | **<0.001** |
| Year [1991] | -1.90 | -2.54 – -1.26 | **<0.001** |
| Year [1992] | -1.81 | -2.44 – -1.17 | **<0.001** |
| Year [1993] | -1.82 | -2.46 – -1.19 | **<0.001** |
| Year [1994] | -1.96 | -2.60 – -1.33 | **<0.001** |
| Year [1995] | -1.79 | -2.43 – -1.16 | **<0.001** |
| Year [1996] | -1.92 | -2.56 – -1.29 | **<0.001** |
| Year [1997] | -2.51 | -3.15 – -1.88 | **<0.001** |
| Year [1998] | -2.97 | -3.61 – -2.34 | **<0.001** |
| Year [1999] | -2.95 | -3.59 – -2.32 | **<0.001** |
| Year [2000] | -2.20 | -2.83 – -1.56 | **<0.001** |
| Year [2001] | -2.24 | -2.87 – -1.60 | **<0.001** |
| Year [2002] | -2.15 | -2.79 – -1.52 | **<0.001** |
| Year [2003] | -2.17 | -2.80 – -1.53 | **<0.001** |
| Year [2004] | -1.98 | -2.61 – -1.34 | **<0.001** |
| Year [2005] | -2.05 | -2.68 – -1.41 | **<0.001** |
| Year [2006] | -1.76 | -2.39 – -1.12 | **<0.001** |
| Year [2007] | -1.57 | -2.21 – -0.93 | **<0.001** |
| Year [2008] | -1.47 | -2.10 – -0.83 | **<0.001** |
| Year [2009] | -1.28 | -1.92 – -0.65 | **<0.001** |
| Year [2010] | -1.13 | -1.76 – -0.49 | **<0.001** |
| Year [2011] | -0.93 | -1.57 – -0.30 | **0.004** |
| Year [2012] | -0.72 | -1.36 – -0.09 | **0.026** |
| Year [2013] | -0.61 | -1.25 – 0.02 | 0.059 |
| Year [2014] | -0.58 | -1.22 – 0.05 | 0.073 |
| Year [2015] | -0.45 | -1.08 – 0.19 | 0.168 |
| Year [2016] | -0.28 | -0.92 – 0.35 | 0.386 |
| Year [2017] | -0.20 | -0.84 – 0.43 | 0.528 |
| Year [2018] | -0.09 | -0.72 – 0.55 | 0.793 |
| Year [2020] | -1.00 | -1.64 – -0.37 | **0.002** |
| Year [2021] | -1.39 | -2.03 – -0.76 | **<0.001** |
| Age | -0.62 | -0.63 – -0.62 | **<0.001** |
| Gender [Male] | -2.05 | -2.21 – -1.89 | **<0.001** |
| Observations | 6464 | | |
| R^2^ / R^2^ adjusted | 0.969 / 0.969 | | |

Table 16 shows that LE_0_ in 2020 and 2021 decreased compared to the value in 2019 with 2021 decreasing more than 2020 (which had a higher LE_0_ estimate between males and females).

**Table 17: Linear regression table of LE_0_ for Liberia (1990-2021).**

|  | **ex** | | |
| --- | --- | --- | --- |
| **Predictors** | **Estimates** | **CI** | **p** |
| (Intercept) | 53.08 | 52.55 – 53.60 | **<0.001** |
| Year [1991] | 2.95 | 2.24 – 3.65 | **<0.001** |
| Year [1992] | 2.92 | 2.21 – 3.62 | **<0.001** |
| Year [1993] | 2.67 | 1.97 – 3.37 | **<0.001** |
| Year [1994] | 2.40 | 1.69 – 3.10 | **<0.001** |
| Year [1995] | 3.16 | 2.46 – 3.87 | **<0.001** |
| Year [1996] | 3.18 | 2.47 – 3.88 | **<0.001** |
| Year [1997] | 4.40 | 3.70 – 5.11 | **<0.001** |
| Year [1998] | 4.56 | 3.86 – 5.26 | **<0.001** |
| Year [1999] | 4.71 | 4.01 – 5.41 | **<0.001** |
| Year [2000] | 4.97 | 4.27 – 5.67 | **<0.001** |
| Year [2001] | 5.24 | 4.53 – 5.94 | **<0.001** |
| Year [2002] | 5.29 | 4.59 – 6.00 | **<0.001** |
| Year [2003] | 4.71 | 4.00 – 5.41 | **<0.001** |
| Year [2004] | 5.56 | 4.86 – 6.27 | **<0.001** |
| Year [2005] | 5.65 | 4.95 – 6.36 | **<0.001** |
| Year [2006] | 5.69 | 4.99 – 6.39 | **<0.001** |
| Year [2007] | 5.77 | 5.07 – 6.47 | **<0.001** |
| Year [2008] | 5.80 | 5.10 – 6.50 | **<0.001** |
| Year [2009] | 5.85 | 5.14 – 6.55 | **<0.001** |
| Year [2010] | 5.83 | 5.12 – 6.53 | **<0.001** |
| Year [2011] | 5.81 | 5.10 – 6.51 | **<0.001** |
| Year [2012] | 5.87 | 5.16 – 6.57 | **<0.001** |
| Year [2013] | 5.80 | 5.10 – 6.51 | **<0.001** |
| Year [2014] | 5.51 | 4.81 – 6.21 | **<0.001** |
| Year [2015] | 5.48 | 4.77 – 6.18 | **<0.001** |
| Year [2016] | 5.85 | 5.15 – 6.56 | **<0.001** |
| Year [2017] | 5.84 | 5.13 – 6.54 | **<0.001** |
| Year [2018] | 5.93 | 5.23 – 6.63 | **<0.001** |
| Year [2019] | 5.99 | 5.29 – 6.69 | **<0.001** |
| Year [2020] | 5.56 | 4.86 – 6.27 | **<0.001** |
| Year [2021] | 5.26 | 4.56 – 5.97 | **<0.001** |
| Age | -0.64 | -0.64 – -0.64 | **<0.001** |
| Gender [Male] | -1.52 | -1.70 – -1.35 | **<0.001** |
| Observations | 6464 | | |
| R^2^ / R^2^ adjusted | 0.964 / 0.964 | | |

Table 18 shows that compared to 1990, LE_0_ value in Liberia increased significantly compared to the other years(p-value<0.05) and the highest LE_0_ value was reached in 2019. Nevertheless, LE_0_ in Liberia also increased in 2020 and 2021.

**Table 18: Comparison of LE_0_ between males and females in Liberia (1990-2021).**

|  | **ex** | | |
| --- | --- | --- | --- |
| **Predictors** | **Estimates** | **CI** | **p** |
| (Intercept) | 59.07 | 58.54 – 59.60 | **<0.001** |
| Year [1990] | -5.99 | -6.69 – -5.29 | **<0.001** |
| Year [1991] | -3.04 | -3.75 – -2.34 | **<0.001** |
| Year [1992] | -3.08 | -3.78 – -2.37 | **<0.001** |
| Year [1993] | -3.32 | -4.03 – -2.62 | **<0.001** |
| Year [1994] | -3.60 | -4.30 – -2.89 | **<0.001** |
| Year [1995] | -2.83 | -3.53 – -2.12 | **<0.001** |
| Year [1996] | -2.81 | -3.52 – -2.11 | **<0.001** |
| Year [1997] | -1.59 | -2.29 – -0.88 | **<0.001** |
| Year [1998] | -1.43 | -2.13 – -0.73 | **<0.001** |
| Year [1999] | -1.28 | -1.98 – -0.58 | **<0.001** |
| Year [2000] | -1.02 | -1.72 – -0.32 | **0.005** |
| Year [2001] | -0.75 | -1.46 – -0.05 | **0.036** |
| Year [2002] | -0.70 | -1.40 – 0.01 | 0.052 |
| Year [2003] | -1.28 | -1.99 – -0.58 | **<0.001** |
| Year [2004] | -0.43 | -1.13 – 0.28 | 0.233 |
| Year [2005] | -0.34 | -1.04 – 0.37 | 0.346 |
| Year [2006] | -0.30 | -1.00 – 0.40 | 0.403 |
| Year [2007] | -0.22 | -0.93 – 0.48 | 0.538 |
| Year [2008] | -0.19 | -0.90 – 0.51 | 0.595 |
| Year [2009] | -0.14 | -0.85 – 0.56 | 0.688 |
| Year [2010] | -0.16 | -0.87 – 0.54 | 0.650 |
| Year [2011] | -0.18 | -0.89 – 0.52 | 0.610 |
| Year [2012] | -0.12 | -0.83 – 0.58 | 0.733 |
| Year [2013] | -0.19 | -0.89 – 0.52 | 0.599 |
| Year [2014] | -0.48 | -1.18 – 0.22 | 0.181 |
| Year [2015] | -0.51 | -1.22 – 0.19 | 0.154 |
| Year [2016] | -0.14 | -0.84 – 0.57 | 0.704 |
| Year [2017] | -0.15 | -0.86 – 0.55 | 0.668 |
| Year [2018] | -0.06 | -0.77 – 0.64 | 0.865 |
| Year [2020] | -0.43 | -1.13 – 0.28 | 0.234 |
| Year [2021] | -0.73 | -1.43 – -0.03 | **0.042** |
| age | -0.64 | -0.64 – -0.64 | **<0.001** |
| Gender [Male] | -1.52 | -1.70 – -1.35 | **<0.001** |
| Observations | 6464 | | |
| R^2^ / R^2^ adjusted | 0.964 / 0.964 | | |

Table 18 shows that in 2020 LE_0_ insignificantly decreased compared to 2019 and significantly decreased in 2021. Before 2019, LE_0_ in Liberia increased gradually to the highest value in 2019.

**Table 19: Linear regression table of LE_0_ for Mali (1990-2021).**

|  | **ex** | | |
| --- | --- | --- | --- |
| *Predictors* | *Estimates* | *CI* | *p* |
| (Intercept) | 56.63 | 56.15 – 57.11 | **<0.001** |
| Year [1991] | 0.17 | -0.47 – 0.82 | 0.598 |
| Year [1992] | 0.19 | -0.45 – 0.84 | 0.552 |
| Year [1993] | 0.21 | -0.43 – 0.86 | 0.512 |
| Year [1994] | 0.12 | -0.52 – 0.77 | 0.706 |
| Year [1995] | 0.05 | -0.59 – 0.69 | 0.882 |
| Year [1996] | -0.06 | -0.71 – 0.58 | 0.847 |
| Year [1997] | 0.17 | -0.47 – 0.82 | 0.597 |
| Year [1998] | 0.36 | -0.28 – 1.00 | 0.274 |
| Year [1999] | 0.53 | -0.11 – 1.17 | 0.106 |
| Year [2000] | 0.73 | 0.09 – 1.38 | **0.025** |
| Year [2001] | 0.92 | 0.28 – 1.57 | **0.005** |
| Year [2002] | 1.20 | 0.56 – 1.85 | **<0.001** |
| Year [2003] | 1.35 | 0.71 – 2.00 | **<0.001** |
| Year [2004] | 1.50 | 0.85 – 2.14 | **<0.001** |
| Year [2005] | 1.55 | 0.91 – 2.19 | **<0.001** |
| Year [2006] | 1.69 | 1.04 – 2.33 | **<0.001** |
| Year [2007] | 1.68 | 1.04 – 2.33 | **<0.001** |
| Year [2008] | 1.70 | 1.06 – 2.34 | **<0.001** |
| Year [2009] | 1.84 | 1.20 – 2.48 | **<0.001** |
| Year [2010] | 1.94 | 1.30 – 2.59 | **<0.001** |
| Year [2011] | 1.99 | 1.34 – 2.63 | **<0.001** |
| Year [2012] | 2.01 | 1.37 – 2.65 | **<0.001** |
| Year [2013] | 2.02 | 1.38 – 2.67 | **<0.001** |
| Year [2014] | 2.15 | 1.51 – 2.79 | **<0.001** |
| Year [2015] | 2.26 | 1.61 – 2.90 | **<0.001** |
| Year [2016] | 2.32 | 1.67 – 2.96 | **<0.001** |
| Year [2017] | 2.41 | 1.77 – 3.05 | **<0.001** |
| Year [2018] | 2.45 | 1.80 – 3.09 | **<0.001** |
| Year [2019] | 2.48 | 1.84 – 3.12 | **<0.001** |
| Year [2020] | 1.42 | 0.77 – 2.06 | **<0.001** |
| Year [2021] | 1.42 | 0.77 – 2.06 | **<0.001** |
| age | -0.65 | -0.65 – -0.65 | **<0.001** |
| Gender [Male] | -0.80 | -0.96 – -0.64 | **<0.001** |
| Observations | 6464 | | |
| R^2^ / R^2^ adjusted | 0.971 / 0.971 | | |

Table 19 shows that in Mali, LE_0_ progressively increased in the other years compared to 1990 except for 1996. It then began to take a significant increase from 2000 to 2019 was had the highest value. LE_0_ significantly decreased from 2020 to 2021 compared to 1990.

**Table 20: Comparison of LE_0_ between males and females in Mali (1990-2021).**

|  | **ex** | | |
| --- | --- | --- | --- |
| **Predictors** | **Estimates** | **CI** | **p** |
| (Intercept) | 59.11 | 58.63 – 59.59 | **<0.001** |
| Year [1990] | -2.48 | -3.12 – -1.84 | **<0.001** |
| Year [1991] | -2.31 | -2.95 – -1.66 | **<0.001** |
| Year [1992] | -2.28 | -2.93 – -1.64 | **<0.001** |
| Year [1993] | -2.26 | -2.91 – -1.62 | **<0.001** |
| Year [1994] | -2.36 | -3.00 – -1.71 | **<0.001** |
| Year [1995] | -2.43 | -3.07 – -1.79 | **<0.001** |
| Year [1996] | -2.54 | -3.18 – -1.90 | **<0.001** |
| Year [1997] | -2.31 | -2.95 – -1.66 | **<0.001** |
| Year [1998] | -2.12 | -2.76 – -1.48 | **<0.001** |
| Year [1999] | -1.95 | -2.59 – -1.31 | **<0.001** |
| Year [2000] | -1.74 | -2.39 – -1.10 | **<0.001** |
| Year [2001] | -1.56 | -2.20 – -0.91 | **<0.001** |
| Year [2002] | -1.27 | -1.92 – -0.63 | **<0.001** |
| Year [2003] | -1.12 | -1.77 – -0.48 | **0.001** |
| Year [2004] | -0.98 | -1.63 – -0.34 | **0.003** |
| Year [2005] | -0.93 | -1.57 – -0.29 | **0.005** |
| Year [2006] | -0.79 | -1.44 – -0.15 | **0.015** |
| Year [2007] | -0.80 | -1.44 – -0.15 | **0.015** |
| Year [2008] | -0.78 | -1.42 – -0.14 | **0.018** |
| Year [2009] | -0.64 | -1.28 – 0.00 | 0.051 |
| Year [2010] | -0.54 | -1.18 – 0.11 | 0.102 |
| Year [2011] | -0.49 | -1.14 – 0.15 | 0.132 |
| Year [2012] | -0.47 | -1.11 – 0.17 | 0.154 |
| Year [2013] | -0.46 | -1.10 – 0.19 | 0.164 |
| Year [2014] | -0.33 | -0.97 – 0.31 | 0.316 |
| Year [2015] | -0.22 | -0.87 – 0.42 | 0.495 |
| Year [2016] | -0.16 | -0.81 – 0.48 | 0.619 |
| Year [2017] | -0.07 | -0.71 – 0.57 | 0.833 |
| Year [2018] | -0.03 | -0.67 – 0.61 | 0.921 |
| Year [2020] | -1.06 | -1.71 – -0.42 | **0.001** |
| Year [2021] | -1.06 | -1.71 – -0.42 | **0.001** |
| age | -0.65 | -0.65 – -0.65 | **<0.001** |
| Gender [Male] | -0.80 | -0.96 – -0.64 | **<0.001** |
| Observations | 6464 | | |
| R^2^ / R^2^ adjusted | 0.971 / 0.971 | | |

Table 20 shows that compared to 2019, the LE_0_ in Mali significantly decreased in 2020 and 2021.

**Table 21: Linear regression table of LE_0_ for Mauritania (1990-2021).**

|  | **ex** | | |
| --- | --- | --- | --- |
| **Predictors** | **Estimates** | **CI** | **p** |
| (Intercept) | 62.44 | 61.91 – 62.98 | **<0.001** |
| Year [1991] | 0.03 | -0.69 – 0.74 | 0.945 |
| Year [1992] | 0.15 | -0.57 – 0.86 | 0.688 |
| Year [1993] | 0.18 | -0.53 – 0.90 | 0.615 |
| Year [1994] | 0.15 | -0.57 – 0.86 | 0.684 |
| Year [1995] | 0.11 | -0.61 – 0.82 | 0.766 |
| Year [1996] | 0.04 | -0.68 – 0.75 | 0.922 |
| Year [1997] | 0.21 | -0.51 – 0.92 | 0.572 |
| Year [1998] | 0.28 | -0.44 – 0.99 | 0.446 |
| Year [1999] | 0.37 | -0.34 – 1.09 | 0.304 |
| Year [2000] | 0.47 | -0.25 – 1.18 | 0.200 |
| Year [2001] | 0.53 | -0.18 – 1.24 | 0.145 |
| Year [2002] | 0.56 | -0.15 – 1.28 | 0.123 |
| Year [2003] | 0.58 | -0.13 – 1.29 | 0.112 |
| Year [2004] | 0.66 | -0.06 – 1.37 | 0.071 |
| Year [2005] | 0.65 | -0.06 – 1.37 | 0.073 |
| Year [2006] | 0.69 | -0.02 – 1.41 | 0.057 |
| Year [2007] | 0.79 | 0.08 – 1.51 | **0.029** |
| Year [2008] | 0.80 | 0.08 – 1.51 | **0.029** |
| Year [2009] | 0.92 | 0.21 – 1.63 | **0.011** |
| Year [2010] | 1.00 | 0.28 – 1.71 | **0.006** |
| Year [2011] | 1.04 | 0.33 – 1.75 | **0.004** |
| Year [2012] | 1.09 | 0.38 – 1.80 | **0.003** |
| Year [2013] | 1.18 | 0.47 – 1.90 | **0.001** |
| Year [2014] | 1.22 | 0.51 – 1.94 | **0.001** |
| Year [2015] | 1.31 | 0.60 – 2.03 | **<0.001** |
| Year [2016] | 1.38 | 0.67 – 2.10 | **<0.001** |
| Year [2017] | 1.47 | 0.75 – 2.18 | **<0.001** |
| Year [2018] | 1.51 | 0.79 – 2.22 | **<0.001** |
| Year [2019] | 1.63 | 0.91 – 2.34 | **<0.001** |
| Year [2020] | 0.54 | -0.17 – 1.26 | 0.136 |
| Year [2021] | 0.32 | -0.39 – 1.03 | 0.378 |
| age | -0.70 | -0.70 – -0.70 | **<0.001** |
| Gender [Male] | -1.70 | -1.88 – -1.53 | **<0.001** |
| Observations | 6464 | | |
| R^2^ / R^2^ adjusted | 0.969 / 0.969 | | |

Table 21 shows that in the 30 years, LE_0_ increased in all the years compared to 1990 and it became significant in 2007 and the highest value was obtained in 2019. In 2021 and 2021, LE_0_ were lower than 2019.

**Table 22: Comparison of LE_0_ between males and females in Mauritania (1990-2021).**

|  | **ex** | | |
| --- | --- | --- | --- |
| **Predictors** | **Estimates** | **CI** | **p** |
| (Intercept) | 64.07 | 63.53 – 64.60 | **<0.001** |
| Year [1990] | -1.63 | -2.34 – -0.91 | **<0.001** |
| Year [1991] | -1.60 | -2.31 – -0.89 | **<0.001** |
| Year [1992] | -1.48 | -2.19 – -0.77 | **<0.001** |
| Year [1993] | -1.44 | -2.16 – -0.73 | **<0.001** |
| Year [1994] | -1.48 | -2.19 – -0.76 | **<0.001** |
| Year [1995] | -1.52 | -2.23 – -0.80 | **<0.001** |
| Year [1996] | -1.59 | -2.30 – -0.88 | **<0.001** |
| Year [1997] | -1.42 | -2.13 – -0.71 | **<0.001** |
| Year [1998] | -1.35 | -2.06 – -0.63 | **<0.001** |
| Year [1999] | -1.25 | -1.96 – -0.54 | **0.001** |
| Year [2000] | -1.16 | -1.87 – -0.44 | **0.001** |
| Year [2001] | -1.09 | -1.81 – -0.38 | **0.003** |
| Year [2002] | -1.06 | -1.78 – -0.35 | **0.004** |
| Year [2003] | -1.05 | -1.76 – -0.33 | **0.004** |
| Year [2004] | -0.97 | -1.68 – -0.25 | **0.008** |
| Year [2005] | -0.97 | -1.69 – -0.26 | **0.008** |
| Year [2006] | -0.93 | -1.64 – -0.22 | **0.011** |
| Year [2007] | -0.83 | -1.55 – -0.12 | **0.022** |
| Year [2008] | -0.83 | -1.54 – -0.11 | **0.023** |
| Year [2009] | -0.70 | -1.42 – 0.01 | 0.053 |
| Year [2010] | -0.63 | -1.34 – 0.09 | 0.085 |
| Year [2011] | -0.59 | -1.30 – 0.13 | 0.107 |
| Year [2012] | -0.53 | -1.25 – 0.18 | 0.142 |
| Year [2013] | -0.44 | -1.16 – 0.27 | 0.225 |
| Year [2014] | -0.40 | -1.12 – 0.31 | 0.269 |
| Year [2015] | -0.31 | -1.02 – 0.40 | 0.393 |
| Year [2016] | -0.24 | -0.96 – 0.47 | 0.505 |
| Year [2017] | -0.16 | -0.87 – 0.56 | 0.665 |
| Year [2018] | -0.12 | -0.83 – 0.60 | 0.748 |
| Year [2020] | -1.08 | -1.80 – -0.37 | **0.003** |
| Year [2021] | -1.30 | -2.02 – -0.59 | **<0.001** |
| age | -0.70 | -0.70 – -0.70 | **<0.001** |
| Gender [Male] | -1.70 | -1.88 – -1.53 | **<0.001** |
| Observations | 6464 | | |
| R^2^ / R^2^ adjusted | 0.969 / 0.969 | | |

Table 22 shows that LE_0_ significantly decreased in 2020-2021 in Mauritania.

**Table 23: Linear regression table of LE_0_ for Niger (1990-2021).**

|  | **ex** | | |
| --- | --- | --- | --- |
| **Predictors** | **Estimates** | **CI** | **p** |
| (Intercept) | 57.96 | 57.46 – 58.46 | **<0.001** |
| Year [1991] | 0.01 | -0.66 – 0.68 | 0.971 |
| Year [1992] | 0.13 | -0.54 – 0.80 | 0.703 |
| Year [1993] | 0.30 | -0.38 – 0.97 | 0.388 |
| Year [1994] | 0.41 | -0.26 – 1.09 | 0.226 |
| Year [1995] | 0.56 | -0.11 – 1.23 | 0.102 |
| Year [1996] | 0.63 | -0.04 – 1.30 | 0.066 |
| Year [1997] | 0.69 | 0.02 – 1.36 | **0.043** |
| Year [1998] | 0.72 | 0.05 – 1.39 | **0.035** |
| Year [1999] | 0.70 | 0.03 – 1.37 | **0.040** |
| Year [2000] | 0.71 | 0.04 – 1.38 | **0.038** |
| Year [2001] | 0.67 | 0.00 – 1.34 | **0.050** |
| Year [2002] | 0.72 | 0.04 – 1.39 | **0.037** |
| Year [2003] | 0.81 | 0.14 – 1.48 | **0.018** |
| Year [2004] | 1.05 | 0.38 – 1.72 | **0.002** |
| Year [2005] | 1.21 | 0.54 – 1.88 | **<0.001** |
| Year [2006] | 1.38 | 0.71 – 2.05 | **<0.001** |
| Year [2007] | 1.55 | 0.88 – 2.22 | **<0.001** |
| Year [2008] | 1.73 | 1.06 – 2.40 | **<0.001** |
| Year [2009] | 1.89 | 1.22 – 2.56 | **<0.001** |
| Year [2010] | 2.11 | 1.44 – 2.78 | **<0.001** |
| Year [2011] | 2.19 | 1.52 – 2.86 | **<0.001** |
| Year [2012] | 2.31 | 1.64 – 2.99 | **<0.001** |
| Year [2013] | 2.43 | 1.76 – 3.10 | **<0.001** |
| Year [2014] | 2.55 | 1.88 – 3.22 | **<0.001** |
| Year [2015] | 2.58 | 1.91 – 3.25 | **<0.001** |
| Year [2016] | 2.72 | 2.05 – 3.39 | **<0.001** |
| Year [2017] | 2.87 | 2.20 – 3.54 | **<0.001** |
| Year [2018] | 2.91 | 2.24 – 3.58 | **<0.001** |
| Year [2019] | 3.03 | 2.36 – 3.70 | **<0.001** |
| Year [2020] | 1.73 | 1.06 – 2.40 | **<0.001** |
| Year [2021] | 1.69 | 1.02 – 2.36 | **<0.001** |
| age | -0.67 | -0.67 – -0.66 | **<0.001** |
| Gender [Male] | -0.87 | -1.04 – -0.71 | **<0.001** |
| Observations | 6464 | | |
| R^2^ / R^2^ adjusted | 0.970 / 0.970 | | |

Table 23 shows that compared to 1990, LE_0_ in Niger increased in the last 30 years and significantly increased from 1997 to the highest value in 2019. LE_0_ then significantly decreased between 2020 and 2021.

**Table 24: Comparison of LE_0_ between males and females in Niger (1990-2021).**

|  | **ex** | | |
| --- | --- | --- | --- |
| **Predictors** | **Estimates** | **CI** | **p** |
| (Intercept) | 60.99 | 60.49 – 61.50 | **<0.001** |
| Year [1990] | -3.03 | -3.70 – -2.36 | **<0.001** |
| Year [1991] | -3.02 | -3.69 – -2.35 | **<0.001** |
| Year [1992] | -2.90 | -3.57 – -2.23 | **<0.001** |
| Year [1993] | -2.74 | -3.41 – -2.07 | **<0.001** |
| Year [1994] | -2.62 | -3.29 – -1.95 | **<0.001** |
| Year [1995] | -2.47 | -3.14 – -1.80 | **<0.001** |
| Year [1996] | -2.40 | -3.07 – -1.73 | **<0.001** |
| Year [1997] | -2.34 | -3.01 – -1.67 | **<0.001** |
| Year [1998] | -2.31 | -2.98 – -1.64 | **<0.001** |
| Year [1999] | -2.33 | -3.00 – -1.66 | **<0.001** |
| Year [2000] | -2.32 | -2.99 – -1.65 | **<0.001** |
| Year [2001] | -2.36 | -3.03 – -1.69 | **<0.001** |
| Year [2002] | -2.32 | -2.99 – -1.65 | **<0.001** |
| Year [2003] | -2.22 | -2.89 – -1.55 | **<0.001** |
| Year [2004] | -1.99 | -2.66 – -1.31 | **<0.001** |
| Year [2005] | -1.82 | -2.49 – -1.15 | **<0.001** |
| Year [2006] | -1.65 | -2.32 – -0.98 | **<0.001** |
| Year [2007] | -1.49 | -2.16 – -0.81 | **<0.001** |
| Year [2008] | -1.30 | -1.97 – -0.63 | **<0.001** |
| Year [2009] | -1.14 | -1.81 – -0.47 | **0.001** |
| Year [2010] | -0.92 | -1.59 – -0.25 | **0.007** |
| Year [2011] | -0.84 | -1.51 – -0.17 | **0.014** |
| Year [2012] | -0.72 | -1.39 – -0.05 | **0.036** |
| Year [2013] | -0.60 | -1.27 – 0.07 | 0.080 |
| Year [2014] | -0.48 | -1.15 – 0.19 | 0.161 |
| Year [2015] | -0.45 | -1.12 – 0.22 | 0.189 |
| Year [2016] | -0.31 | -0.98 – 0.36 | 0.360 |
| Year [2017] | -0.16 | -0.84 – 0.51 | 0.631 |
| Year [2018] | -0.12 | -0.79 – 0.55 | 0.728 |
| Year [2020] | -1.30 | -1.97 – -0.63 | **<0.001** |
| Year [2021] | -1.34 | -2.01 – -0.67 | **<0.001** |
| Age | -0.67 | -0.67 – -0.66 | **<0.001** |
| Gender [Male] | -0.87 | -1.04 – -0.71 | **<0.001** |
| Observations | 6464 | | |
| R^2^ / R^2^ adjusted | 0.970 / 0.970 | | |

Table 24 shows that in 2020 and 2021 LE_0_ in Niger decreased significantly compared to 2019.

**Table 25: Linear regression table of LE_0_ for Nigeria (1990-2021).**

|  | **ex** | | |
| --- | --- | --- | --- |
| **Predictors** | **Estimates** | **CI** | **p** |
| (Intercept) | 53.64 | 53.24 – 54.04 | **<0.001** |
| Year [1991] | -0.19 | -0.73 – 0.35 | 0.485 |
| Year [1992] | -0.25 | -0.78 – 0.29 | 0.368 |
| Year [1993] | -0.21 | -0.75 – 0.33 | 0.443 |
| Year [1994] | -0.42 | -0.96 – 0.11 | 0.122 |
| Year [1995] | -0.49 | -1.03 – 0.04 | 0.071 |
| Year [1996] | -0.53 | -1.07 – 0.01 | 0.052 |
| Year [1997] | -0.52 | -1.06 – 0.02 | 0.057 |
| Year [1998] | -0.52 | -1.06 – 0.01 | 0.055 |
| Year [1999] | -0.35 | -0.88 – 0.19 | 0.207 |
| Year [2000] | -0.19 | -0.73 – 0.34 | 0.477 |
| Year [2001] | -0.13 | -0.67 – 0.41 | 0.632 |
| Year [2002] | -0.13 | -0.67 – 0.41 | 0.633 |
| Year [2003] | -0.02 | -0.56 – 0.52 | 0.946 |
| Year [2004] | -0.00 | -0.54 – 0.53 | 0.987 |
| Year [2005] | 0.12 | -0.42 – 0.65 | 0.673 |
| Year [2006] | 0.19 | -0.34 – 0.73 | 0.480 |
| Year [2007] | 0.22 | -0.31 – 0.76 | 0.418 |
| Year [2008] | 0.21 | -0.33 – 0.74 | 0.453 |
| Year [2009] | 0.36 | -0.18 – 0.90 | 0.189 |
| Year [2010] | 0.40 | -0.14 – 0.94 | 0.144 |
| Year [2011] | 0.54 | 0.00 – 1.08 | **0.049** |
| Year [2012] | 0.55 | 0.02 – 1.09 | **0.043** |
| Year [2013] | 0.61 | 0.08 – 1.15 | **0.025** |
| Year [2014] | 0.62 | 0.08 – 1.15 | **0.024** |
| Year [2015] | 0.60 | 0.06 – 1.13 | **0.029** |
| Year [2016] | 0.64 | 0.10 – 1.18 | **0.020** |
| Year [2017] | 0.71 | 0.17 – 1.25 | **0.009** |
| Year [2018] | 0.76 | 0.22 – 1.29 | **0.006** |
| Year [2019] | 0.85 | 0.31 – 1.38 | **0.002** |
| Year [2020] | 0.44 | -0.10 – 0.98 | 0.107 |
| Year [2021] | 0.09 | -0.44 – 0.63 | 0.737 |
| Age | -0.59 | -0.59 – -0.59 | **<0.001** |
| Gender [Male] | -0.88 | -1.02 – -0.75 | **<0.001** |
| Observations | 6464 | | |
| R^2^ / R^2^ adjusted | 0.975 / 0.975 | | |

Table 25 shows that LE_0_ in Nigeria decreased to 2004 compared to 1990. However, in 2004 it took the same value of the 1990 (estimate =0 in 2004). Again, LE_0_ insignificantly increased from 2005 until it became significant in 2011, attaining the highest value 2019. However, it insignificantly decreased in 2020 and 2021.

**Table 26: Comparison of LE_0_ between males and females in Nigeria (1990-2021).**

|  | **ex** | | |
| --- | --- | --- | --- |
| **Predictors** | **Estimates** | **CI** | **p** |
| (Intercept) | 54.49 | 54.09 – 54.89 | **<0.001** |
| Year [1990] | -0.85 | -1.38 – -0.31 | **0.002** |
| Year [1991] | -1.04 | -1.57 – -0.50 | **<0.001** |
| Year [1992] | -1.09 | -1.63 – -0.56 | **<0.001** |
| Year [1993] | -1.06 | -1.59 – -0.52 | **<0.001** |
| Year [1994] | -1.27 | -1.81 – -0.73 | **<0.001** |
| Year [1995] | -1.34 | -1.88 – -0.80 | **<0.001** |
| Year [1996] | -1.38 | -1.91 – -0.84 | **<0.001** |
| Year [1997] | -1.37 | -1.90 – -0.83 | **<0.001** |
| Year [1998] | -1.37 | -1.91 – -0.83 | **<0.001** |
| Year [1999] | -1.19 | -1.73 – -0.65 | **<0.001** |
| Year [2000] | -1.04 | -1.58 – -0.50 | **<0.001** |
| Year [2001] | -0.98 | -1.51 – -0.44 | **<0.001** |
| Year [2002] | -0.98 | -1.51 – -0.44 | **<0.001** |
| Year [2003] | -0.86 | -1.40 – -0.33 | **0.002** |
| Year [2004] | -0.85 | -1.39 – -0.31 | **0.002** |
| Year [2005] | -0.73 | -1.27 – -0.19 | **0.008** |
| Year [2006] | -0.65 | -1.19 – -0.12 | **0.017** |
| Year [2007] | -0.62 | -1.16 – -0.09 | **0.023** |
| Year [2008] | -0.64 | -1.18 – -0.10 | **0.019** |
| Year [2009] | -0.49 | -1.02 – 0.05 | 0.075 |
| Year [2010] | -0.45 | -0.98 – 0.09 | 0.103 |
| Year [2011] | -0.31 | -0.84 – 0.23 | 0.263 |
| Year [2012] | -0.29 | -0.83 – 0.24 | 0.284 |
| Year [2013] | -0.23 | -0.77 – 0.30 | 0.392 |
| Year [2014] | -0.23 | -0.77 – 0.31 | 0.400 |
| Year [2015] | -0.25 | -0.79 – 0.29 | 0.363 |
| Year [2016] | -0.21 | -0.74 – 0.33 | 0.449 |
| Year [2017] | -0.14 | -0.67 – 0.40 | 0.619 |
| Year [2018] | -0.09 | -0.63 – 0.45 | 0.745 |
| Year [2020] | -0.40 | -0.94 – 0.13 | 0.139 |
| Year [2021] | -0.75 | -1.29 – -0.22 | **0.006** |
| Age | -0.59 | -0.59 – -0.59 | **<0.001** |
| Gender [Male] | -0.88 | -1.02 – -0.75 | **<0.001** |
| Observations | 6464 | | |
| R^2^ / R^2^ adjusted | 0.975 / 0.975 | | |

Table 26 shows that compared to 2019, LE_0_ in Nigeria insignificantly decreased in 2020 but significantly in 2021.

**Table 27: Linear regression table of LE_0_ for Senegal (1990-2021).**

|  | **Ex** | | |
| --- | --- | --- | --- |
| **Predictors** | **Estimates** | **CI** | **P** |
| (Intercept) | 62.30 | 61.79 – 62.81 | **<0.001** |
| Year [1991] | 0.03 | -0.66 – 0.71 | 0.938 |
| Year [1992] | -0.00 | -0.69 – 0.68 | 0.994 |
| Year [1993] | -0.05 | -0.73 – 0.64 | 0.895 |
| Year [1994] | -0.05 | -0.73 – 0.64 | 0.891 |
| Year [1995] | -0.06 | -0.75 – 0.62 | 0.854 |
| Year [1996] | -0.12 | -0.80 – 0.57 | 0.740 |
| Year [1997] | -0.18 | -0.87 – 0.50 | 0.597 |
| Year [1998] | -0.31 | -0.99 – 0.37 | 0.372 |
| Year [1999] | -0.31 | -0.99 – 0.37 | 0.371 |
| Year [2000] | -0.31 | -1.00 – 0.37 | 0.368 |
| Year [2001] | -0.21 | -0.90 – 0.47 | 0.542 |
| Year [2002] | -0.12 | -0.80 – 0.56 | 0.727 |
| Year [2003] | 0.06 | -0.62 – 0.75 | 0.856 |
| Year [2004] | 0.26 | -0.42 – 0.94 | 0.454 |
| Year [2005] | 0.48 | -0.21 – 1.16 | 0.172 |
| Year [2006] | 0.66 | -0.03 – 1.34 | 0.059 |
| Year [2007] | 0.85 | 0.17 – 1.53 | **0.015** |
| Year [2008] | 1.02 | 0.34 – 1.70 | **0.003** |
| Year [2009] | 1.21 | 0.52 – 1.89 | **0.001** |
| Year [2010] | 1.41 | 0.73 – 2.10 | **<0.001** |
| Year [2011] | 1.61 | 0.93 – 2.29 | **<0.001** |
| Year [2012] | 1.59 | 0.91 – 2.28 | **<0.001** |
| Year [2013] | 1.78 | 1.10 – 2.47 | **<0.001** |
| Year [2014] | 1.86 | 1.18 – 2.54 | **<0.001** |
| Year [2015] | 1.97 | 1.29 – 2.65 | **<0.001** |
| Year [2016] | 2.18 | 1.50 – 2.87 | **<0.001** |
| Year [2017] | 2.22 | 1.54 – 2.91 | **<0.001** |
| Year [2018] | 2.32 | 1.64 – 3.00 | **<0.001** |
| Year [2019] | 2.48 | 1.79 – 3.16 | **<0.001** |
| Year [2020] | 1.88 | 1.20 – 2.56 | **<0.001** |
| Year [2021] | 1.21 | 0.53 – 1.90 | **<0.001** |
| Age | -0.70 | -0.71 – -0.70 | **<0.001** |
| Gender [Male] | -1.91 | -2.08 – -1.74 | **<0.001** |
| Observations | 6464 | | |
| R^2^ / R^2^ adjusted | 0.972 / 0.972 | | |

Table 27 shows that LE_0_ in Senegal increased in 1991 than in 1992 however, it began to decrease till 2002. After that it insignificantly increased until 2006. In 2007, it significantly increased up to 2021 with a slight decrease in LE_0_ from 2020 to 2021.

**Table 28: Comparison of LE_0_ between males and females in Senegal over 31 years (1990-2021).**

|  | **Ex** | | |
| --- | --- | --- | --- |
| **Predictors** | **Estimates** | **CI** | **P** |
| (Intercept) | 64.77 | 64.26 – 65.29 | **<0.001** |
| Year [1990] | -2.48 | -3.16 – -1.79 | **<0.001** |
| Year [1991] | -2.45 | -3.13 – -1.77 | **<0.001** |
| Year [1992] | -2.48 | -3.16 – -1.80 | **<0.001** |
| Year [1993] | -2.52 | -3.20 – -1.84 | **<0.001** |
| Year [1994] | -2.52 | -3.21 – -1.84 | **<0.001** |
| Year [1995] | -2.54 | -3.22 – -1.86 | **<0.001** |
| Year [1996] | -2.59 | -3.27 – -1.91 | **<0.001** |
| Year [1997] | -2.66 | -3.34 – -1.98 | **<0.001** |
| Year [1998] | -2.79 | -3.47 – -2.10 | **<0.001** |
| Year [1999] | -2.79 | -3.47 – -2.10 | **<0.001** |
| Year [2000] | -2.79 | -3.47 – -2.11 | **<0.001** |
| Year [2001] | -2.69 | -3.37 – -2.01 | **<0.001** |
| Year [2002] | -2.60 | -3.28 – -1.91 | **<0.001** |
| Year [2003] | -2.41 | -3.10 – -1.73 | **<0.001** |
| Year [2004] | -2.21 | -2.90 – -1.53 | **<0.001** |
| Year [2005] | -2.00 | -2.68 – -1.32 | **<0.001** |
| Year [2006] | -1.82 | -2.50 – -1.14 | **<0.001** |
| Year [2007] | -1.62 | -2.31 – -0.94 | **<0.001** |
| Year [2008] | -1.45 | -2.14 – -0.77 | **<0.001** |
| Year [2009] | -1.27 | -1.95 – -0.59 | **<0.001** |
| Year [2010] | -1.06 | -1.75 – -0.38 | **0.002** |
| Year [2011] | -0.87 | -1.55 – -0.18 | **0.013** |
| Year [2012] | -0.88 | -1.57 – -0.20 | **0.011** |
| Year [2013] | -0.69 | -1.38 – -0.01 | **0.047** |
| Year [2014] | -0.61 | -1.30 – 0.07 | 0.078 |
| Year [2015] | -0.50 | -1.19 – 0.18 | 0.147 |
| Year [2016] | -0.29 | -0.98 – 0.39 | 0.401 |
| Year [2017] | -0.25 | -0.94 – 0.43 | 0.469 |
| Year [2018] | -0.16 | -0.84 – 0.53 | 0.655 |
| Year [2020] | -0.59 | -1.28 – 0.09 | 0.088 |
| Year [2021] | -1.26 | -1.95 – -0.58 | **<0.001** |
| age | -0.70 | -0.71 – -0.70 | **<0.001** |
| Gender [Male] | -1.91 | -2.08 – -1.74 | **<0.001** |
| Observations | 6464 | | |
| R^2^ / R^2^ adjusted | 0.972 / 0.972 | | |

Table 29 shows that 2019 was the year with the highest LE_0_ but insignificantly decreased 2020 but significantly in 2021. During the COVID-19 in 2020 and 2021 LE_0_ significantly decreased compared to 2019; an indication that LE_0_ in 2021 was distant to the highest value.

**Table 29: Linear regression table of LE_0_ for Sierra Leone (1990-2021).**

|  | **ex** | | |
| --- | --- | --- | --- |
| **Predictors** | **Estimates** | **CI** | **p** |
| (Intercept) | 54.98 | 54.47 – 55.48 | **<0.001** |
| Year [1991] | -0.32 | -1.00 – 0.35 | 0.346 |
| Year [1992] | -0.36 | -1.04 – 0.31 | 0.292 |
| Year [1993] | -0.45 | -1.12 – 0.22 | 0.190 |
| Year [1994] | -0.82 | -1.49 – -0.14 | **0.018** |
| Year [1995] | -1.01 | -1.68 – -0.34 | **0.003** |
| Year [1996] | -0.69 | -1.37 – -0.02 | **0.043** |
| Year [1997] | -0.81 | -1.48 – -0.13 | **0.019** |
| Year [1998] | -1.35 | -2.02 – -0.67 | **<0.001** |
| Year [1999] | -1.85 | -2.53 – -1.18 | **<0.001** |
| Year [2000] | -0.61 | -1.28 – 0.06 | 0.076 |
| Year [2001] | -0.49 | -1.17 – 0.18 | 0.152 |
| Year [2002] | -0.33 | -1.00 – 0.35 | 0.344 |
| Year [2003] | -0.21 | -0.88 – 0.47 | 0.545 |
| Year [2004] | -0.09 | -0.77 – 0.58 | 0.789 |
| Year [2005] | 0.03 | -0.64 – 0.71 | 0.920 |
| Year [2006] | 0.37 | -0.30 – 1.04 | 0.283 |
| Year [2007] | 0.66 | -0.01 – 1.33 | 0.055 |
| Year [2008] | 1.00 | 0.33 – 1.68 | **0.004** |
| Year [2009] | 1.32 | 0.65 – 2.00 | **<0.001** |
| Year [2010] | 1.61 | 0.94 – 2.28 | **<0.001** |
| Year [2011] | 1.84 | 1.17 – 2.52 | **<0.001** |
| Year [2012] | 2.05 | 1.38 – 2.73 | **<0.001** |
| Year [2013] | 2.27 | 1.59 – 2.94 | **<0.001** |
| Year [2014] | 2.22 | 1.55 – 2.90 | **<0.001** |
| Year [2015] | 2.43 | 1.76 – 3.10 | **<0.001** |
| Year [2016] | 2.78 | 2.11 – 3.45 | **<0.001** |
| Year [2017] | 2.79 | 2.12 – 3.47 | **<0.001** |
| Year [2018] | 3.16 | 2.49 – 3.84 | **<0.001** |
| Year [2019] | 3.25 | 2.58 – 3.93 | **<0.001** |
| Year [2020] | 2.50 | 1.82 – 3.17 | **<0.001** |
| Year [2021] | 2.49 | 1.82 – 3.16 | **<0.001** |
| age | -0.61 | -0.62 – -0.61 | **<0.001** |
| Gender [Male] | -1.31 | -1.48 – -1.14 | **<0.001** |
| Observations | 6464 | | |
| R^2^ / R^2^ adjusted | 0.965 / 0.965 | | |

Table 29 shows that compared to 1990, LE_0_ in Sierra Leone decreased till 2004 when it became statistically significant from 1994 till 1999. Again from 2005 it started to increase compared to 1990 till 2008 when the LE_0_ gains became statistically significant and reaching the highest value in 2019. During the COVID-19 pandemic (2020-2021), LE_0_ in Sierra Leone decreased significantly.

**Table 30: Comparison of LE_0_ between males and females in Sierra Leone over 31 years (1990-2021).**

|  | **ex** | | |
| --- | --- | --- | --- |
| **Predictors** | **Estimates** | **CI** | **p** |
| (Intercept) | 58.23 | 57.72 – 58.73 | **<0.001** |
| Year [1990] | -3.25 | -3.93 – -2.58 | **<0.001** |
| Year [1991] | -3.58 | -4.25 – -2.90 | **<0.001** |
| Year [1992] | -3.61 | -4.29 – -2.94 | **<0.001** |
| Year [1993] | -3.70 | -4.38 – -3.03 | **<0.001** |
| Year [1994] | -4.07 | -4.74 – -3.39 | **<0.001** |
| Year [1995] | -4.26 | -4.94 – -3.59 | **<0.001** |
| Year [1996] | -3.95 | -4.62 – -3.27 | **<0.001** |
| Year [1997] | -4.06 | -4.73 – -3.39 | **<0.001** |
| Year [1998] | -4.60 | -5.27 – -3.93 | **<0.001** |
| Year [1999] | -5.10 | -5.78 – -4.43 | **<0.001** |
| Year [2000] | -3.86 | -4.54 – -3.19 | **<0.001** |
| Year [2001] | -3.74 | -4.42 – -3.07 | **<0.001** |
| Year [2002] | -3.58 | -4.25 – -2.90 | **<0.001** |
| Year [2003] | -3.46 | -4.13 – -2.79 | **<0.001** |
| Year [2004] | -3.34 | -4.02 – -2.67 | **<0.001** |
| Year [2005] | -3.22 | -3.89 – -2.54 | **<0.001** |
| Year [2006] | -2.88 | -3.56 – -2.21 | **<0.001** |
| Year [2007] | -2.59 | -3.27 – -1.92 | **<0.001** |
| Year [2008] | -2.25 | -2.92 – -1.58 | **<0.001** |
| Year [2009] | -1.93 | -2.60 – -1.26 | **<0.001** |
| Year [2010] | -1.64 | -2.32 – -0.97 | **<0.001** |
| Year [2011] | -1.41 | -2.08 – -0.74 | **<0.001** |
| Year [2012] | -1.20 | -1.87 – -0.53 | **<0.001** |
| Year [2013] | -0.99 | -1.66 – -0.31 | **0.004** |
| Year [2014] | -1.03 | -1.70 – -0.35 | **0.003** |
| Year [2015] | -0.82 | -1.50 – -0.15 | **0.017** |
| Year [2016] | -0.47 | -1.14 – 0.20 | 0.170 |
| Year [2017] | -0.46 | -1.13 – 0.22 | 0.184 |
| Year [2018] | -0.09 | -0.76 – 0.59 | 0.799 |
| Year [2020] | -0.75 | -1.43 – -0.08 | **0.028** |
| Year [2021] | -0.76 | -1.43 – -0.09 | **0.027** |
| age | -0.61 | -0.62 – -0.61 | **<0.001** |
| Gender [Male] | -1.31 | -1.48 – -1.14 | **<0.001** |
| Observations | 6464 | | |
| R^2^ / R^2^ adjusted | 0.965 / 0.965 | | |

Table 30 shows that compared with 2019, LE_0_ decreased significantly between 2020 and 2021 (during the COVID-19 pandemic).

**Table 31: Linear regression table of LE_0_ for Togo over 31 years (1990-2021).**

|  | **ex** | | |
| --- | --- | --- | --- |
| **Predictors** | **Estimates** | **CI** | **p** |
| (Intercept) | 58.24 | 57.77 – 58.71 | **<0.001** |
| Year [1991] | -0.10 | -0.73 – 0.52 | 0.747 |
| Year [1992] | -0.16 | -0.79 – 0.47 | 0.622 |
| Year [1993] | -0.28 | -0.91 – 0.35 | 0.383 |
| Year [1994] | -0.43 | -1.06 – 0.20 | 0.180 |
| Year [1995] | -0.51 | -1.13 – 0.12 | 0.115 |
| Year [1996] | -0.56 | -1.19 – 0.07 | 0.081 |
| Year [1997] | -0.60 | -1.23 – 0.02 | 0.059 |
| Year [1998] | -0.68 | -1.31 – -0.05 | **0.033** |
| Year [1999] | -0.63 | -1.26 – -0.00 | **0.049** |
| Year [2000] | -0.63 | -1.26 – -0.01 | **0.048** |
| Year [2001] | -0.67 | -1.30 – -0.04 | **0.037** |
| Year [2002] | -0.56 | -1.19 – 0.07 | 0.080 |
| Year [2003] | -0.50 | -1.13 – 0.13 | 0.119 |
| Year [2004] | -0.62 | -1.25 – 0.01 | 0.052 |
| Year [2005] | -0.52 | -1.15 – 0.11 | 0.104 |
| Year [2006] | -0.48 | -1.11 – 0.15 | 0.136 |
| Year [2007] | -0.42 | -1.05 – 0.21 | 0.188 |
| Year [2008] | -0.41 | -1.04 – 0.21 | 0.197 |
| Year [2009] | -0.35 | -0.97 – 0.28 | 0.279 |
| Year [2010] | -0.28 | -0.91 – 0.34 | 0.377 |
| Year [2011] | -0.08 | -0.70 – 0.55 | 0.815 |
| Year [2012] | -0.08 | -0.71 – 0.55 | 0.801 |
| Year [2013] | 0.13 | -0.50 – 0.76 | 0.688 |
| Year [2014] | 0.13 | -0.49 – 0.76 | 0.674 |
| Year [2015] | 0.31 | -0.31 – 0.94 | 0.326 |
| Year [2016] | 0.32 | -0.31 – 0.95 | 0.321 |
| Year [2017] | 0.54 | -0.09 – 1.17 | 0.093 |
| Year [2018] | 0.51 | -0.12 – 1.14 | 0.113 |
| Year [2019] | 0.75 | 0.12 – 1.38 | **0.019** |
| Year [2020] | 0.75 | 0.13 – 1.38 | **0.019** |
| Year [2021] | 0.94 | 0.31 – 1.57 | **0.003** |
| age | -0.65 | -0.66 – -0.65 | **<0.001** |
| Gender [Male] | -0.80 | -0.95 – -0.64 | **<0.001** |
| Observations | 6464 | | |
| R^2^ / R^2^ adjusted | 0.972 / 0.972 | | |

Table 31 shows that compared to 1990, LE_0_ in Togo decreased until 2012 when it started increasing until it reached the highest and significant value in 2021. LE_0_ in Togo didn’t decrease during the COVID-19 in 2020 and 2021 instead there were LE_0_ gains.

**Table 32: Comparison of LE_0_ between males and females in Togo over 31 years (1990-2021).**

|  | **ex** | | |
| --- | --- | --- | --- |
| **Predictors** | **Estimates** | **CI** | **p** |
| (Intercept) | 58.99 | 58.52 – 59.46 | **<0.001** |
| Year [1990] | -0.75 | -1.38 – -0.12 | **0.019** |
| Year [1991] | -0.86 | -1.48 – -0.23 | **0.008** |
| Year [1992] | -0.91 | -1.54 – -0.28 | **0.004** |
| Year [1993] | -1.03 | -1.66 – -0.40 | **0.001** |
| Year [1994] | -1.18 | -1.81 – -0.55 | **<0.001** |
| Year [1995] | -1.26 | -1.89 – -0.63 | **<0.001** |
| Year [1996] | -1.31 | -1.94 – -0.68 | **<0.001** |
| Year [1997] | -1.36 | -1.98 – -0.73 | **<0.001** |
| Year [1998] | -1.43 | -2.06 – -0.81 | **<0.001** |
| Year [1999] | -1.38 | -2.01 – -0.75 | **<0.001** |
| Year [2000] | -1.39 | -2.01 – -0.76 | **<0.001** |
| Year [2001] | -1.42 | -2.05 – -0.79 | **<0.001** |
| Year [2002] | -1.31 | -1.94 – -0.69 | **<0.001** |
| Year [2003] | -1.25 | -1.88 – -0.62 | **<0.001** |
| Year [2004] | -1.37 | -2.00 – -0.75 | **<0.001** |
| Year [2005] | -1.27 | -1.90 – -0.65 | **<0.001** |
| Year [2006] | -1.23 | -1.86 – -0.60 | **<0.001** |
| Year [2007] | -1.17 | -1.80 – -0.55 | **<0.001** |
| Year [2008] | -1.17 | -1.79 – -0.54 | **<0.001** |
| Year [2009] | -1.10 | -1.73 – -0.47 | **0.001** |
| Year [2010] | -1.04 | -1.66 – -0.41 | **0.001** |
| Year [2011] | -0.83 | -1.46 – -0.20 | **0.010** |
| Year [2012] | -0.83 | -1.46 – -0.21 | **0.009** |
| Year [2013] | -0.62 | -1.25 – 0.00 | 0.051 |
| Year [2014] | -0.62 | -1.25 – 0.01 | 0.054 |
| Year [2015] | -0.44 | -1.07 – 0.19 | 0.171 |
| Year [2016] | -0.44 | -1.06 – 0.19 | 0.174 |
| Year [2017] | -0.21 | -0.84 – 0.41 | 0.502 |
| Year [2018] | -0.25 | -0.87 – 0.38 | 0.444 |
| Year [2020] | 0.00 | -0.63 – 0.63 | 0.997 |
| Year [2021] | 0.19 | -0.44 – 0.82 | 0.554 |
| age | -0.65 | -0.66 – -0.65 | **<0.001** |
| Gender [Male] | -0.80 | -0.95 – -0.64 | **<0.001** |
| Observations | 6464 | | |
| R^2^ / R^2^ adjusted | 0.972 / 0.972 | | |

Table 32 shows that LE in the Togo across the 31 years progressively increased until it reached the highest level in 2021, In contrast to other countries, LE in Togo over the pandemic period (2020 and 2021) didn’t decrease but stayed approximately close to the value of 2019 compared to 2020 and 2021.

**Table 33: Linear regression table of LE_0_ for all West African countries over 31 years (1990-2021).**

|  | **ex** | | |
| --- | --- | --- | --- |
| **Predictors** | **Estimates** | **CI** | **p** |
| (Intercept) | 55.63 | 55.19 – 56.06 | **<0.001** |
| Year [1991] | -0.04 | -0.62 – 0.54 | 0.894 |
| Year [1992] | -0.05 | -0.64 – 0.53 | 0.854 |
| Year [1993] | -0.03 | -0.62 – 0.55 | 0.908 |
| Year [1994] | -0.18 | -0.76 – 0.40 | 0.541 |
| Year [1995] | -0.20 | -0.78 – 0.38 | 0.502 |
| Year [1996] | -0.21 | -0.79 – 0.37 | 0.480 |
| Year [1997] | -0.18 | -0.76 – 0.41 | 0.553 |
| Year [1998] | -0.18 | -0.76 – 0.40 | 0.548 |
| Year [1999] | -0.08 | -0.66 – 0.50 | 0.793 |
| Year [2000] | 0.04 | -0.54 – 0.62 | 0.890 |
| Year [2001] | 0.07 | -0.51 – 0.65 | 0.818 |
| Year [2002] | 0.09 | -0.49 – 0.68 | 0.751 |
| Year [2003] | 0.18 | -0.40 – 0.77 | 0.533 |
| Year [2004] | 0.24 | -0.34 – 0.82 | 0.417 |
| Year [2005] | 0.37 | -0.22 – 0.95 | 0.217 |
| Year [2006] | 0.47 | -0.11 – 1.05 | 0.112 |
| Year [2007] | 0.54 | -0.04 – 1.13 | 0.067 |
| Year [2008] | 0.60 | 0.02 – 1.18 | **0.044** |
| Year [2009] | 0.75 | 0.17 – 1.33 | **0.011** |
| Year [2010] | 0.84 | 0.25 – 1.42 | **0.005** |
| Year [2011] | 0.97 | 0.39 – 1.55 | **0.001** |
| Year [2012] | 1.02 | 0.44 – 1.61 | **0.001** |
| Year [2013] | 1.10 | 0.52 – 1.68 | **<0.001** |
| Year [2014] | 1.14 | 0.56 – 1.72 | **<0.001** |
| Year [2015] | 1.17 | 0.59 – 1.75 | **<0.001** |
| Year [2016] | 1.26 | 0.68 – 1.85 | **<0.001** |
| Year [2017] | 1.34 | 0.76 – 1.92 | **<0.001** |
| Year [2018] | 1.40 | 0.82 – 1.98 | **<0.001** |
| Year [2019] | 1.50 | 0.92 – 2.08 | **<0.001** |
| Year [2020] | 0.96 | 0.38 – 1.54 | **0.001** |
| Year [2021] | 0.63 | 0.05 – 1.21 | **0.034** |
| age | -0.62 | -0.62 – -0.62 | **<0.001** |
| sex [Male] | -1.09 | -1.24 – -0.94 | **<0.001** |
| Observations | 6464 | | |
| R^2^ / R^2^ adjusted | 0.974 / 0.973 | | |

Table 33 shows a linear regression result for all West African countries. It shows an increase in LE_0_ that was significant compared to 1990. In 2008 (estimate achieved a positive CI and p-value<0.05). In this table, we note that the highest LE_0_ was achieved in 2019 and between 2020 and 2021 LE_0_ in West African countries gradually decreased.

**Table 34: Comparison of LE_0_ between males and females in West Africa over 31 years (1990-2021).**

|  | **ex** | | |
| --- | --- | --- | --- |
| **Predictors** | **Estimates** | **CI** | **p** |
| (Intercept) | 57.13 | 56.69 – 57.56 | **<0.001** |
| Year [1990] | -1.50 | -2.08 – -0.92 | **<0.001** |
| Year [1991] | -1.54 | -2.12 – -0.96 | **<0.001** |
| Year [1992] | -1.56 | -2.14 – -0.98 | **<0.001** |
| Year [1993] | -1.54 | -2.12 – -0.96 | **<0.001** |
| Year [1994] | -1.68 | -2.27 – -1.10 | **<0.001** |
| Year [1995] | -1.70 | -2.28 – -1.12 | **<0.001** |
| Year [1996] | -1.71 | -2.29 – -1.13 | **<0.001** |
| Year [1997] | -1.68 | -2.26 – -1.10 | **<0.001** |
| Year [1998] | -1.68 | -2.26 – -1.10 | **<0.001** |
| Year [1999] | -1.58 | -2.16 – -1.00 | **<0.001** |
| Year [2000] | -1.46 | -2.04 – -0.88 | **<0.001** |
| Year [2001] | -1.43 | -2.02 – -0.85 | **<0.001** |
| Year [2002] | -1.41 | -1.99 – -0.83 | **<0.001** |
| Year [2003] | -1.32 | -1.90 – -0.74 | **<0.001** |
| Year [2004] | -1.26 | -1.84 – -0.68 | **<0.001** |
| Year [2005] | -1.14 | -1.72 – -0.55 | **<0.001** |
| Year [2006] | -1.03 | -1.61 – -0.45 | **0.001** |
| Year [2007] | -0.96 | -1.54 – -0.38 | **0.001** |
| Year [2008] | -0.90 | -1.49 – -0.32 | **0.002** |
| Year [2009] | -0.75 | -1.33 – -0.17 | **0.012** |
| Year [2010] | -0.67 | -1.25 – -0.09 | **0.024** |
| Year [2011] | -0.53 | -1.11 – 0.05 | 0.073 |
| Year [2012] | -0.48 | -1.06 – 0.10 | 0.107 |
| Year [2013] | -0.40 | -0.98 – 0.18 | 0.175 |
| Year [2014] | -0.36 | -0.94 – 0.22 | 0.225 |
| Year [2015] | -0.33 | -0.91 – 0.25 | 0.263 |
| Year [2016] | -0.24 | -0.82 – 0.34 | 0.422 |
| Year [2017] | -0.16 | -0.74 – 0.42 | 0.583 |
| Year [2018] | -0.11 | -0.69 – 0.48 | 0.721 |
| Year [2020] | -0.54 | -1.13 – 0.04 | 0.067 |
| Year [2021] | -0.87 | -1.45 – -0.29 | **0.003** |
| Age | -0.62 | -0.62 – -0.62 | **<0.001** |
| Gender [Male] | -1.09 | -1.24 – -0.94 | **<0.001** |
| Observations | 6464 | | |
| R^2^ / R^2^ adjusted | 0.974 / 0.973 | | |

Table 34 is a linear regression result for West African countries compared to 2019 between males and females. It shows that West African countries reached the highest LE_0_ differences between the sexes in 2019 (all the estimates are negative) and LE_0_ decreased between 2020 – 2021.
